# Insights into human health from phenome- and genome-wide analyses of UK Biobank retinal optical coherence tomography phenotypes

**DOI:** 10.1101/2023.05.16.23290063

**Authors:** Seyedeh Maryam Zekavat, Saman Doroodgar Jorshery, Yusrah Shweikh, Katrin Horn, Franziska G. Rauscher, Sayuri Sekimitsu, Satoshi Kayoma, Yixuan Ye, Vineet Raghu, Hongyu Zhao, Marzyeh Ghassemi, Tobias Elze, Ayellet V. Segrè, Janey L. Wiggs, Markus Scholz, Lucian Del Priore, Jay C. Wang, Pradeep Natarajan, Nazlee Zebardast

**Affiliations:** Department of Ophthalmology, Massachusetts Eye and Ear, Harvard Medical School, Boston, MA, USA; Cardiovascular Research Center, Massachusetts General Hospital, Harvard Medical School, Boston, MA, USA; Program in Medical and Population Genetics and Cardiovascular Disease Initiative, Broad Institute of MIT and Harvard, Cambridge, MA, USA; Departments of Computer Science/Medicine, University of Toronto, Toronto, Canada; Vector Institute for Artificial Intelligence, Toronto, ON, Canada; Department of Computer Science and Electrical Engineering, Massachusetts Institute of Technology, Cambridge, MA, USA; Cardiovascular Imaging Research Center, Massachusetts General Hospital, Harvard Medical School, Boston, MA, USA; Institute for Medical Informatics, Statistics and Epidemiology University of Leipzig, Germany and Leipzig Research Centre for Civilization Diseases (LIFE), Leipzig University, Leipzig, Germany; Tufts University School of Medicine, Boston, MA, USA; Department of Ophthalmology and Visual Science, Yale School of Medicine, New Haven, CT, USA; Computational Biology and Bioinformatics Program, Yale University, New Haven, CT, USA; School of Public Health, Yale University, New Haven, CT, USA; Northern California Retina Vitreous Associates, Mountain View, CA

## Abstract

The human retina is a complex multi-layered tissue which offers a unique window into systemic health and disease. Optical coherence tomography (OCT) is widely used in eye care and allows the non-invasive, rapid capture of retinal measurements in exquisite detail. We conducted genome- and phenome-wide analyses of retinal layer thicknesses using macular OCT images from 44,823 UK Biobank participants. We performed phenome-wide association analyses, associating retinal thicknesses with 1,866 incident ICD-based conditions (median 10-year follow-up) and 88 quantitative traits and blood biomarkers. We performed genome-wide association analyses, identifying inherited genetic markers which influence the retina, and replicated our associations among 6,313 individuals from the LIFE-Adult Study. And lastly, we performed comparative association of phenome- and genome-wide associations to identify putative causal links between systemic conditions, retinal layer thicknesses, and ocular disease.

Independent associations with incident mortality were detected for photoreceptor thinning and ganglion cell complex thinning. Significant phenotypic associations were detected between retinal layer thinning and ocular, neuropsychiatric, cardiometabolic and pulmonary conditions. Genome-wide association of retinal layer thicknesses yielded 259 loci. Consistency between epidemiologic and genetic associations suggested putative causal links between thinning of the retinal nerve fiber layer with glaucoma, photoreceptor segment with AMD, as well as poor cardiometabolic and pulmonary function with PS thinning, among other findings.

In conclusion, retinal layer thinning predicts risk of future ocular and systemic disease. Furthermore, systemic cardio-metabolic-pulmonary conditions promote retinal thinning. Retinal imaging biomarkers, integrated into electronic health records, may inform risk prediction and potential therapeutic strategies.

**One Sentence Summary:** Phenome- and genome-wide associations of retinal OCT images across nearly 50,000 individuals identifies ocular and systemic phenotypes linked to retinal layer thinning, inherited genetic variants linked to retinal layer thickness, and putative causal links between systemic conditions, retinal layer thickness, and ocular disease.

## Introduction

The human retina is an intricate, highly stratified central nervous system tissue responsible for phototransduction and the transmission of neuronal signals to the visual cortex. Existing literature details the interaction between retinal cells and many factors including senescence(*1*), environmental exposure (e.g. smoking(*2*) and diet(*3*)), genetics(*4*), ocular conditions (e.g. glaucoma(*5*) and age-related macular degeneration [AMD](*6*)), cardiometabolic diseases (e.g. diabetes(*7*), hypertension(*8*) and coronary microvascular disease(*9*)), neurological diseases (e.g. dementia(*10, 11*), stroke(*12*) and multiple sclerosis [MS](*13*)), pulmonary diseases (e.g. pulmonary hypertension(*14*), sleep apnea(*15, 16*) and chronic obstructive pulmonary disease [COPD](*17*)) and renal diseases (e.g. chronic kidney disease [CKD](*18*) and glomerulonephropathies(*19, 20*)). Within the human body, the retina is uniquely positioned posterior to optically clear structures, permitting *in vivo* visualization and imaging in ways not possible for any other tissue of its cellular complexity and composition.

The use of retinal findings is longstanding and commonplace in the assessment of both ocular and systemic diseases(*21–23*). Since the turn of the century, high-resolution retinal imaging modalities have been developed and iterated exponentially. Modern spectral domain optical coherence tomography (OCT) allows the non-invasive, rapid capture of retinal measurements in exquisite detail (axial resolution 3-8μm) and is now routinely used in eye care settings, with high patient acceptability and minimal technical training requirements(*24, 25*). The now automated delineation and measurement of OCT layers obviates the need for time-intensive manual layer segmentation, and is accurate and highly repeatable(*26*). This combined with the known physiological interactions between the retina and other organ systems makes retinal layer thickness measurement a potentially attractive addition to diagnostic, risk stratification, disease surveillance and treatment response assessment methods.

Quantitative associations between retinal structure and systemic diseases demonstrate the predictive value of retinal measurements and have been particularly well characterized for cardiovascular and neurodegenerative diseases(*11, 27–30*). Analyses using large, deeply-phenotyped datasets with associated retinal imaging offer the potential to gain new insights into pathophysiological processes and disease risk factors, as well as opportunities to identify novel biomarkers and therapeutic targets.

The first genome-wide association study (GWAS) of macular thickness using UK Biobank data was published in 2019 and identified 139 significant loci(*31*). Top macular thickness genes were shown to be present in the retina using gene expression data and many variants were associated with ocular and systemic diseases, including AMD, neurodegenerative conditions, cancer, and metabolic traits(*31*). More recently, 46 loci associated with inner retinal layer thickness have been reported(*32*). However, GWAS across specific cellular layers of the retina has not been performed, and epidemiologic and causal links between retinal layer thicknesses and phenome-wide associations remain under-explored at present.

This article summarizes the results from a phenome-wide association study (PheWAS), GWAS, and complementary genetic discovery and comparative genomic and epidemiologic analyses to investigate the associations between retinal layer thicknesses from 44,823 UK Biobank participants and 1,866 incident international classification of diseases (ICD)-based conditions (median of 10 years of follow up), 88 quantitative clinical and blood traits, 168 plasma metabolites, and over 13 million genetic variants. In summary, the results we present offer opportunities to better understand disease mechanisms, identify ocular biomarkers and discover novel treatment targets for ocular and systemic diseases.

## Methods

### UK Biobank cohort and sample exclusion

The UK Biobank is a prospective population-based cohort study of 502,649 adults aged 40-69 recruited from 2006-2010 with existing genomic and longitudinal phenotypic data (median 10 years of follow-up)(*33*). Comprehensive baseline assessments were conducted at 22 assessment centres across the UK, consisting of questionnaires, physical measurements and biological sample collections including blood-derived DNA. Retinal imaging was performed at enrollment using the Topcon 3D OCT 1000 Mark II (Topcon GB, Newberry, Berkshire, UK) instrument which captures a 3D scan and photograph of the retina. Data use was approved by the Massachusetts General Hospital Institutional Review Board (protocol 2021P002040) and facilitated through UK Biobank Applications 7089 and 50211.

Of the 67,339 genotyped individuals with available retinal OCT imaging at enrollment, we analyzed data from 44,823 consenting participants with good quality scans of white British ancestry with genotypic-phenotypic sex concordance. We randomly excluded one from each pair of first- or second-degree relatives. Poor-quality images and image outliers were excluded as detailed below.

### Retinal OCT and retinal layer segmentation

Nonmydriatic spectral domain OCT scans of the macula were obtained using Spectral Domain Topcon 3D OCT 1000 Mark II (Topcon GB, Newberry, Berkshire, UK). Three-dimensional 6x6 mm^2^ macular volume scans were obtained (512 horizontal A-scans per B-scan; 128 B-scans in a 6x6 mm raster pattern). The right eye of each participant was imaged first, followed by the left eye. All OCT images were stored as .fds image files without prior analysis of macular thickness. We used the Topcon Advanced Boundary Segmentation (TABS) algorithm to automatically segment all scans, which uses dual-scale gradient information to allow for automated segmentation of the inner and outer retinal boundaries and retinal sublayers(*34*). The boundaries segmented are the internal limiting membrane (ILM), nerve fiber layer (NFL), ganglion cell layer (GCL), inner plexiform layer (IPL), inner nuclear layer (INL), external limiting membrane (ELM), photoreceptor inner segment/outer segment (IS/OS) junction, retinal pigment epithelium (RPE), Bruch’s membrane (BM) and choroid-sclera interface (CSI). The software provides an image quality score and segmentation indicators which were used for quality control. Segmentation indicators included the ILM indicator; a measure of the minimum localized edge strength around the ILM boundary across the scan, which can be used to identify blinks, scans that contain regions of signal fading, and errors in segmentation(*35*). We excluded all images with image quality scores less than 40 and images representing the poorest 10% as designated by the ILM indicator. We also excluded any image with a layer thickness greater than 2.5 standard deviations away from the mean. The thickness of each retinal sub-layer was determined by calculating the difference between boundaries of interest and averaging this across all scans. For example, retinal nerve fiber layer (RNFL) thickness was calculated as the difference between ILM and NFL boundary lines.

### UK Biobank array genotyping, whole exome sequencing and quality control

Genome-wide genotyping of blood-derived DNA was performed by UK Biobank for 488,377 individuals using two genotyping arrays sharing 95% marker content: Applied Biosystems UK BiLEVE Axiom Array (807,411 markers in 49,950 participants) and Applied Biosystems UK Biobank Axiom Array (825,927 markers in 438,427 participants), both produced by Affymetrix (Santa Clara, CA, USA)(*33*). Variants used in the present analysis include those also imputed using the Haplotype Reference Consortium reference panel of up to 39 million bi-allelic variants and 88 million variants from the UK10K+1000 Genomes reference panels)(*33*). Poor quality variants and genotypes were filtered as previously described(*33*), with additional filters including high-quality imputed variants (INFO score >0.4), minor allele frequency >0.005, and Hardy-Weinberg Equilibrium P≥1x10^-10^, as previously implemented using Hail-0.2 (https://hail.is/docs/0.2/index.html)(36–38).

UK Biobank whole exome sequencing was performed for 200,627 individuals using blood-derived DNA at the Regeneron Sequencing Center(*39*); the methods of which have been previously described for the earlier data release for approximately 50,000 individuals(*40*). In brief, the IDT xGen Exome Research Panel v1.0 was used to capture exomes at over 20x coverage across 95% of sites. Extensive additional genotype, variant and sample-level exclusion filters were applied to study high-quality autosomal exome sequence variants as previously described(*39, 41*) using Hail-0.2 (**Supplementary Note 1**).

### OCT layer phenome-wide association study (OCT-PheWAS)

Four sets of OCT-PheWAS analyses were performed, corresponding to the association of retinal layer thicknesses with: 1) prevalent phenotypes at enrollment 2) incident phenotypes developed following enrollment 3) quantitative systemic biomarkers and 4) quantitative ocular traits.

OCT-PheWAS with prevalent and incident phenotypes was performed across all of the 1,866 hierarchical phenotypes defined from the Phecode Map 1.2(*42*) ICD-9 (https://phewascatalog.org/phecodes) and ICD-10 (https://phewascatalog.org/phecodes_icd10) phenotype groupings(*43*). Associations of retinal layer thicknesses with prevalent phenotypes were performed using logistic regression models, and associations with incident phenotypes were performed using Cox proportional hazards models after excluding individuals with the corresponding diagnosis at or prior to enrollment. Both models were adjusted for age, age^2^, sex, smoking status (current/prior/never smoker), height, weight, spherical equivalent, and self-reported ethnicity (Data field 21000, https://biobank.ctsu.ox.ac.uk/crystal/field.cgi?id=21000). These covariates were used to highlight disease-related states associated with retinal layer thickness and remove natural differences conferred by age, sex, smoking, ethnicity, body habitus, and orbit size. We used both age and age-squared in our models to account for both linear and quadratic relationships with age. We utilized sex, smoking status, and self-reported ethnicity as these are important confounders to use in conventional epidemiologic analyses and were found to have significant association with the retinal layer thicknesses as observed in Supplementary Table 2. Furthermore, we added on height and weight as covariates given body size was also observed to associated with retinal layer thicknesses (as highlighted in Supplementary Figure 2 and Supplementary Table 2). Indeed, a larger body habitus is associated with overall thicker retinal layers as visualized in Supplementary Figure 2. Lastly, adjustment for spherical equivalent was performed as the size of the orbit, ie: how myopic or hyperopic one is, also stretches out the orbit thereby influencing the retinal layer thicknesses (as visualized in Supplementary Figure 2 and quantified in Supplementary Table 2). The proportional hazards assumption was assessed by Schoenfeld residuals and was satisfied for each model. Analysis was performed across disease phenotypes. Statistical significance was defined using a false discovery rate (FDR) of <0.05.

OCT-PheWAS across 88 quantitative systemic biomarkers measured at enrollment, including blood counts (Category ID 100081), blood biochemistry markers (Category ID 17518), liver MRI, iron and inflammation measurements (Category 126), arterial stiffness and reflection index measurements from finger photoplethysmography (Category 100007), blood pressure (Category 100011), pulmonary function results from spirometry (Category 100020), left ventricular size and function and pulse wave analysis from cardiac MRI (Category 102), and eye measurements (Category 100011) were performed. For all phenotypes, sex-specific extreme outliers were excluded by adjusting the traditional box and whisker upper and lower bounds and accounting for skewness in the phenotypic data using the Robustbase package in R (setting range=3) as previously performed(*36–38*) (https://cran.r-project.org/web/packages/robustbase/robustbase.pdf). Quantitative traits were inverse rank normalized to mean 0 and standard deviation 1. Analyses were performed using linear regression in models adjusted for age, age^2^, sex, smoking status (current/prior/never smoker), height, weight, spherical equivalent, and ethnicity. Statistical significance was defined using a false discovery rate of <0.05.

### GWAS and secondary *in silico* analyses

GWAS was performed using Hail-0.2 software (https://hail.is/) on the Google cloud for individuals with retinal OCT imaging at enrollment. A linear regression model was used for analysis, adjusting for age, age^2^, sex, smoking, spherical equivalent, the first ten principal components of genetic ancestry, and genotyping array.

Using the GWAS summary statistics, SNP heritability analysis was performed using LD-score regression with LDSC-v1.0.1 (https://github.com/bulik/ldsc) and European LD scores from 1000 Genomes(*44*). Putative causal genes were prioritized using PoPS software(*45*) (https://github.com/FinucaneLab/pops), which integrates GWAS summary statistics with gene expression, biological pathways, and predicted protein-protein interaction data to identify likely causal genes at each genome-wide significant locus (combines information from gene expression data from 73 publicly available bulk RNASeq and scRNASeq databases including from retinal tissues, 8,479 biological pathways, and 8,718 protein-protein interactions). Enrichment analysis was performed using the FUMA and EnrichR web browsers across genes with a PoPS z-score>1. Genetic correlation analysis was performed using GNOVA, using GWAS summary statistics from the datasets listed in the legend of **Supplementary Table 12**.

### Replication of genome-wide significant associations in the LIFE-Adult-Study

The population-based cohort LIFE-Adult-Study (Leipzig Research Center for Civilization Diseases) examined 10,000 randomly selected participants (registry office) from the city of Leipzig, Germany(*46*). The sample was recruited stratified by age and sex. 9,600 participants were in the age group between 40 and 79 years and 400 between 18 and 39 years. The study investigates the effect of molecular-genetic, environmental and life-style factors on the prevalence and incidence of major civilization diseases (ex: diabetes mellitus, obesity, allergies, cardiovascular diseases). All subjects participated in a comprehensive examination program including structured medical interviews, questionnaires, physical and medical examination, and clinical chemistry (*46*).

Prior to participation, all participants provided written informed consent. The study was approved by the Ethics Committee of the Medical Faculty of Leipzig University, and the research was conducted in accordance with the Declaration of Helsinki.

Spectral-domain optical coherence tomography (SD-OCT, Spectralis, Heidelberg Engineerníng, Heidelberg, Germany) scans of the macular area with a field size of 20° (temporal-nasal) x 20°(superior-inferior) were acquired in High Speed mode. Every B-scan contained 512 A-scans, corresponding to a distance between single A-scans of 0.039°. 97 B-scans were acquired within the specified field resulting in a distance between individual B-scans of ∼0.205°. Data was segmented by built-in Heyex algorithm yielding the following thicknesses: retinal nerve fiber layer (RNFL); ganglion cell layer (GCL); inner plexiform layer (IPL); inner nuclear layer (INL); outer plexiform layer (OPL); outer nuclear layer (ONL) which includes thickness of external limiting membrane (ELM) as layer is segmented by Spectralis SD-OCT software at the inferior border of the bright band (ELM); myoid zone (MZ); ellipsoid zone (EZ) and outer-photoreceptor segment (OS) combined (named EZ+OS here) as segmentation by Spectralis SD-OCT software of this zone/ this section consists of a bright (EZ) and a dark (OS) band; interdigitation zone (IZ); retinal pigment epithelium (RPE). These layers were summarized as follows to match the UK Biobank TOPCON data, resulting in the following analyzed layers: RNFL, GCL, IPL, INL, OPL+ONL, PS as computed by MZ+ EZ+OS+IZ, and RPE; furthermore GCC was computed by RNFL+GCL+IPL.

The measurements of the thickness of the retinal layers were made separately for the left and right eye. In order to perform an analysis for the average thickness of the retinal layers, outliers were first removed (mean +/- 3 SD). After determination of the residuals with adjustment to the spherical equivalent, the mean value was determined and used after scaling for the GWAS with PLINK2. Ultimately, 6313 unrelated samples with measurements of retinal layer thickness were included in analyses across the 259 top genome-wide significant loci identified in the UK Biobank.

### Polygenic risk score (PRS) development

Polygenic risk scores (PRS) for the retinal layer thicknesses were developed among individuals not included in the GWAS study (i.e. genotyped UK Biobank participants without retinal OCT images available). This method of excluding individuals used in the GWAS in the PRS-PheWAS is done to minimize bias in interpretation of downstream PRS-PheWAS analysis such that the compared epidemiologic OCT-PheWAS analyses and genetic PRS-PheWAS analyses represent results from distinct and non-overlapping sets of individuals. Significant, independent loci were identified using variants with P<5x10^-8^, clumped in Plink-2.0 using an r^2^ threshold of 0.1 across 1-MB genomic windows from the 1000 Genomes Project European reference panel. Additive PRSs for each separate retinal layer thickness were developed as follows: 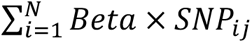, where *Beta* is the weight for each of the *N* independent genome-wide significant variants in the GWAS, and *SNP*_*ij*_ is the number of alleles (i.e. 0, 1, or 2) for *SNP*_*ij*_ carried by individual *j* in the UK Biobank. Further sensitivity analyses were performed to assess association of the retinal layer thickness PRS and possible environmental confounders other than those included as covariates in the analysis (ex: socioeconomic status as indicated through Townsend deprivation index, alcohol intake, diet, stress, exercise).

Additionally, we created polygenic risk scores (PRS) for clinical quantitative traits previously detailed (88 quantitative systemic biomarkers measured at enrollment, including blood counts (Category ID 100081), blood biochemistry markers (Category ID 17518), liver MRI, iron and inflammation measurements (Category 126), arterial stiffness and reflection index measurements from finger photoplethysmography (Category 100007), blood pressure (Category 100011), pulmonary function results from spirometry (Category 100020), left ventricular size and function and pulse wave analysis from cardiac MRI (Category 102), and eye measurements (Category 100011)) used in a PRS-PheWAS as follows. We restricted the analysis to variants present in the hapmap3 dataset (∼1.5 million variants) and excluded variants within major histocompatibility complex locus (defined as chromosome 6: base pairs 25,000,000-35,000,000). First, we inverse-rank normalized and regressed out the phenotypic values for the covariates (age, age-squared, genetic sex, usage of lipid-lowering medicine, usage of anti-diabetic medicine, smoking status, and alcohol consumption status). Using these normalized quantitative phenotypes, we estimated the effect sizes of each variant through genome-wide association study by BOLT-LMM (ver2.3.5), adjusting for genotyping array among European ancestry ascertained by K-means clustering (K=4) on the first four genetic principal components (as done previously here: https://research-information.bris.ac.uk/en/datasets/mrc-ieu-uk-biobank-gwas-pipeline-version-2). Samples included in the retinal OCT analyses were excluded from the genome-wide analyses to prevent sample overlap. Then, we applied the pruning and thresholding method to choose the best parameters by PLINK software. The tested parameters were R^2^ = (0.2, 0.4, 0.6, 0.8), P = ([0, 5 × 10^-8^], [0, 5 × 10^-6^], [0, 5 × 10^-5^], [0, 5 × 10^-2^], [0, 1]) on the holdout population in UKBB. We tested the performance of PRSs on the independent validation cohort (the second holdout UKBB population, N ≤ 5,000), using a linear regression model adjusting for age, age-squared, and the first ten principal components. We chose the combination of the R^2^/P-value cutoff parameters which attained the smallest P-value in the regression model on the validation cohort (provided in **Supplementary Table 15**).

### Polygenic risk score (PRS) Phenome-Wide Association Study

PRS-PheWAS was performed to assess for potential causal quantitative systemic traits that concordantly influence retinal layer thicknesses epidemiologically and genetically. Phenome-wide associations of the retinal layer thickness PRS were performed to test the associations between genetically influenced layers across combined prevalent and incident phenotypes from PheCode Map 1.2(*42*) ICD-9 and ICD-10 codes, adjusted for age, age^2^, sex, smoking, the first ten principal components of genetic ancestry, and genotyping array. Given slight differences in the magnification of OCT images correlated with the presence and severity of myopia (spherical equivalent), we also performed sensitivity analyses adjusting for spherical equivalent, as previously described(*32*).

Additionally, phenome-wide associations of the systemic quantitative trait PRS were performed to test their association with retinal layer thickness, adjusting for age, age^2^, sex, smoking, the first ten principal components of genetic ancestry, and genotyping array.

### Rare variant association study (RVAS)

RVAS was performed among unrelated individuals with both whole exome sequencing (WES) and retinal OCT images available using burden tests through the REGENIE package(*47*) (https://rgcgithub.github.io/regenie/options/). Exonic variants were filtered as rare (minor allele frequency <1%) variants, predicted high-confidence loss-of-function variants by LOFTEE(*48*) or predicted missense deleterious variants by MetaSVM(*49*), and grouped by protein-coding gene. RVAS gene burden analysis was conducted, adjusting for age, age^2^, sex, ever smoking, and the first ten principal components of genetic ancestry. Significance was defined based on a Bonferroni cutoff dependent upon the number of genes analyzed (P<0.05/1176*7=*4.2x10^-6^ for retinal layer thickness).

## Results

### Baseline characteristics

Data from 44,823 UK Biobank participants with genotyped samples and retinal OCT images were included in analyses. Mean age was 56.8 (SD 7.9) years and 24,058 (53.7%) were female. While all individuals included in our analyses were of white, British ancestry as determined through their genotypic data, 1.3% of participants self-reported their ethnicity as either South Asian (N=40, 0.1%) or as Black or Mixed race (N=530, 1.2%). 4,291 (9.6%) were tobacco smokers at enrollment, 16,199 (36.1%) reported previously smoking and 24,333 (54.3%) reported no history of smoking. Mean BMI was 27.2 (SD 4.7) kg/m^2^. Prevalent systemic conditions included 951 (2.1%) participants with type 2 diabetes, 12,843 (28.7%) hypertension, 2,292 (5.1%) coronary artery disease, 7,452 (16.6%) hypercholesterolemia, 143 (0.3%) chronic kidney disease, and 639 (1.4%) stroke. The prevalence of ocular conditions was low overall: the number of participants with a self-reported or diagnosed history of cataract was 1,585 (3.5%), 624 (1.4%) for glaucoma, 512 (1.1%) for AMD, and 108 (0.2%) for retinal detachment **(Supplementary Table 1)**.

### Quantification of retinal layer thicknesses from OCT imaging segmentation

The mean OCT retinal layer thicknesses (N=44,823), as determined by automatic segmentation was 102.2μm (SD 8.2) for the ganglion cell complex (GCC), 40.0μm (SD 4.7) for the retinal nerve fiber layer (RNFL), 33.3μm (SD 2.8) for the ganglion cell layer (GCL), 28.9μm (SD 2.57) for the inner plexiform layer (IPL), 30.9μm (SD 2.0) for the inner nuclear layer (INL), 77.4μm (SD 5.8) for the outer plexiform layer (OPL), 63.3μm (SD 2.7) for the photoreceptor segment (PS) layer, 23.2μm (SD 1.8) for the retinal pigment epithelium-Bruch’s membrane complex (RPE-BM) and 200.3μm (SD 46.1) for the choroid-scleral interface (CSI) **(Supplementary Table 1, Figure 1a)**.

**Figure 1:**
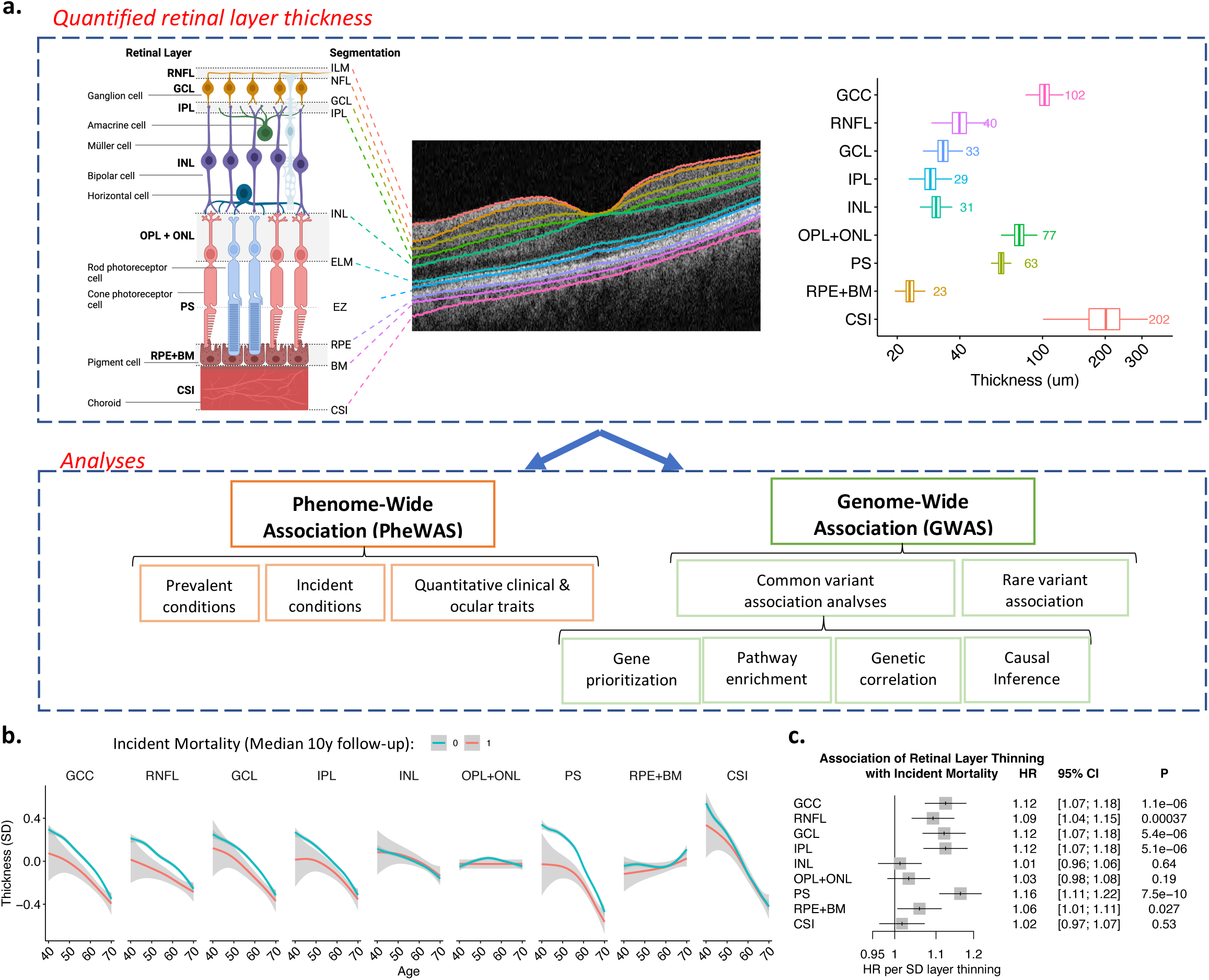
Study schematic and retinal layer thickness measurements. a. Here, we first quantified the thickness of 9 retinal layers across all individuals with retinal OCT imaging data available in the UK Biobank (N=44,823). We then used these measurements to perform phenome- and genome-wide association analyses to identify what phenotypes are associated with the different layers of the retina, and what genetic variants influence these layers. Measured thickness of the retinal layers in microns is provided. b. Relationship of normalized retinal layer thickness with age stratified by incident mortality (1,746 cases, 39,188 controls) across a median 10-year follow-up period. c. Association of retinal layer thinning with incident mortality, adjusted for age, age^2^, sex, height, weight, ethnicity, and spherical equivalent. ILM = Internal limiting membrane, NFL = nerve fiber layer, GCC = ganglion cell complex or RNFL+GCL+IPL, GCL = ganglion cell layer, IPL = inner plexiform layer, INL = inner nuclear layer, ELM = external limiting membrane, EZ = ellipsoid zone, also known as photoreceptor inner segment/outer segment junction, RPE = retinal pigment epithelium, BM = Bruch’s membrane, CSI = choroid-sclera interface, RNFL = retinal nerve fiber layer, OPL+ONL = outer plexiform layer and outer nuclear layer, PS = photoreceptor segment, RPE+BM = retinal pigment epithelium and Bruch’s membrane complex.

The calculated retinal layer thicknesses were highly correlated between right and left eyes as shown in **Supplementary Figure 1**. Mean retinal layer thicknesses were greater for left eyes compared to right for all retinal layers except for the GCL with a mean right-left difference of 0.04μm (P=6.8x10^-6^).

Other retinal layers reaching statistical significance for interocular asymmetry were the RNFL with a mean right-left difference of -0.05μm (P=0.01), the OPL+ONL with a mean right-left difference of -0.29μm (P=4.6x10^-140^), the PS with a mean right-left difference of -0.05μm (P=4.4x10^-10^), the RPE-BM complex with a mean right-left difference of -0.04μm (P=1.2x10^-5^) and the CSI with a mean right-left difference of -0.72μm (P=8x10^-7^) **(Supplementary Figure 1)**.

Univariate associations of retinal layer thicknesses with sex, smoking status, height, and weight are shown in **Supplementary Figure 2,** and multivariate associations are provided in **Supplementary Table 2**. As seen, all retinal layers tend to thin with age, except for the RPE+BM layer which tends to thicken after approximately age 60 (**Supplementary Figure 2**), possibly related to the accumulation of drusenoid deposits in the BM with age. Additionally, most layers tend to get thicker with increased height while more variable directionality of univariate associations are seen with weight (**Supplementary Figure 2b**). Additionally, more myopic eyes (i.e. more negative spherical equivalent) tend to have thinner retinal layers across all layers except the RPE+BM and RNFL layers, which are thicker both in univariate and multivariate models (**Supplementary Figure 2b, Supplementary Table 2**). Smoking was associated with significant thinning of all layers in multivariate models, except for the RPE+BM layer which showed no significant association with smoking status (**Supplementary Table 2**). Finally, in multivariate models, male sex was associated with thinner inner layers (GCC, RNFL, GCL) but thicker outer INL, OPL, PS, and RPE layers (**Supplementary Table 2**). While only 1.3% of the individuals of genetically-determined white British Ancestry included in our analyses did not self-identify as white, as seen in **Supplementary Table 2**, self-reported ethnicity categories were associated with retinal layers thickness (GCC, GCL, OPL, PS, RPE+BM, CSI), supporting inclusion as covariates in analysis. Expected univariate associations were also detected between thinning of the GCC and associated layers (RNFL, GCL, IPL) and increased incidence of glaucoma, as well as between thinning of the PS layer and thickening of the RPE+BM layer and increased incidence of age-related macular degeneration (AMD) (**Supplementary Figure 3**).

### OCT Layer phenome-wide association study (OCT-PheWAS)

We first associated retinal layer thicknesses on OCT with incident mortality and found that the incidence of mortality over a 10 year follow up period was greatest among individuals with thinner PS layers (HR 1.16, 95% CI 1.11-1.22, P=7.5x10^-10^) and thinner GCC layers (HR 1.12, 95% CI 1.07-1.18, P=1.1x10^-6^) at baseline, after adjusting for age, age^2^, sex, height, weight, smoking status, ethnicity and spherical equivalent **(Figure 1b and 1c)**. PS thinning and GCC thinning are positively correlated with incident mortality for participants stratified by age in decades as shown in **Supplementary Figure 4,** highlighting the heightened risk of mortality within each age decile among individuals with either thin (<2SD lower than the mean) PS or GCC layers. The associations between retinal layer thinning and incident mortality are detailed for each layer in unadjusted and adjusted analyses shown in **Supplementary Figure 5**. For both PS and GCC layer thinning, the significant associations with incident mortality persist in the maximally adjusted models, adjusted for age, age^2^, sex, height, weight, self-reported ethnicity, spherical equivalent, hemoglobin A1c (HbA1c), systolic blood pressure, diastolic blood pressure, and incident coronary artery disease (**Supplementary Figure 5**).

OCT layer phenome-wide association was then conducted separately for retinal layer thicknesses across 1,866 prevalent (**Supplementary Table 3, Figure 2a**) and incident (**Supplementary Table 4, Figure 2b**) phenotypes (phecodes)(*42*) which combined ICD-9 and ICD-10 groupings, as well as quantitative clinical traits and serological biomarkers **(Supplementary Table 5, Figure 3)**. All analyses were adjusted for age, age^2^, sex, height, weight, mean spherical equivalent, smoking status and self-reported ethnicity. Significant associations were identified between retinal layer thinning and both ocular and systemic conditions, as detailed below.

**Figure 2:**
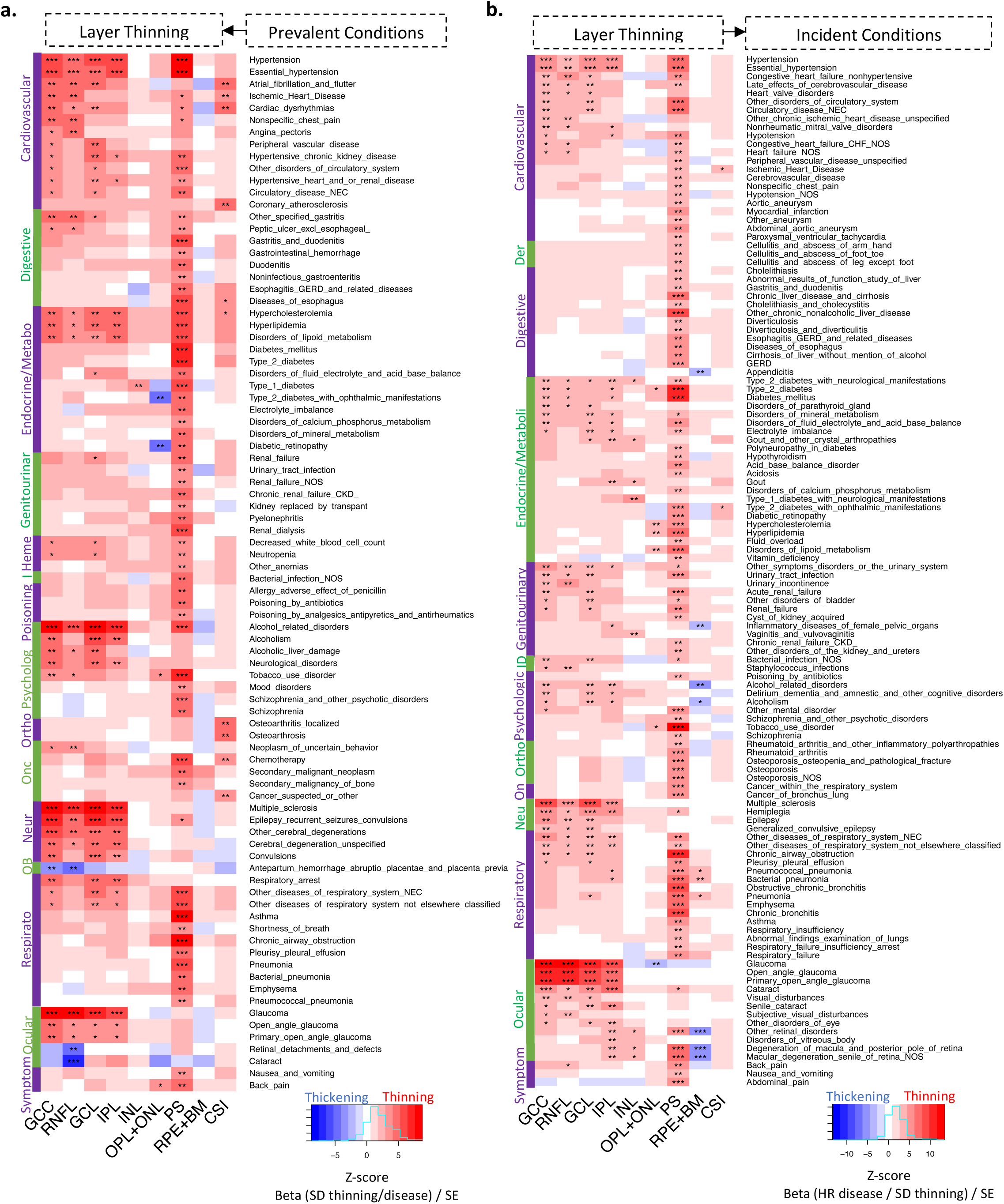
Phenome-wide association study of retinal layer thickness with prevalent and incident disease. a. Association of prevalent disease (i.e., disease present at time of retinal imaging) with retinal layer thinning in logistic regression models. b. Association of retinal layer thickness with incident disease (i.e., disease that developed after retinal imaging during follow-up), in Cox Proportional hazard models. All analyses are adjusted for age, age^2^, sex, height, weight, ethnicity, and spherical equivalent. Across both panels, phenotypes with at least one layer having false discovery rate (FDR)<0.01 were included in the plots. Within cells, association strength is denoted as such: ***FDR<1x10-4, **1x10-4≤FDR<0.01, *0.01≤FDR<0.05. The color reflects the z-score (or beta in units of SD thinning/standard error) for association, where red reflects layer thinning and blue reflects thickening. Full results are available in Supplementary Tables 3,4. GCC = ganglion cell complex or RNFL+GCL+IPL, RNFL = retinal nerve fiber layer, GCL = ganglion cell layer, IPL = inner plexiform layer, INL = inner nuclear layer, OPL+ONL = outer plexiform layer and outer nuclear layer, PS = photoreceptor segment, RPE+BM = retinal pigment epithelium and Bruch’s membrane complex, CSI = choroid layer.

**Figure 3:**
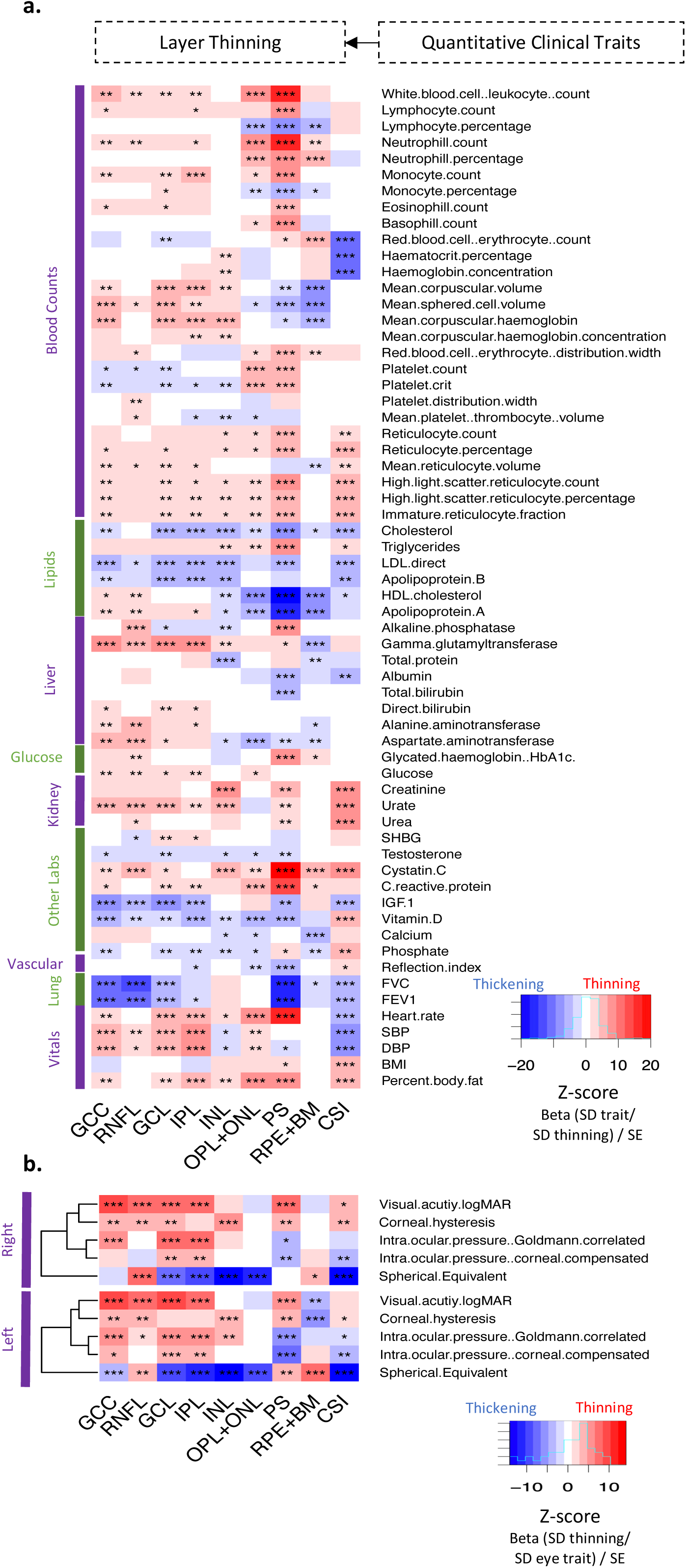
Phenome-wide association study of quantitative clinical traits with retinal layer thinning. Association of quantitative a. clinical and b. ocular traits acquired at time of imaging with retinal layer thinning in linear regression models. All analyses are adjusted for age, age^2^, sex, height, weight, ethnicity, and spherical equivalent. Across both panels, phenotypes with at least one layer having false discovery rate (FDR)<0.01 were included in the plots. Within cells, association strength is denoted as such: ***FDR<1x10-4, **1x10-4≤FDR<0.01, *0.01≤FDR<0.05. The color reflects the z-score (or beta in units of SD quantitative trait/ SD thinning/standard error) for association, where red reflects layer thinning and blue reflects thickening. Full results are available in Supplementary Table 5. GCC = ganglion cell complex or RNFL+GCL+IPL, RNFL = retinal nerve fiber layer, GCL = ganglion cell layer, IPL = inner plexiform layer, INL = inner nuclear layer, OPL+ONL = outer plexiform layer and outer nuclear layer, PS = photoreceptor segment, RPE+BM = retinal pigment epithelium and Bruch’s membrane complex, CSI = choroid layer.

Expected associations were seen for GCC thinning and prevalent and incident glaucoma, as well as for PS thinning and incident macular degeneration (**Figure 2**). The strongest effect estimates was observed for the association between each SD of RNFL thinning and prevalent primary open angle glaucoma (POAG) (OR=4.20, P=1.6x10^-5^, 95% CI 2.20-8.05) as well as incident POAG (HR=2.52, P=3.0x10^-24^, 95% CI 2.11-3.03). Accordingly, each SD higher Goldmann-correlated intraocular pressure (IOPg) was significantly associated with a thinner GCL layer of the retina (β=-0.05 SD, P=8.1x10^-24^, 95% CI -0.15-0.05). Furthermore, each SD of thinner PS was also associated with increased risk of incident macular degeneration (HR=1.39, P=1.3x10^-10^, 95% CI 1.26-1.54).

Thinner retinal layers were also significantly associated with higher odds of a range of prevalent and incident cardiometabolic phenotypes and related conditions. Each 1 SD of retinal inner layers (GCC, RNFL, GCL, IPL) as well as thinner PS was associated with higher odds of having a history of hypertension and a greater risk of developing incident hypertension. The strongest risk of prevalent essential hypertension was observed in participants with thinner PS (OR=1.18, P=7.2x10^-24^, 95% CI 1.15-1.22); concordant associations were also observed with systolic and diastolic blood pressures (**Supplementary Table 5**). The largest effect size between thinner retinal layers and incident hypertension was also observed with the PS layer (HR=1.09, P=5.9x10^-10^, 95% CI 1.06-1.12). Of all retinal layers, thinning of the PS layer had the strongest and most significant associations with prevalent (OR=1.18, P=1.2x10^-11^, 95% CI 1.13-1.24) and incident (HR=1.10, P=2.0x10^-8^, 95% CI 1.06-1.14) hypercholesterolemia. In keeping with these findings, a thinner PS layer also had the strongest associations with prevalent hypertensive chronic kidney disease (OR=1.56, P=8.7x10^-5^, 95% CI 1.25-1.96) and incident circulatory disease (HR=1.17, P=3.6x10^-12^, 95% CI 1.12-1.23), and was significantly associated with other prevalent hypertensive complications (OR=2.80, P=7.9x10^-5^, 95% CI 1.68-4.66). Additionally, each SD thinner PS layer was significantly associated with risk of future circulatory conditions including myocardial infarction (HR 1.17, P=8.1x10^-7^, 95% CI 1.10-1.25), nonhypertensive congestive heart failure (HR 1.25, P=4.9x10^-5^, 95% CI 1.12-1.39), cerebrovascular disease (HR 1.15, P=1.9x10^-5^, 95% CI 1.08-1.23), peripheral vascular disease (HR 1.32, P=4.5x10^-5^, 95% CI 1.16-1.52), and abdominal aortic aneurysms (HR 1.47, P=6.4x10^-6^, 95% CI 1.24-1.74). Other retinal layers where thinning was associated with increased risk of future circulatory disease included GCC for aortic aneurysms and congestive heart failure, RNFL for hypertension, heart failure, cerebrovascular disease, and paroxysmal supraventricular tachycardia, and CSI for ischemic heart disease and hypertrophic cardiomyopathy.

A thinner PS layer was associated with higher odds of having a history of type 1 and type 2 diabetes, and ophthalmic and neurological manifestations of diabetes, with the strongest effect estimates observed for prevalent ophthalmic manifestations of type 1 diabetes (OR=2.93, P=1.3x10^-5^, 95% CI 1.81-4.75), and incident neurological manifestations of type 2 diabetes (HR=1.65, P=4.2x10^-5^, 95% CI 1.30-2.10). Further associations with traits linked to metabolic disease were also detected, including significant associations between thinner PS and elevated glycosylated hemoglobin (HBA1c) (β 0.05, P=1.5x10^-34^, 95% CI 0.05 – 0.06), total body fat percentage (β 0.08, P=5.7x10^-7^, 95% CI 0.08 – 0.07), serum triglyceride level (β 0.06, P=5.3x10^-41^, 95% CI 0.07 – 0.05), and resting heart rate (β 0.10, P=8.2x10^-100^, 95% CI 0.10 – 0.09). Conversely, a relatively thicker PS layer was correlated with higher serum high-density lipoprotein (HDL) (β 0.11, P=5.0x10^-131^, 95% CI 0.10 – 0.12) and apolipoprotein A levels (β 0.10, P=9.4x10^-107^, 95% CI 0.09 – 0.11).

Highly significant (P<5x10^-8^) associations among respiratory conditions were observed among participants with thinning of the PS layer. A thinner PS was associated with higher odds of having a history of asthma (OR=1.26, P=2.5x10^-21^, 95% CI 1.20-1.33), chronic airway obstruction (OR=1.35, P=4.5x10^-9^, 95% CI 1.22-1.49), and pneumonia (OR=1.30, P=3.5x10^-8^, 95% CI 1.18-1.43). Thinner PS was also associated with incident chronic airway obstruction (HR=1.31, P=2.9x10^-23^, 95% CI 1.24 – 1.38), pneumonia (HR=1.22, P=4.8x10^-14^, 95% CI 1.16 – 1.29), chronic bronchitis (HR=1.52, P=5.5x10^-17^, 95% CI 1.38 – 1.67) and emphysema (HR=1.47, P=1.8x10^-10^, 95% CI 1.30 – 1.65). Significant associations were also identified with quantitative pulmonary phenotypes, with relative PS thinning associated with worse pulmonary function test performance, i.e. reduced forced expiratory volume in the first second (FEV1) (β -0.05, P=7.5x10^-46^, 95% CI -0.05 – -0.06) and forced vital capacity (FVC) (β -0.05, P=1.2x10^-46^, 95% CI -0.05– -0.06).

A thinner retinal layer was also strongly associated with neurological conditions. Thinner GCC, RNFL, GCL, and IPL layers were each independently associated with higher odds of having a history of MS and epilepsy, as well as incident MS, with the strongest effect estimate observed for GCL thinning and prevalent MS (OR=2.89, P=1.8x10^-22^, 95% CI 2.34-3.58).

Finally, significant associations were identified between thinner retinal layers and substance use disorders. Thinner inner retinal layer (GCC, RNFL, GCL and IPL) and PS layer were independently associated with a history of alcohol-related disorders, for example, a thinner RNFL was associated with alcoholic liver damage (OR=1.98, P=3.3x10^-5^, 95% CI 1.44-2.74). In keeping with this, a thinner GCC layer was associated with higher serum hepatic enzyme levels, such as gamma-glutamyltransferase (GGT) (ϕ3 0.04, P=8.4x10^-20^, 95% CI 0.03 – 0.05). A thinner inner retinal layer was also associated with both a history of tobacco use disorder and incident tobacco use disorder, with the strongest effect estimate for a thinner RNFL layer and prevalent tobacco use disorder (OR=1.18, P=1.2x10^-5^, 95% CI 1.09-1.27).

Further conditional analyses of the photoreceptor segment prevalent disease and quantitative clinical trait OCT-PheWAS was performed adjusting for combined prevalent and incident T2DM, HbA1c (as a finer marker of glycemic control), HTN, and a more detailed 25-factor smoking covariate as previously used (*50*). We identified that after adjusting for smoking, diabetes, and HTN, initially the conditions with strongest associations with photoreceptor segment thickness, continued significant associations with thinner photoreceptor segments were present for pulmonary (asthma, chronic airway obstruction or COPD), renal (renal dialysis, hypertensive chronic kidney disease, pyelonephritis), and oncologic (chemotherapy and malignancies) phenotypes (**Supplementary Figure 10a**). Additionally, adjustment for smoking, diabetes, and HTN, while attenuating the association for glycemic traits, did not significantly change the other quantitative clinical trait associations (**Supplementary Figure 10b**), which still largely remain significant despite adjustment for these common comorbidities.

### Common variant genome-wide association study and *in silico* analyses

Genome-wide association of the 9 retinal layers identified 259 independent loci **(Figure 4, Supplementary Figure 11,12, Supplementary Table 6-7)**, of which 64 were shared between at least 2 layers. The layer thickness with the highest heritability was the OPL+ONL (h^2^=34.4%, 97 independent loci) followed by INL (h^2^=28.5%, 67 independent loci), IPL (h^2^=25.4%, 56 independent loci, PS (h^2^=24.6%, 61 independent loci), RNFL (h^2^=23.3%, 43 independent loci), GCC (h^2^=23.1%, 38 independent loci), GCL (h^2^=21.9%, 40 independent loci), CSI (h^2^=16.8%, 18 independent loci), and RPE+BM (h^2^=11.1%, 21 independent loci) (**Figure 4b**). Among the genome-wide significant independent loci, the variants with strongest association (P<1x10^-50^) are highlighted in **Figure 4c**. Notably, when searching these variants across the phenoscanner portal, rs62075723-G (allele frequency 36%), an intron variant in *TSPAN10* linked to the *ACTG1* gene by PoPS, which is associated with thickening of the GCL layer (beta=0.13 SD per A allele, P=3.93x10^-63^) and thinning of the PS layer (beta=-0.07 per A allele, P=9.53x10^-20^), is also associated with increased risk of AMD (OR 1.12, 95% CI: 1.09-1.16,P=2.5x10^-11^). Furthermore, rs17421627-G, a *LINC00461* intron variant linked to *MEF2C* by PoPS, which is associated with increased thickness of the INL layer (Beta 0.41SD, P=7.5x10^-193^) has also been associated with a 3um increase in retinal vascular caliber (P=7.0x10^-16^)(*51*). Moreover, the *CFH*, *HTRA1/ARMS2* loci previously linked to AMD were significantly associated with PS, RPE+BM thickness. Similarly, top loci in glaucoma GWAS, *THRB*, *RARB*, and *ACOXL/BCL2L11*, *STOX2/WWC*2 were significantly associated with inner retinal layer thickness(*52*). Lastly loci related to cataract development: *BMP3*, *OCA2*, *CASZ1*, *QKI*, and *HMGA2*, were significantly associated with inner layer thickness(*53*).

**Figure 4:**
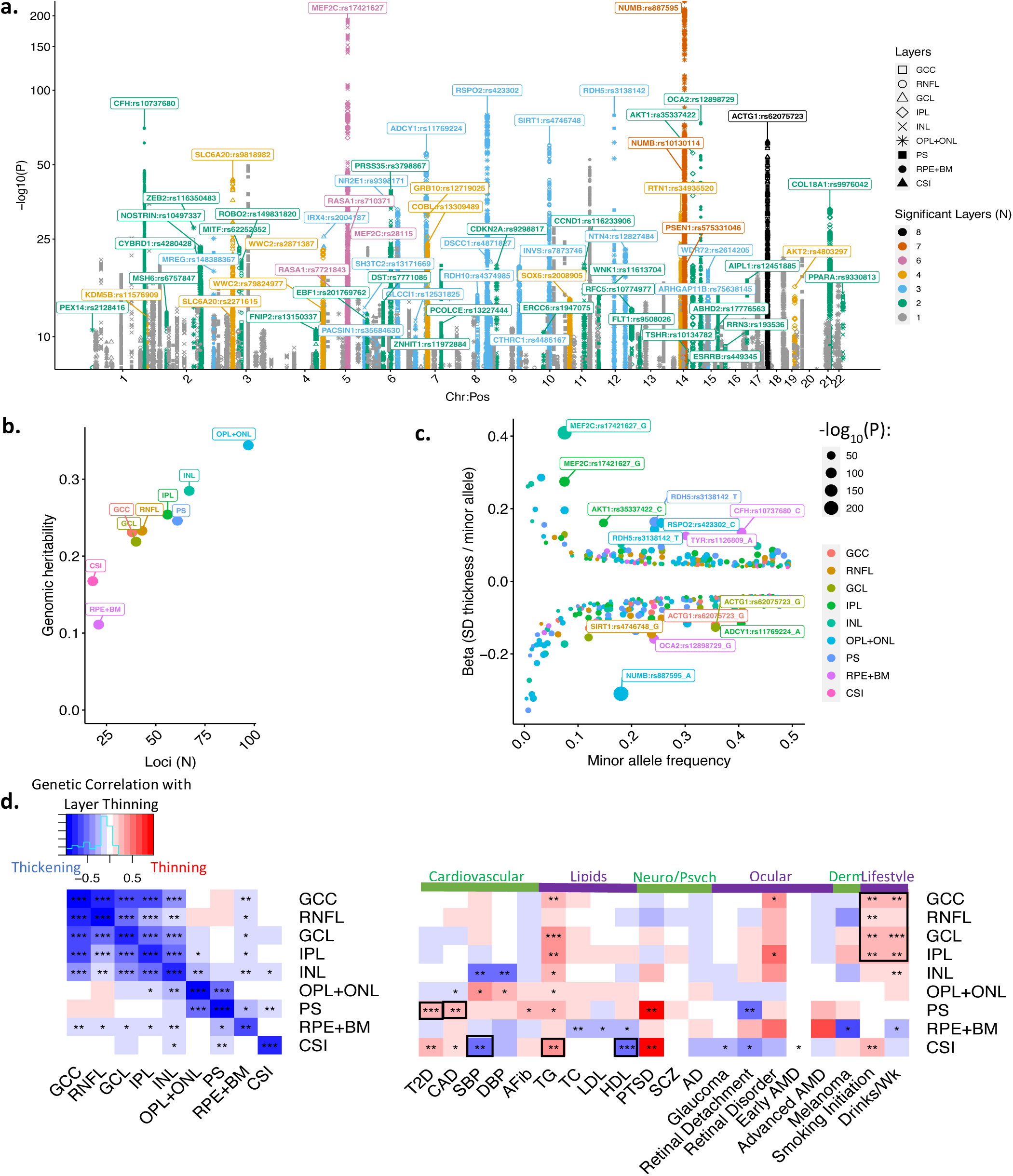
Genome-wide association study of retinal layer thickness. a. Manhattan plot of genome-wide significant variants (P<5x10^-8^) with retinal layer thickness across 9 retinal layers. Analyses are adjusted for age, age^2^, sex, spherical equivalent, genetic ancestry, and genotyping array. Individual dots reflect genomic variants which are colored by the number of layers significant for the variant, and shape reflects the layer with the most significant association for that variant. Labeled loci are genome-wide significant in at least 2 layers and are labeled by the top predicted PoPS gene within a 1MB window of the variant. Full results are available in Supplementary Tables 6-7. b. Predicted genomic heritability by layer plotted against the number of genome-wide significant, independent loci (LD clumping with r^2< 0.1) for that layer. c. plot of the minor allele frequency versus effect size beta across all 259 independent, top genome-wide significant variants. Variants with P<1x10-50 are labeled with the gene and variant as well as the minor allele. d. genomic correlation analyses across the 9 layers (left) as well as several available GWAS traits previously published (right). Within cells, association strength is denoted through the corrected P-value as such: ***P<1x10-4, **1x10-4≤P<0.01, *0.01≤P<0.05. The color reflects the corrected genomic correlation for association, where red reflects layer thinning and blue reflects thickening. Full quantitative results for the genomic correlation analyses are provided in Supplementary Table 12. GCC = ganglion cell complex or RNFL+GCL+IPL, RNFL = retinal nerve fiber layer, GCL = ganglion cell layer, IPL = inner plexiform layer, INL = inner nuclear layer, OPL+ONL = outer plexiform layer and outer nuclear layer, PS = photoreceptor segment, RPE+BM = retinal pigment epithelium and Bruch’s membrane complex, CSI = choroid layer.

Replication of the 259 independent genome-wide significant associations from the UK Biobank analyses was performed using the LIFE-Adult-Study(*46*), a population of 6,313 individuals of central European descent from Germany with OCT imaging and layer segmentation performed similarly to the UK Biobank as well as genotyping (mean age 57.6 years (SD 12.5), with 3098 Males, 3215 Females)^46,53^ (**Supplementary Table 8**)(*46, 54*). Significant positive correlation (R>0.8) was identified between the betas of independent genome-wide significant loci in the UK Biobank and those in the LIFE-Adult-Study replicating 154/234 (66%) of top independent UK Biobank variants with nominal significance (P<0.05) across the top 8 layers of the retina (**Supplementary Figure 16, Supplementary Table 9-10**).

To hypothesize possible genes implicated by each locus, we performed gene prioritization using PoPS (**Supplementary Table 11, Supplementary Figures 17-20**). The top prioritized gene at the only locus significantly associated with 8 retinal layers (all layers except CSI) was *ACTG1*, with the top variant being an intron variant in *TSPAN10*: rs62075723-A, wherein individuals with the alternative A allele (Allele frequency 64%) had thicker retinal layers. Four identified top genome-wide significant loci were coding missense variants predicted to be damaging by PolyPhen(*46, 55*) and deleterious by SIFT(*56*): 1) *TRPM1*:rs75638145-A which was significantly associated with thinning of the INL, IPL, and GCL layers, 2) *RD3L*:rs35337422-C which was significantly associated with IPL and INL thickening, 3) *TYR*: rs1126809-A which was significantly associated with thickening of specifically the RPE, and 4) *RAX2*:rs76076446-A which was significantly associated with thinning of the OPL layer (**Supplementary Table 7,11**).

After performing GWAS and implementing gene prioritization, we performed analyses to prioritize the biological pathways and traits linked to the identified GWAS loci. We performed biological pathway enrichment analyses, using the FUMA and EnrichR web browsers for genes with a PoPS z-score>1. Prioritized genes were enriched for pathways linked to apoptosis and cell death, cellular senescence (telomere maintenance, DNA damage checkpoints and repair, TP53 signaling), the innate immune system (e.g. interleukin, C3/C5 signaling), angiogenesis (e.g. VEGF, NOTCH, WNT and Hedgehog signaling pathways), melanin biosynthesis, and pathways related to neural processes (axon and neuron growth, phototransduction, FGFR signaling) (**Supplementary Figure 21-23**). Genetic correlation analysis was subsequently performed using GNOVA(*57*), using GWAS summary statistics across multiple ocular and systemic diseases. Firstly, there was significant genetic correlation identified between layers of the retina, most notably between the inner layers (GCC through INL), and also between the outer layers (OPL+ONL and PS, CSI and PS). Several significant associations were also identified between inner and outer layer thicknesses, most notably between the INL and OPL+ONL, and between RPE+BM and INL and GCC layers. Significant (P<0.01) associations concordant with the phenotypic associations were identified for cardiovascular phenotypes type 2 diabetes mellitus and coronary artery disease with PS thinning, elevated systolic BP and HDL with CSI thickening, elevated TG with CSI thinning, and smoking and alcohol use with thinning of the inner retinal layers, particularly the GCL and IPL (**Figure 4d**).

### Polygenic Risk Score Phenome-wide Association Study (PRS-PheWAS)

Mendelian randomization uses human genetics for causal inference by leveraging the random assortment of genetic variants during meiosis at conception, which diminishes susceptibility to confounding or reverse causality(*58*). Here, polygenic risk score phenome-wide association study (PRS-PheWAS) was performed to assess both the putative causal relationships between 1) systemic phenotypes and retinal layer thicknesses, and 2) retinal layer thicknesses and systemic and ocular disease.

First, additive polygenic risk scores (PRS) were created for each retinal layer thickness using independent, genome-wide significant variants for each specific layer. The retinal layer thickness PRS were strongly associated with their respective retinal layers in the UK Biobank (Supplementary Table 15a). Sensitivity analyses identified no significant association between the retinal layer thickness PRS and possible environmental confounders (ex: socioeconomic status as indicated through Townsend deprivation index, alcohol intake, diet, stress, exercise), suggesting further robustness of our retinal OCT PRS to possible confounders (Supplementary Table 15b). Association of OCT layer thickness PRS with ocular and systemic conditions were performed, adjusting for different covariates (**Figure 5**, **Supplementary Figure 24, Supplementary Table 14**). Several consistent associations between genetic and epidemiological retinal layer thicknesses and common ophthalmic conditions were observed (**Figure 5a,c, Supplementary Figure 25**). Notably, several expected positive control associations were identified, namely: both epidemiological and genetic thinning of the PS layer were associated with increased incidence and prevalence of AMD (**Supplementary Figure 25**). Moreover, both epidemiological and genetic thinning of the RNFL layer were associated with increased incidence and prevalence of Glaucoma (**Supplementary Figure 25**). A novel association was identified between epidemiological and genetic thinning of the inner layers of the retina (RNFL, GCL, IPL) and increased risk of incident and prevalent cataracts (**Supplementary Figure 25**). Several significant, concordant associations were also identified between epidemiologic and genetic systemic traits with retinal layer thickness (**Figure 5a,b**, **Supplementary Figure 26, Supplementary Table 15-16**). In particular, concordant genetic and epidemiological associations were identified between several ocular measures and their effect on retinal layer thicknesses, including: 1) increased intraocular pressure and thinning of the inner retinal layers (RNFL,GCL, IPL, INL), and 2) decreased spherical equivalent (myopia) and thinning of all retinal layers except RPE+BM and RNFL layers (Supplementary Figure 26). Furthermore, significant concordant genetic and epidemiological associations were identified between cardiopulmonary factors (increased percent body fat, BMI, heart rate, reduced performance on pulmonary function tests) and thinning of the PS. Furthermore, elevated systolic blood pressure had significant genetic and epidemiological associations with thinning of the inner retinal layers (GCC and RNFL) (**Supplementary Figure 26**). Other concordant significant associations were also identified between quantitative genetic systemic phenotypes and their respective epidemiological phenotypes with retinal layer thickness, including between reduced left ventricular ejection fraction and thinning of the PS, and reduced kidney function (increased creatinine) and thinning of the INL, PS, and CSI layers.

**Figure 5:**
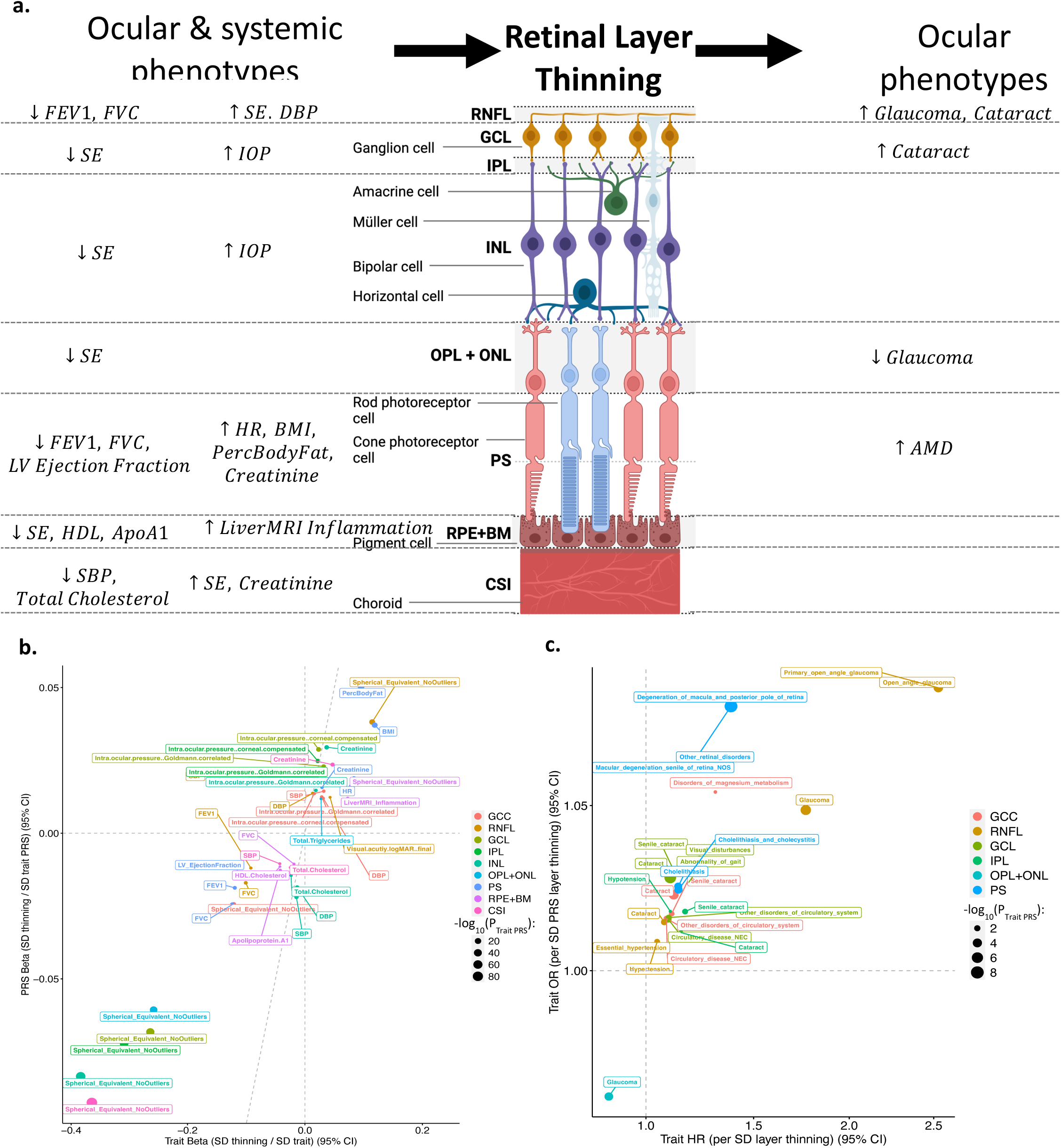
Concordant epidemiological and genetic associations with retinal layer thickness. a. Schematic of concordant epidemiological and genetic associations with retinal layer thickness visually showing ocular and systemic phenotypes that may causally influence retinal layer thinning, and the effect of retinal layer thinning on incident ocular phenotypes. b. Plot of quantitative phenotypes which are significantly and directionally concordant in their association with retinal layer thickness both epidemiologically (x-axis) and genomically between polygenic risk scores for the the quantitative phenotypes and retinal layer thickness (y-axis). In both models, analyses are adjusted for age, age^2^, sex, spherical equivalent, genetic ancestry, and genotyping array. Full results comparing epidemiological and genetic associations between all quantitative traits and retinal layers are provided in Supplementary Table 16. c. Plot of retinal layers which are significantly and directionally concordant in their association with diseases both epidemiologically with incident disease (x-axis) and genomically between polygenic risk scores for the retinal layer and any prevalent or incident disease (y-axis). In both models, analyses are adjusted for age, age^2^, sex, spherical equivalent, genetic ancestry, and genotyping array. Full results of the retinal layer polygenic risk score associations with all analyzed phenotypes are provided in Supplementary Table 15. GCC = ganglion cell complex or RNFL+GCL+IPL, RNFL = retinal nerve fiber layer, GCL = ganglion cell layer, IPL = inner plexiform layer, INL = inner nuclear layer, OPL+ONL = outer plexiform layer and outer nuclear layer, PS = photoreceptor segment, RPE+BM = retinal pigment epithelium and Bruch’s membrane complex, CSI = choroid layer.

### Rare variant association study

Lastly, rare variant association analysis was performed in order to analyze rare disruptive variants grouped by gene to improve power of association of this class of variants not included in the aforementioned genome-wide association analysis. 11 significant associations were identified, after adjusting for age, age^2^, sex, spherical equivalent, genetic ancestry, and genotyping array (**Figure 6a**, **Supplementary Table 17**). The strongest association identified was for carriers of rare *IMPG2* deleterious variants who had on average 3um thinner PS layer (Beta -1.19 SD, P=1.13x10^-9^) (**Figure 6b**). Of note, IMPG2 gene encodes for the interphotoreceptor matrix proteoglycan 2 protein known to play a role in the organization of the interphotoreceptor matrix, defects of which are a cause of retinitis pigmentosa type 56 and IMPG2-related maculopathy(*59–61*). This gene was also a significant locus identified in the common variant genome-wide association analyses for PS thickness, further highlighting its important role both across common and rare genomic variants, in PS thickness.

**Figure 6:**
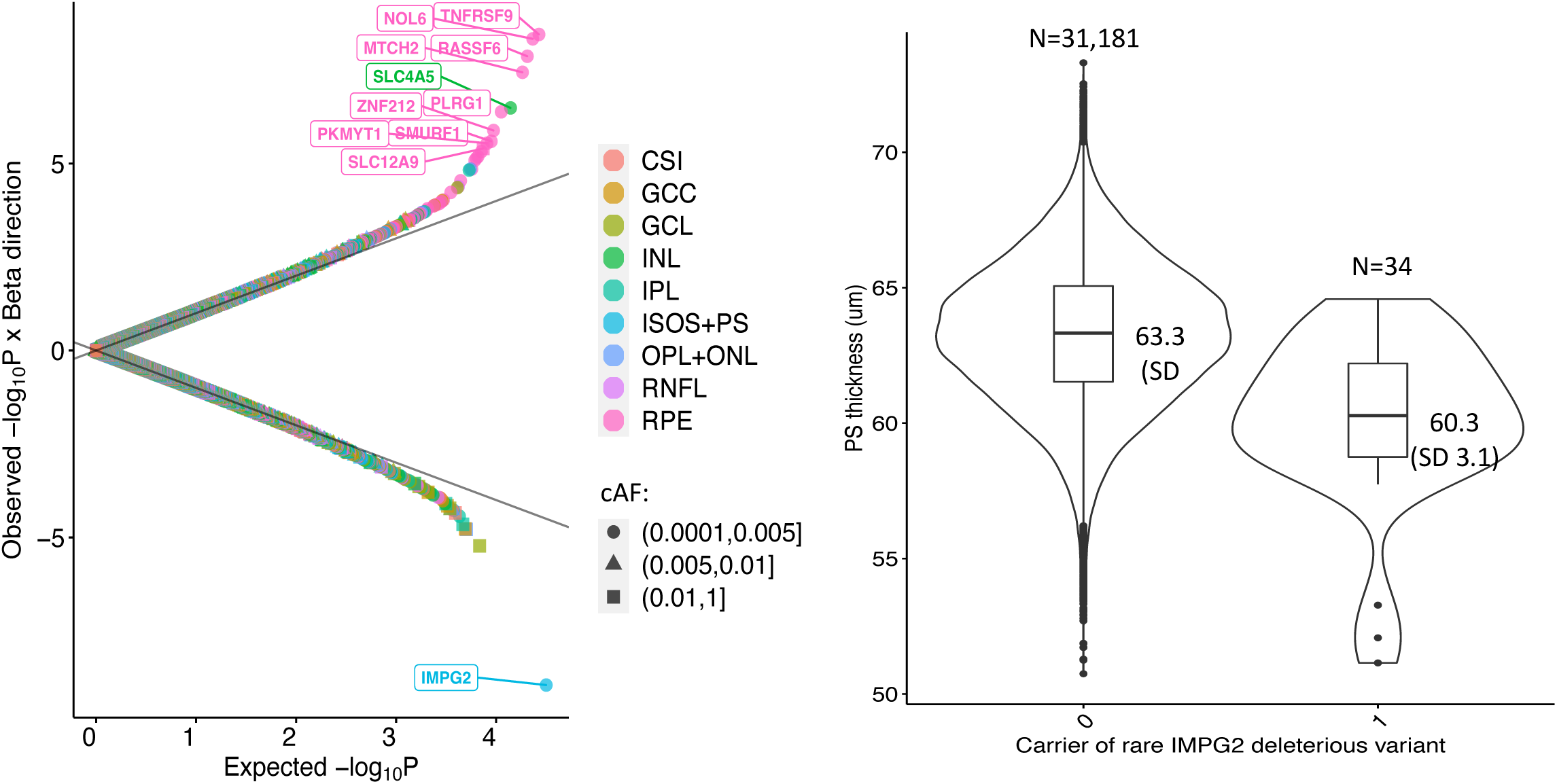
Rare variant association study of retinal layer thickness. a. Quantile-quantile plot of rare disruptive variants grouped by gene with retinal layer thickness across 9 retinal layers. Analyses are adjusted for age, age^2^, sex, spherical equivalent, genetic ancestry, and genotyping array. Labeled dots reflect genome-wide significant genes (P<0.05/11,767=4.2x10^-6^) which are colored by the respective layer. Full results are available in Supplementary Table 17. b. For the strongest association, of rare disruptive variants in IMPG2 with photoreceptor segment (PS) thickness, violin plots showing that the mean PS thickness in non-carriers (0) is 63.3 (SD 2.7) um versus carriers of a disruptive variant in IMPG2 (1) is 60.3 (SD 3.1) um. GCC = ganglion cell complex or RNFL+GCL+IPL, RNFL = retinal nerve fiber layer, GCL = ganglion cell layer, IPL = inner plexiform layer, INL = inner nuclear layer, OPL+ONL = outer plexiform layer and outer nuclear layer, PS = photoreceptor segment, RPE+BM = retinal pigment epithelium and Bruch’s membrane complex, CSI = choroid layer.

## Discussion

We present the findings of in-depth, unbiased phenome-wide and genome-wide assessments using retinal OCT images from 44,823 individuals. We identified systemic and ocular phenotypes associated with retinal layer thicknesses through phenome-wide analyses. Genome-wide association of the 9 retinal layer thicknesses yielded 259 loci, and burden analyses of rare (allele frequency <1%) disruptive coding alleles identified 11 genes influencing retinal layer thickness. Comparative epidemiological and genetic associations identified multiple ocular and systemic traits linked to retinal layer thinning, and also hypothesize retinal layers wherein thinning may predispose to common ocular conditions. Together, these results permit several conclusions.

First, phenome-wide association analysis identified that retinal layer thicknesses measured on OCT images are significantly linked to diverse conditions, providing new insights on the retina as a biomarker for systemic disease risk and severity. In particular, thinner GCC and PS layers were both associated with higher risk of incident mortality, independent of age, sex, blood pressure, HbA1c, height, weight, and coronary artery disease. This suggests that retinal inner layer and PS thinning are associated with mortality, overall health, and frailty among those with existing cardiometabolic disease. To the best of our knowledge, our work represents the first discovered association between retinal layer thinning and incident mortality. A recent paper describing predicted age derived from retinal fundus images shows that the ‘retinal age gap’ (retinal age – chronological age) is strongly correlated with all-cause mortality, thus implying the clinical significance of retinal age as a biomarker of systemic health(*62*). We further explored the conditions contributing to incident mortality through phenome-wide association, and observed significant associations between retinal layer thinning and higher prevalence of, and separately higher risk for, ocular (glaucoma, AMD, cataract), neuropsychiatric (MS, epilepsy, schizophrenia, mood disorders), cardiometabolic (hypertension, diabetes, atherosclerosis, heart failure), and end-organ failure (renal failure, liver cirrhosis, heart failure, respiratory failure). This suggests that retinal layer thinning may signify more widespread cardio-pulmonary-renal and neurologic disease, and may be used as an ophthalmic biomarker for risk of future systemic conditions.

Second, we identify concordant epidemiological and genetic associations representing a likely causal link between thinner layers and ocular conditions: thinner RNFL and thicker OPL+ONL with glaucoma, thinner RNFL and GCL with cataracts, and thinner PS with AMD. Of note, prior work in the UK Biobank has shown that a genetic risk score for GCC thickness is not associated with glaucoma, concordant with our findings(*63*). Here, we show that further differentiation of the layers making up the GCC identifies epidemiologic and genetic thinning of the RNFL layer as being specifically associated with glaucoma development. While the role of OPL+ONL thickening in glaucoma progression is unclear, it is possible that it is a compensatory response to RNFL thinning in glaucoma development. Furthermore, PS thinning precedes AMD diagnosis by decades epidemiologically, with individuals genetically predisposed to PS thinning having increased risk of AMD compared to others. This has been further detailed in our prior work(*64*), where we also show an AMD polygenic risk score to be associated with PS thinning, suggesting a bidirectional relationship between PS thinning and AMD. Lastly, the link between a thinner RNFL and GCL and cataract development is new and has not been previously described. Further understanding of the roles of the retinal inner layer health on the health of the vitreous fluid and lens may further enable understanding of this association.

Third, our study highlights the link between cardiometabolic, pulmonary, and renal conditions with a thinner PS layer. We show consistent epidemiological and genetic associations, suggestive of causal associations, linking poor cardiac, metabolic, pulmonary, and renal function with thinner PS. Firstly, polygenic risk score analyses identified that individuals born with genetically elevated risk for elevated cardiovascular (heart rate, blood pressure, left ventricular ejection fraction), metabolic (BMI, percent body fat), pulmonary (lower FEV1), and renal (creatinine) phenotypes have thinner photoreceptor segment layer (as measured at the baseline UKBB visit). Secondly, we have shown that phenotypically, individuals with a history of cardiometabolic, renal, and pulmonary disease have thinner retinal photoreceptor segment layers on imaging at their baseline UKBB assessment as seen in the prevalent disease analysis. Thirdly, cross-sectionally individuals with higher cardiovascular (heart rate, blood pressure, left ventricular ejection fraction), metabolic (BMI, percent body fat), pulmonary (FEV1), and renal (creatinine) phenotypes have thinner photoreceptor segment layers as well (all as measured at the baseline UK Biobank visit). The incident analyses also corroborated these findings showing that a thinner photoreceptor segment layer is associated with future incident cardiometabolic, renal, and pulmonary disease, suggesting that individuals with thinner photoreceptor segment layers already have some existing undiagnosed pre-clinical cardiometabolic or pulmonary disease (ex: pre-diabetes, metabolic disease) which makes them at higher risk for becoming diagnosed with incident disease. Overall, our study provides evidence in support of the association between cardiopulmonary and renal disease and photoreceptor layer thickness which have the potential to augment existing clinical risk prediction methods. This information may also be used towards prevention in individuals at risk for photoreceptor degeneration, such as those at risk for macular degeneration. Further studies are needed to determine whether improved cardiometabolic, pulmonary, and renal status – ex: via changes in lifestyle, diet, environment, or use of risk-lowering medications, may dampen the rate of PS thinning with age.

From a cardiometabolic perspective, while a large body of evidence supports the value of retinal microvasculature metrics from fundus photography (*27, 30, 71–73*) in determining cardiometabolic risk, few publications examine the associations between different layers from retinal OCT data and cardiovascular events and/or risk factors. A recent study in a small cohort identified that subretinal drusenoid deposits, one of the pathways to advanced AMD, are associated with cardiovascular disease and stroke(*74*). Several other studies have similarly identified associations between AMD and risk of heart failure(*75*) and cardiometabolic disease(*76*). Moreover, the limited number of small studies on the association between OCT parameters and cardiovascular disease report contradictory results. For example, one prospective case-control study found no differences in chorioretinal thickness measurements between patients with hypertension and matched controls(*65*). However, subsequent studies have shown choroidal thinning(*77, 78*) and retinal thinning in hypertensive subjects(*78, 79*). A more recent study concludes that increased subfoveal choroidal thickness was associated with hypertensive retinopathy but not the presence of systemic hypertension in the absence of ocular manifestations(*80*). In our study, we clearly show consistent epidemiologic and genetic relationship between hypotension and choroidal thinning. Overall, the reported associations between OCT parameters and hypertension are conflicting and interpreting the results together is hindered by varied cohort demographics, the use of different imaging devices, and inconsistently reported comorbidities and medication use. Given the limitations of the small studies published to date and the inconsistency of reported results, our study in a large population cohort represents meaningful progress in establishing the significance of OCT-derived thickness measurements in relation to cardiovascular risk factors and disease.

Moreover, from a renal perspective, we identify a new and significant genetic and epidemiologic association between poor renal function and a thinner PS layer. Retinal thinning in the context of early CKD is supported by previous work demonstrating that patients with non-diabetic predialysis CKD have approximately 5% retinal thinning compared to both healthy matched controls and patients with hypertension(*65*). Further small clinical OCT studies corroborate the finding of retinal and choroidal thinning in patients on hemodialysis(*66–69*). The eye and kidney are homologous developmentally and structurally, with shared microcirculatory characteristics and basement membrane constituents(*70*), which provides rationale for the utility of ocular imaging in signalling renal pathology.

Fourth, a significant likely causal link between poor pulmonary function and thinner RNFL as well as PS layers was identified. Having a history of asthma, chronic airway obstruction, and pneumonia was associated with a thinner PS layer, which was also associated with incident chronic airway obstruction, pneumonia, chronic bronchitis, and emphysema. In keeping with these findings, significant associations were also observed with quantitative pulmonary phenotypes, with poor performance on pulmonary function tests (i.e. lower FEV1 and FVC) being associated with thinner RNFL and PS layers. Corroboratory genetic evidence for this was also observed, with genetic predisposition to lower FEV1 and FVC being associated with thinner RNFL and PS layers. Thinning of the RNFL and choroid have been associated with COPD in a number of cross-sectional studies(*81–85*). OCT studies in the context of infectious pneumonia are few in number and mostly relate to SARS-CoV-2 (COVID-19), which is variably reported to be associated with reduced macular thickness and RNFL thinning(*86*), as well as increased RNFL and GCC thickness compared to controls(*87*). A further similar study showed no significant OCT changes in patients diagnosed with COVID-19(*88*), and several authors urge caution in the interpretation of OCT findings that may represent variants of normal or imaging artefact(*89, 90*). In the presence of non-infectious idiopathic interstitial pneumonia, a single paper demonstrates significant RNFL thinning(*91*). The relative paucity and, in some cases, contradictory nature of publications in this field emphasizes the need for ongoing appropriately powered studies to find evidence for the clinical utility of retinal imaging in the context of respiratory disease. Here, we clearly show evidence that both epidemiologically and genetically lower FEV1 and FVC are associated with thinning of the RNFL and PS layers, and provide further evidence from phenome-wide association across prevalent and incident traits which further substantiates the link between RNFL and PS thinning and poor pulmonary function.

Fifth, our phenome-wide association analyses highlight the utility of retinal OCT imaging for risk prediction of future neurological conditions. Ocular imaging is intuitively relevant for examining axonal degeneration as unmyelinated axons that synapse directly within the CNS lie in the RNFL. Indeed, our results demonstrate that thinning of the inner retinal layers (GCC, RNFL, GCL, and IPL) were each independently associated with higher odds of having a history of multiple sclerosis and epilepsy, as well as risk of future multiple sclerosis. Pathological inner retinal changes have previously been linked with a range of neurodegenerative diseases, and retinal OCT measurements thus provide potential surrogate markers of disease states. Many previous studies also demonstrate RNFL and GCL thinning in patients with multiple sclerosis, both with and without a history of optic neuritis(*92, 93*). Similarly, the association between inner retinal thinning and epilepsy has also been corroborated in previous small studies(*94, 95*). Retinal OCT assessment has the potential to determine the presence and severity of atrophic cerebral changes previously only diagnosed and monitored using magnetic resonance imaging (MRI) brain scanning(*96*). OCT offers significant advantages over MRI in terms of availability, patient acceptability, clinical resource allocation, and the capability to precisely quantify neuronal tissue loss. Importantly, retinal OCT, in addition to known clinical and imaging factors, may facilitate the diagnosis of multiple sclerosis earlier than reliance on MRI alone(*96*), and may allow more patients to benefit from reduced secondary disability and improved life expectancy with the timely initiation of disease-modifying treatments(*97*).

Sixth, among substance use conditions, our results also show significant independent associations between PS and inner retinal layer thinning (GCC, RNFL, GCL and IPL) and a history of alcohol-related disorders and relevant traits, including alcoholic liver damage and increased serum hepatic enzyme levels, for example, GGT. Retinal layer thinning was also associated with both a history of tobacco use disorder and incident tobacco use disorder, with the strongest effect estimate observed for RNFL thinning and prevalent tobacco use disorder. Several studies of limited sample size unanimously conclude that alcohol use is significantly associated with reduced retinal thickness(*98–100*). A recent meta-analysis found no significant differences in overall retinal and choroidal thicknesses between tobacco smokers and non-smokers, but found that smoking was associated with a thinner RNFL and GCL(*101*). Retinal OCT imaging may, therefore, be valuable in corroborating the presence and chronicity of tobacco and alcohol misuse, and may potentially provide information on adherence to recommended cessation if supported by longitudinal studies.

Seventh, genome-wide association of retinal layer thicknesses yielded 259 loci, enriched in pathways linked to apoptosis and cell death, cellular senescence (telomer maintenance, DNA damage checkpoints and repair, TP53 signaling), the innate immune system (e.g. interleukin, C3/C5 signaling), angiogenesis (e.g. VEGF, NOTCH, WNT and Hedgehog signaling pathways), melanin biosynthesis, and pathways related to neural processes (axon and neuron growth, phototransduction, FGFR signalling). We have developed a user-interface for all of the retinal layer thickness GWAS data and further downstream analyses that other users may do on a webplatform called the "Ocular knowledge portal" detailed here: https://ocular.hugeamp.org/dinspector.html?dataset=Zekavat2021_RetinalLayerThickness_EU. Moreover, burden analyses of rare (allele frequency <1%) disruptive coding alleles identified 11 influential genes. Furthermore, genetic correlations were observed between retinal layer thinning and systemic traits including diabetes, coronary artery disease, hypertension, hyperlipidemia and neuropsychiatric traits. Further dissection of the identified genetic loci, and overlap with single-cell RNA sequencing across different disease states will enable confirmation of layer-specific transcripts and proteins contributing towards retinal layer thickness in different conditions. Together, genetic loci linked to retinal layer thicknesses may enable better understanding of mechanistic pathways linked to ocular conditions. For example, multiple loci linked to glaucoma, AMD, and cataracts are present among the retinal layer thickness GWAS analyses. The *CFH*, *HTRA1/ARMS2* loci previously linked to AMD were significantly associated with PS, RPE+BM thickness. Similarly, top loci in glaucoma GWAS, *THRB*, *RARB*, and *ACOXL/BCL2L11*, *STOX2/WWC*2 were significantly associated with retinal inner layer thickness(*52*). Lastly loci related to cataract development, *BMP3*, *OCA2*, *CASZ1*, *QKI*, and *HMGA2*, were significantly associated with inner layer thickness(*53*). Thus, knowledge of overlaps between retinal layer thickness GWAS and ocular GWAS may lead to insights on pathophysiology of these genetic loci on ocular conditions. Similarly, knowledge of other biological pathways also linked to retinal layer thickness may lead to potential therapeutic targets for ocular pathologies.

Overall, our study aligns well with the shared aims of a number of longitudinal studies internationally; to identify biomarkers that allow risk stratification and provide surrogate end points to enable novel means of disease surveillance as well as catalyze the discovery of new treatments. The strengths of our study include the large sample size, standardized data collection protocols, data validity, and statistical adjustment for many confounding factors in our models. However, we acknowledge that our study is limited by the sole use of UK Biobank data derived from voluntary participants, including a limited ethnic diversity among a cohort of European ancestry. As more diverse cohorts become available with retinal imaging and genomics data, it will become imperative to incorporate them into similar analyses to enable discovery more broadly applicable across different ancestries. Furthermore, we note that UK Biobank is known to be a healthier sample compared to the general population(*102*); while this may influence the baseline statistics of retinal layer thicknesses compared to the general population, it is unlikely to affect exposure-disease relationships which are more generalizable. The presence of external validation of our results using the LIFE-Adult-Study provides further confidence in our association analyses. However, despite adjustment for known confounding factors, it is not possible to exclude the presence of residual, unknown and/or unmeasured confounders.

For many conditions, currently used clinical methods are lacking with respect to risk stratification and there is a pressing need to define novel means of identifying at-risk patients, specifically before the onset of potentially irreversible end-organ damage. To help fulfil this unmet need, our results add to existing evidence to support the use of retinal OCT imaging parameters as biomarkers for systemic and ocular diseases. Future research is warranted to determine and quantify the value of retinal OCT measurements used independently or in combination with existing risk assessment methods. Furthermore, our study highlights how retinal imaging may be integrated with electronic health records, genomic data, and other biomarkers to advance our understanding of disease mechanisms and help inform risk prediction and risk modification strategies.

## Data Availability

We have developed a user-interface for all of the retinal layer thickness GWAS data and further downstream analyses that other users can do on a webplatform called the "Ocular knowledge portal" detailed here: https://ocular.hugeamp.org/dinspector.html?dataset=Zekavat2021_RetinalLayerThickness_EU.

## Acknowledgements

The authors wish to thank the UK Biobank and LIFE-Adult-Study participants for their time and furthermore we gratefully acknowledge Dr. Kerstin Wirkner and the LIFE-Adult-Study team for their commitment to the eye investigation and corresponding exams to make this analysis possible.

## Funding

N.Z. is supported by the National Eye Institute (NEI) 1K23EY032634. P.N. is supported by a Hassenfeld Scholar Award from the Massachusetts General Hospital, and grants from the National Heart, Lung, and Blood Institute (R01HL1427, R01HL148565, and R01HL148050). S.M.Z. is supported by the Leducq Early Career Investigator Award and the NIH National Heart, Lung, and Blood Institute (1F30HL149180-01). J.L.W. is supported in part by NEI (R01EY020928, R01EY022305, R01EY031820, R01EY032559). T. E. is supported by grants from NIH (NIH R21 EY030631, NIH R01 EY030575, NIH P30 EY003790). This research was supported by LIFE Leipzig Research Center for Civilization Diseases, Leipzig University (LIFE is funded by the EU, the European Social Fund, the European Regional Development Fund, and Free State Saxony’s excellence initiative; project numbers: 713-241202, 14505/2470, 14575/2470). F.G.R. was supported by the German Research Foundation (grant number DFG 497989466). The opinions expressed by the authors are their own and this material should not be interpreted as representing the official viewpoint of the National Institutes of Health or National Eye Institute.

## Notes

### Competing Interest Statement

The authors have declared no competing interest.

### Author Declarations

UK Biobank data use was approved by the Massachusetts General Hospital Institutional Review Board (protocol 2021P002040) and facilitated through UK Biobank Applications 7089 and 50211. The LIFE-Adult Study replication was approved by the Ethics Committee of the Medical Faculty of Leipzig University, and the research was conducted in accordance with the Declaration of Helsinki.

## References

1. J. Marshall, The ageing retina: Physiology or pathology. Eye (Basingstoke) 1 (1987), doi:10.1038/eye.1987.47.

2. Y. Solberg, M. Rosner, M. Belkin, The association between cigarette smoking and ocular diseases. [Review] [162 refs]. Surv Ophthalmol 42 (1998).

3. U. Schmidt-Erfurth, Nutrition and retina.Dev Ophthalmol 38 (2005), doi:10.1159/000082772.

4. X. Corso-Díaz, C. Jaeger, V. Chaitankar, A. Swaroop, Epigenetic control of gene regulation during development and disease: A view from the retina*Prog Retin Eye Res* 65 (2018), doi:10.1016/j.preteyeres.2018.03.002.

5. J. D. Unterlauft, M. Rehak, M. R. R. Böhm, F. G. Rauscher, Analyzing the impact of glaucoma on the macular architecture using spectral-domain optical coherence tomography. PLoS One 13 (2018), doi:10.1371/journal.pone.0209610.

6. M. Fleckenstein, T. D. L. Keenan, R. H. Guymer, U. Chakravarthy, S. Schmitz-Valckenberg, C. C. Klaver, W. T. Wong, E. Y. Chew, Age-related macular degeneration. Nat Rev Dis Primers 7 (2021), doi:10.1038/s41572-021-00265-2.

7. C. Y. Cheung, M. K. Ikram, R. Klein, T. Y. Wong, The clinical implications of recent studies on the structure and function of the retinal microvasculature in diabetes*Diabetologia* 58 (2015), doi:10.1007/s00125-015-3511-1.

8. C. Y. L. Cheung, M. K. Ikram, C. Sabanayagam, T. Y. Wong, Retinal microvasculature as a model to study the manifestations of hypertension Hypertension 60 (2012), doi:10.1161/HYPERTENSIONAHA.111.189142.

9. R. Allon, M. Aronov, M. Belkin, E. Maor, M. Shechter, I. D. Fabian, Retinal Microvascular Signs as Screening and Prognostic Factors for Cardiac Disease: A Systematic Review of Current Evidence American Journal of Medicine 134 (2021), doi:10.1016/j.amjmed.2020.07.013.

10. S. M. Heringa, W. H. Bouvy, E. Van Den Berg, A. C. Moll, L. Jaap Kappelle, G. Jan Biessels, Associations between retinal microvascular changes and dementia, cognitive functioning, and brain imaging abnormalities: A systematic review Journal of Cerebral Blood Flow and Metabolism 33 (2013), doi:10.1038/jcbfm.2013.58.

11. C. Y. Cheung, V. T. T. Chan, V. C. Mok, C. Chen, T. Y. Wong, Potential retinal biomarkers for dementia: What is new? Curr Opin Neurol 32 (2019), doi:10.1097/WCO.0000000000000645.

12. M. L. Baker, P. J. Hand, J. J. Wang, T. Y. Wong, Retinal signs and stroke: Revisiting the link between the eye and brain Stroke 39 (2008), doi:10.1161/STROKEAHA.107.496091.

13. G. Cennamo, M. R. Romano, E. C. Vecchio, C. Minervino, C. Della Guardia, N. Velotti, A. Carotenuto, S. Montella, G. Orefice, G. Cennamo, Anatomical and functional retinal changes in multiple sclerosis. Eye (Basingstoke) 30 (2016), doi:10.1038/eye.2015.256.

14. N. Lewczuk, A. Zdebik, J. Bogusławska, A. Turno-Kręcicka, M. Misiuk-Hojło, Ocular manifestations of pulmonary hypertension Surv Ophthalmol 64 (2019), doi:10.1016/j.survophthal.2019.02.009.

15. M. Santos, R. J. Hofmann, Ocular manifestations of obstructive sleep apnea Journal of Clinical Sleep Medicine 13 (2017), doi:10.5664/jcsm.6812.

16. M. Mentek, F. Aptel, D. Godin-Ribuot, R. Tamisier, J. L. Pepin, C. Chiquet, Diseases of the retina and the optic nerve associated with obstructive sleep apnea Sleep Med Rev 38 (2018), doi:10.1016/j.smrv.2017.05.003.

17. A. W. Vaes, M. A. Spruit, J. Theunis, N. Goswami, L. E. Vanfleteren, F. M. E. Franssen, E. F. M. Wouters, P. De Boever, Looking into the eye of patients with chronic obstructive pulmonary disease: an opportunity for better microvascular profiling of these complex patients Acta Ophthalmol 96 (2018), doi:10.1111/aos.13765.

18. M. Aronov, R. Allon, D. Stave, M. Belkin, E. Margalit, I. D. Fabian, B. Rosenzweig, Retinal vascular signs as screening and prognostic factors for chronic kidney disease: A systematic review and meta-analysis of current evidence. J Pers Med 11 (2021), doi:10.3390/jpm11070665.

19. J. Duvall-Young, C. D. Short, M. F. Raines, R. Gokal, W. Lawler, Fundus changes in mesangiocapillary glomerulonephritis type II: Clinical and fluorescein angiographic findings. British Journal of Ophthalmology 73 (1989), doi:10.1136/bjo.73.11.900.

20. C. E. McAvoy, G. Silvestri, Retinal changes associated with type 2 glomerulonephritis. Eye 19 (2005), doi:10.1038/sj.eye.6701697.

21. R. Bright, Tabular view of the morbid appearances in 100 cases connected with albuminous urine. Guy’s Hosp Rep. 1, 380–400 (1836).

22. N. M. Keith, H. P. Wagener, N. W. Barker, Some different types of essential hypertension: their course and prognosis. American Journal of the Medical Sciences 268 (1974), doi:10.1097/00000441-197412000-00004.

23. I. P. Chatziralli, The Value of Fundoscopy in General Practice. Open Ophthalmol J 6 (2012), doi:10.2174/1874364101206010004.

24. W. Drexler, J. G. Fujimoto, State-of-the-art retinal optical coherence tomography Prog Retin Eye Res 27 (2008), doi:10.1016/j.preteyeres.2007.07.005.

25. J. Fujimoto, E. Swanson, The development, commercialization, and impact of optical coherence tomography Invest Ophthalmol Vis Sci 57 (2016), doi:10.1167/iovs.16-19963.

26. L. Ngo, J. Cha, J. H. Han, Deep Neural Network Regression for Automated Retinal Layer Segmentation in Optical Coherence Tomography Images. IEEE Transactions on Image Processing 29 (2020), doi:10.1109/TIP.2019.2931461.

27. S. Guo, S. Yin, G. Tse, G. Li, L. Su, T. Liu, Association Between Caliber of Retinal Vessels and Cardiovascular Disease: a Systematic Review and Meta-Analysis Curr Atheroscler Rep 22 (2020), doi:10.1007/s11883-020-0834-2.

28. K. L. Thomson, J. M. Yeo, B. Waddell, J. R. Cameron, S. Pal, A systematic review and meta-analysis of retinal nerve fiber layer change in dementia, using optical coherence tomography*Alzheimer’s and Dementia: Diagnosis*, Assessment and Disease Monitoring 1 (2015), doi:10.1016/j.dadm.2015.03.001.

29. U. Mutlu, J. M. Colijn, M. A. Ikram, P. W. M. Bonnemaijer, S. Licher, F. J. Wolters, H. Tiemeier, P. J. Koudstaal, C. C. W. Klaver, M. K. Ikram, Association of Retinal Neurodegeneration on Optical Coherence Tomography with Dementia: A Population-Based Study. JAMA Neurol 75 (2018), doi:10.1001/jamaneurol.2018.1563.

30. T. Y. Wong, R. Klein, A. R. Sharrett, B. B. Duncan, D. J. Couper, J. M. Tielsch, B. E. K. Klein, L. D. Hubbard, Retinal arteriolar narrowing and risk of coronary heart disease in men and women: The Atherosclerosis Risk in Communities Study. J Am Med Assoc 287 (2002), doi:10.1001/jama.287.9.1153.

31. X. R. Gao, H. Huang, H. Kim, Genome-wide association analyses identify 139 loci associated with macular thickness in the UK Biobank cohort. Hum Mol Genet 28 (2019), doi:10.1093/hmg/ddy422.

32. H. Currant, P. Hysi, T. W. Fitzgerald, P. Gharahkhani, P. W. M. Bonnemaijer, A. Senabouth, A. W. Hewitt, D. Atan, T. Aung, J. Charng, H. Choquet, J. Craig, P. T. Khaw, C. C. W. Klaver, M. Kubo, J. S. Ong, L. R. Pasquale, C. A. Reisman, M. Daniszewski, J. E. Powell, A. Pébay, M. J. Simcoe, A. A. H. J. Thiadens, C. M. van Duijn, S. Yazar, E. Jorgenson, S. MacGregor, C. J. Hammond, D. A. Mackey, J. L. Wiggs, P. J. Foster, P. J. Patel, E. Birney, A. P. Khawaja, Genetic variation affects morphological retinal phenotypes extracted from UK Biobank optical coherence tomography images. PLoS Genet 17 (2021), doi:10.1371/journal.pgen.1009497.

33. C. Bycroft, C. Freeman, D. Petkova, G. Band, L. T. Elliott, K. Sharp, A. Motyer, D. Vukcevic, O. Delaneau, J. O’Connell, A. Cortes, S. Welsh, A. Young, M. Effingham, G. McVean, S. Leslie, N. Allen, P. Donnelly, J. Marchini, The UK Biobank resource with deep phenotyping and genomic data. Nature 562 (2018), doi:10.1038/s41586-018-0579-z.

34. P. A. Keane, C. M. Grossi, P. J. Foster, Q. Yang, C. A. Reisman, K. Chan, T. Peto, D. Thomas, P. J. Patel, Optical coherence tomography in the UK Biobank study - Rapid automated analysis of retinal thickness for large population-based studies. PLoS One 11 (2016), doi:10.1371/journal.pone.0164095.

35. P. Patel, P. J. Foster, C. Grossi, P. Keane, A. Lotery, T. Peto, C. Reisman, N. Strouthidis, Q. Yang, Spectral-domain optical coherence tomography imaging in 67,321 adults: associations with macular thickness in the UK Biobank Study. Ophthalmology (In press/epub) (2015).

36. S. M. Zekavat, C. Roselli, G. Hindy, S. A. Lubitz, P. T. Ellinor, H. Zhao, P. Natarajan, Genetic Link Between Arterial Stiffness and Atrial Fibrillation Circ Genom Precis Med 12 (2019), doi:10.1161/CIRCGEN.118.002453.

37. S. M. Zekavat, K. Aragam, C. Emdin, A. V. Khera, D. Klarin, H. Zhao, P. Natarajan, Genetic Association of Finger Photoplethysmography-Derived Arterial Stiffness Index with Blood Pressure and Coronary Artery Disease. Arterioscler Thromb Vasc Biol 39 (2019), doi:10.1161/ATVBAHA.119.312626.

38. S. M. Zekavat, M. Honigberg, J. P. Pirruccello, P. Kohli, E. W. Karlson, C. Newton-Cheh, H. Zhao, P. Natarajan, Elevated Blood Pressure Increases Pneumonia Risk: Epidemiological Association and Mendelian Randomization in the UK Biobank. Med 2 (2021), doi:10.1016/j.medj.2020.11.001.

39. S. J. Jurgens, S. H. Choi, V. N. Morrill, M. Chaffin, J. P. Pirruccello, J. L. Halford, L. C. Weng, V. Nauffal, C. Roselli, A. W. Hall, K. G. Aragam, K. L. Lunetta, S. A. Lubitz, P. T. Ellinor, Rare genetic variation underlying human diseases and traits: Results from 200,000 individuals in the uk biobank. bioRxiv (2020).

40. A. C. V. Van Hout, I. Tachmazidou, J. D. Backman, J. D. Hoffman, D. Liu, A. K. Pandey, C. Gonzaga-Jauregui, S. Khalid, B. Ye, N. Banerjee, A. H. Li, C. O’Dushlaine, A. Marcketta, J. Staples, C. Schurmann, A. Hawes, E. Maxwell, L. Barnard, A. Lopez, J. Penn, L. Habegger, A. L. Blumenfeld, X. Bai, S. O’Keeffe, A. Yadav, K. Praveen, M. Jones, W. J. Salerno, W. K. Chung, I. Surakka, C. J. Willer, K. Hveem, J. B. Leader, D. J. Carey, D. H. Ledbetter, L. Cardon, G. D. Yancopoulos, A. Economides, G. Coppola, A. R. Shuldiner, S. Balasubramanian, M. Cantor, M. R. Nelson, J. Whittaker, J. G. Reid, J. Marchini, J. D. Overton, R. A. Scott, G. R. Abecasis, L. Yerges-Armstrong, A. Baras, Exome sequencing and characterization of 49,960 individuals in the UK Biobank. Nature 586 (2020), doi:10.1038/s41586-020-2853-0.

41. S. M. Zekavat, V. K. Raghu, M. Trinder, Y. Ye, S. Koyama, M. C. Honigberg, Z. Yu, A. Pampana, S. Urbut, S. Haidermota, D. P. O’Regan, H. Zhao, P. T. Ellinor, A. V Segrè, T. Elze, J. L. Wiggs, J. Martone, R. A. Adelman, N. Zebardast, L. Del Priore, J. C. Wang, P. Natarajan, Deep Learning of the Retina Enables Phenome- and Genome-wide Analyses of the Microvasculature. Circulation (2021), doi:10.1161/circulationaha.121.057709.

42. P. Wu, A. Gifford, X. Meng, X. Li, H. Campbell, T. Varley, J. Zhao, R. Carroll, L. Bastarache, J. C. Denny, E. Theodoratou, W.-Q. Wei, Mapping ICD-10 and ICD-10-CM Codes to Phecodes: Workflow Development and Initial Evaluation. JMIR Med Inform 7 (2019), doi:10.2196/14325.

43. J. C. Denny, M. D. Ritchie, M. A. Basford, J. M. Pulley, L. Bastarache, K. Brown-Gentry, D. Wang, D. R. Masys, D. M. Roden, D. C. Crawford, PheWAS: Demonstrating the feasibility of a phenome-wide scan to discover gene-disease associations. Bioinformatics 26 (2010), doi:10.1093/bioinformatics/btq126.

44. H. K. Finucane, B. Bulik-Sullivan, A. Gusev, G. Trynka, Y. Reshef, P. R. Loh, V. Anttila, H. Xu, C. Zang, K. Farh, S. Ripke, F. R. Day, S. Purcell, E. Stahl, S. Lindstrom, J. R. B. Perry, Y. Okada, S. Raychaudhuri, M. J. Daly, N. Patterson, B. M. Neale, A. L. Price, Partitioning heritability by functional annotation using genome-wide association summary statistics. Nat Genet 47 (2015), doi:10.1038/ng.3404.

45. E. M. Weeks, J. C. Ulirsch, N. Y. Cheng, B. L. Trippe, R. S. Fine, J. Miao, T. A. Patwardhan, M. Kanai, J. Nasser, C. P. Fulco, K. C. Tashman, F. Aguet, T. Li, J. Ordovas-Montanes, C. S. Smillie, M. Biton, A. K. Shalek, A. N. Ananthakrishnan, R. J. Xavier, A. Regev, R. M. Gupta, K. Lage, K. G. Ardlie, J. N. Hirschhorn, E. S. Lander, J. M. Engreitz, H. K. Finucane, Leveraging polygenic enrichments of gene features to predict genes underlying complex traits and diseases medRxiv (2020), doi:10.1101/2020.09.08.20190561.

46. M. Loeffler, C. Engel, P. Ahnert, D. Alfermann, K. Arelin, R. Baber, F. Beutner, H. Binder, E. Brähler, R. Burkhardt, U. Ceglarek, C. Enzenbach, M. Fuchs, H. Glaesmer, F. Girlich, A. Hagendorff, M. Häntzsch, U. Hegerl, S. Henger, T. Hensch, A. Hinz, V. Holzendorf, D. Husser, A. Kersting, A. Kiel, T. Kirsten, J. Kratzsch, K. Krohn, T. Luck, S. Melzer, J. Netto, M. Nüchter, M. Raschpichler, F. G. Rauscher, S. G. Riedel-Heller, C. Sander, M. Scholz, P. Schönknecht, M. L. Schroeter, J.-C. Simon, R. Speer, J. Stäker, R. Stein, Y. Stöbel-Richter, M. Stumvoll, A. Tarnok, A. Teren, D. Teupser, F. S. Then, A. Tönjes, R. Treudler, A. Villringer, A. Weissgerber, P. Wiedemann, S. Zachariae, K. Wirkner, J. Thiery, The LIFE-Adult-Study: objectives and design of a population-based cohort study with 10,000 deeply phenotyped adults in Germany. BMC Public Health 15, 691 (2015).

47. J. Mbatchou, L. Barnard, J. Backman, A. Marcketta, J. A. Kosmicki, A. Ziyatdinov, C. Benner, C. O’Dushlaine, M. Barber, B. Boutkov, L. Habegger, M. Ferreira, A. Baras, J. Reid, G. Abecasis, E. Maxwell, J. Marchini, Computationally efficient whole-genome regression for quantitative and binary traits. Nat Genet 53 (2021), doi:10.1038/s41588-021-00870-7.

48. D. G. MacArthur, S. Balasubramanian, A. Frankish, N. Huang, J. Morris, K. Walter, L. Jostins, L. Habegger, J. K. Pickrell, S. B. Montgomery, C. A. Albers, Z. D. Zhang, D. F. Conrad, G. Lunter, H. Zheng, Q. Ayub, M. A. DePristo, E. Banks, M. Hu, R. E. Handsaker, J. A. Rosenfeld, M. Fromer, M. Jin, X. J. Mu, E. Khurana, K. Ye, M. Kay, G. I. Saunders, M. M. Suner, T. Hunt, I. H. A. Barnes, C. Amid, D. R. Carvalho-Silva, A. H. Bignell, C. Snow, Yngvadottir, S. Bumpstead, D. N. Cooper, Y. Xue, I. G. Romero, J. Wang, Y. Li, R. A. Gibbs, S. A. McCarroll, E. T. Dermitzakis, J. K. Pritchard, J. C. Barrett, J. Harrow, M. E. Hurles, M. B. Gerstein, C. Tyler-Smith, A systematic survey of loss-of-function variants in human protein-coding genes. Science *(1979)* 335 (2012), doi:10.1126/science.1215040.

49. C. Dong, P. Wei, X. Jian, R. Gibbs, E. Boerwinkle, K. Wang, X. Liu, Comparison and integration of deleteriousness prediction methods for nonsynonymous SNVs in whole exome sequencing studies. Hum Mol Genet 24 (2015), doi:10.1093/hmg/ddu733.

50. S. M. Zekavat, S.-H. Lin, A. G. Bick, A. Liu, K. Paruchuri, C. Wang, M. M. Uddin, Y. Ye, Z. Yu, X. Liu, Y. Kamatani, R. Bhattacharya, J. P. Pirruccello, A. Pampana, P.-R. Loh, P. Kohli, S. A. McCarroll, K. Kiryluk, B. Neale, I. Ionita-Laza, E. A. Engels, D. W. Brown, J. W. Smoller, R. Green, E. W. Karlson, M. Lebo, P. T. Ellinor, S. T. Weiss, M. J. Daly, Biobank Japan Project, FinnGen Consortium, C. Terao, H. Zhao, B. L. Ebert, M. P. Reilly, A. Ganna, M. J. Machiela, G. Genovese, P. Natarajan, Hematopoietic mosaic chromosomal alterations increase the risk for diverse types of infection. Nat Med 27, 1012–1024 (2021).

51. M. K. Ikram, X. Sim, S. Xueling, R. A. Jensen, M. F. Cotch, A. W. Hewitt, M. A. Ikram, J. J. Wang, R. Klein, B. E. K. Klein, M. M. B. Breteler, N. Cheung, G. Liew, P. Mitchell, A. G. Uitterlinden, F. Rivadeneira, A. Hofman, P. T. V. M. de Jong, C. M. van Duijn, L. Kao, C.-Y. Cheng, A. V. Smith, N. L. Glazer, T. Lumley, B. McKnight, B. M. Psaty, F. Jonasson, G. Eiriksdottir, T. Aspelund, Global BPgen Consortium, T. B. Harris, L. J. Launer, K. D. Taylor, X. Li, S. K. Iyengar, Q. Xi, T. A. Sivakumaran, D. A. Mackey, S. Macgregor, N. G. Martin, T. L. Young, J. C. Bis, K. L. Wiggins, S. R. Heckbert, C. J. Hammond, T. Andrew, S. Fahy, J. Attia, E. G. Holliday, R. J. Scott, F. M. A. Islam, J. I. Rotter, A. K. McAuley, E. Boerwinkle, E. S. Tai, V. Gudnason, D. S. Siscovick, J. R. Vingerling, T. Y. Wong, Four novel Loci (19q13, 6q24, 12q24, and 5q14) influence the microcirculation in vivo. PLoS Genet 6, e1001184 (2010).

52. J. E. Craig, X. Han, A. Qassim, M. Hassall, J. N. Cooke Bailey, T. G. Kinzy, A. P. Khawaja, J. An, H. Marshall, P. Gharahkhani, R. P. Igo, S. L. Graham, P. R. Healey, J.-S. Ong, T. Zhou, O. Siggs, M. H. Law, E. Souzeau, B. Ridge, P. G. Hysi, K. P. Burdon, R. A. Mills, J. Landers, J. B. Ruddle, A. Agar, A. Galanopoulos, A. J. R. White, C. E. Willoughby, N. H. Andrew, S. Best, A. L. Vincent, I. Goldberg, G. Radford-Smith, N. G. Martin, G. W. Montgomery, V. Vitart, R. Hoehn, R. Wojciechowski, J. B. Jonas, T. Aung, L. R. Pasquale, A. J. Cree, S. Sivaprasad, N. A. Vallabh, NEIGHBORHOOD consortium, UK Biobank Eye and Vision Consortium, A. C. Viswanathan, F. Pasutto, J. L. Haines, C. C. W. Klaver, C. M. van Duijn, R. J. Casson, P. J. Foster, P. T. Khaw, C. J. Hammond, D. A. Mackey, P. Mitchell, A. J. Lotery, J. L. Wiggs, A. W. Hewitt, S. MacGregor, Multitrait analysis of glaucoma identifies new risk loci and enables polygenic prediction of disease susceptibility and progression. Nat Genet 52, 160–166 (2020).

53. H. Choquet, R. B. Melles, D. Anand, J. Yin, G. Cuellar-Partida, W. Wang, 23andMe Research Team, T. J. Hoffmann, K. S. Nair, P. G. Hysi, S. A. Lachke, E. Jorgenson, A large multiethnic GWAS meta-analysis of cataract identifies new risk loci and sex-specific effects. Nat Commun 12, 3595 (2021).

54. C. Engel, K. Wirkner, S. Zeynalova, R. Baber, H. Binder, U. Ceglarek, C. Enzenbach, M. Fuchs, A. Hagendorff, S. Henger, A. Hinz, F. G. Rauscher, M. Reusche, S. G. Riedel-Heller, S. Röhr, J. Sacher, C. Sander, M. L. Schroeter, A. Tarnok, R. Treudler, A. Villringer, R. Wachter, A. V. Witte, J. Thiery, M. Scholz, M. Loeffler, LIFE-Adult-Study working group, Cohort Profile: The LIFE-Adult-Study. Int J Epidemiol (2022), doi:10.1093/ije/dyac114.

55. I. Adzhubei, D. M. Jordan, S. R. Sunyaev, Predicting functional effect of human missense mutations using PolyPhen-2. Curr Protoc Hum Genet **Chapter** 7, Unit7.20 (2013).

56. P. C. Ng, S. Henikoff, SIFT: Predicting amino acid changes that affect protein function. Nucleic Acids Res 31, 3812–4 (2003).

57. Q. Lu, B. Li, D. Ou, M. Erlendsdottir, R. L. Powles, T. Jiang, Y. Hu, D. Chang, C. Jin, W. Dai, Q. He, Z. Liu, S. Mukherjee, P. K. Crane, H. Zhao, A Powerful Approach to Estimating Annotation-Stratified Genetic Covariance via GWAS Summary Statistics. Am J Hum Genet 101, 939–964 (2017).

58. G. D. Smith, S. Ebrahim, “Mendelian randomization”: Can genetic epidemiology contribute to understanding environmental determinants of disease?Int J Epidemiol 32 (2003), doi:10.1093/ije/dyg070.

59. R. A. C. van Huet, R. W. J. Collin, A. M. Siemiatkowska, C. C. W. Klaver, C. B. Hoyng, F. Simonelli, M. I. Khan, R. Qamar, E. Banin, F. P. M. Cremers, T. Theelen, A. I. den Hollander, L. I. van den Born, B. J. Klevering, IMPG2-associated retinitis pigmentosa displays relatively early macular involvement. Invest Ophthalmol Vis Sci 55, 3939–53 (2014).

60. S. M. Shah, L. A. Schimmenti, A. D. Marmorstein, S. J. Bakri, ADULT-ONSET VITELLIFORM MACULAR DYSTROPHY SECONDARY TO A NOVEL IMPG2 GENE VARIANT. Retin Cases Brief Rep 15, 356–358 (2021).

61. I. Vázquez-Domínguez, C. H. Z. Li, Z. Fadaie, L. Haer-Wigman, F. P. M. Cremers, A. Garanto, C. B. Hoyng, S. Roosing, Identification of a Complex Allele in IMPG2 as a Cause of Adult-Onset Vitelliform Macular Dystrophy. Invest Ophthalmol Vis Sci 63, 27 (2022).

62. Z. Zhu, D. Shi, G. Peng, Z. Tan, X. Shang, W. Hu, H. Liao, X. Zhang, Y. Huang, H. Yu, W. Meng, W. Wang, X. Yang, M. He, Retinal age as a predictive biomarker for mortality riskmedRxiv (2020).

63. H. Currant, P. Hysi, T. W. Fitzgerald, P. Gharahkhani, P. W. M. Bonnemaijer, A. Senabouth, A. W. Hewitt, UK Biobank Eye and Vision Consortium, International Glaucoma Genetics Consortium, D. Atan, T. Aung, J. Charng, H. Choquet, J. Craig, P. T. Khaw, C. C. W. Klaver, M. Kubo, J.-S. Ong, L. R. Pasquale, C. A. Reisman, M. Daniszewski, J. E. Powell, A. Pébay, M. J. Simcoe, A. A. H. J. Thiadens, C. M. van Duijn, S. Yazar, E. Jorgenson, S. MacGregor, C. J. Hammond, D. A. Mackey, J. L. Wiggs, P. J. Foster, P. J. Patel, E. Birney, A. P. Khawaja, Genetic variation affects morphological retinal phenotypes extracted from UK Biobank optical coherence tomography images. PLoS Genet 17, e1009497 (2021).

64. S. M. Zekavat, S. Sekimitsu, Y. Ye, V. Raghu, H. Zhao, T. Elze, A. V Segrè, J. L. Wiggs, P. Natarajan, L. Del Priore, N. Zebardast, J. C. Wang, Photoreceptor Layer Thinning Is an Early Biomarker for Age-Related Macular Degeneration: Epidemiologic and Genetic Evidence from UK Biobank OCT Data. Ophthalmology 129, 694–707 (2022).

65. A. C. Balmforth, J. J. M. H. van Bragt, T. Ruijs, J. R. Cameron, R. Kimmitt, R. Moorhouse, Czopek, M. K. Hu, P. J. Gallacher, J. W. Dear, S. Borooah, I. M. MacIntyre, T. M. C. Pearson, L. Willox, D. Talwar, M. Tafflet, C. Roubeix, F. Sennlaub, S. Chandran, B. Dhillon, D. J. Webb, N. Dhaun, Chorioretinal thinning in chronic kidney disease links to inflammation and endothelial dysfunction. JCI Insight 1 (2016), doi:10.1172/jci.insight.89173.

66. H. Hwang, J. B. Chae, J. Y. Kim, B. G. Moon, B. G. Moon, D. Y. Kim, Changes in optical coherence tomography findings in patients with chronic renal failure undergoing dialysis for the first time. Retina 39 (2019), doi:10.1097/iae.0000000000002312.

67. P. G. Theodossiadis, S. Theodoropoulou, G. Neamonitou, V. Grigoropoulos, V. Liarakos, E. Triantou, G. P. Theodossiadis, D. V. Vlahakos, Hemodialysis-induced alterations in macular thickness measured by optical coherence tomography in diabetic patients with end-stage renal disease. Ophthalmologica 227 (2012), doi:10.1159/000331321.

68. D. Pahor, B. Gracner, T. Gracner, R. Hojs, Optical Coherence Tomography Findings in Hemodialysis Patients. Klin Monbl Augenheilkd 1 (2007), doi:10.1055/s-2007-963761.

69. J. W. Jung, M. H. Yoon, S. W. Lee, H. S. Chin, Effect of hemodialysis (HD) on intraocular pressure, ocular surface, and macular change in patients with chronic renal failure: Effect of hemodialysis on the ophthalmologic findings. Graefe’s Archive for Clinical and Experimental Ophthalmology 251 (2013), doi:10.1007/s00417-012-2032-6.

70. D. Colville, J. Savige, P. Branley, D. Wilson, Ocular abnormalities in thin basement membrane disease. British Journal of Ophthalmology 81 (1997), doi:10.1136/bjo.81.5.373.

71. S. M. Zekavat, V. K. Raghu, M. Trinder, Y. Ye, S. Koyama, M. C. Honigberg, Z. Yu, A. Pampana, S. Urbut, S. Haidermota, D. P. O’Regan, H. Zhao, P. T. Ellinor, A. V. Segrè, T. Elze, J. L. Wiggs, J. Martone, R. A. Adelman, N. Zebardast, L. Del Priore, J. C. Wang, P. Natarajan, Deep Learning of the Retina Enables Phenome- and Genome-wide Analyses of the Microvasculature. Circulation (2021), doi:10.1161/circulationaha.121.057709.

72. T. Y. Wong, R. Klein, A. R. Sharrett, T. A. Manolio, L. D. Hubbard, E. K. Marino, L. Kuller, G. Burke, R. P. Tracy, J. F. Polak, J. S. Gottdiener, D. S. Siscovick, The prevalence and risk factors of retinal microvascular abnormalities in older persons: The cardiovascular health study. Ophthalmology 110 (2003), doi:10.1016/S0161-6420(02)01931-0.

73. T. Y. Wong, R. Klein, D. J. Couper, L. S. Cooper, E. Shahar, L. D. Hubbard, M. R. Wofford, A. R. Sharrett, Retinal microvascular abnormalities and incident stroke: The Atherosclerosis Risk in Communities Study. Lancet 358 (2001), doi:10.1016/S0140-6736(01)06253-5.

74. R. J. Thomson, J. Chazaro, O. Otero-Marquez, G. Ledesma-Gil, Y. Tong, A. C. Coughlin, Z. R. Teibel, S. Alauddin, K. Tai, H. Lloyd, M. Scolaro, A. Govindaiah, A. Bhuiyan, M. S. Dhamoon, A. Deobhakta, J. Narula, R. B. Rosen, L. A. Yannuzzi, K. B. Freund, R. T. Smith, SUBRETINAL DRUSENOID DEPOSITS AND SOFT DRUSEN: Are They Markers for Distinct Retinal Diseases? Retina 42, 1311–1318 (2022).

75. C.-C. Chang, C.-H. Huang, Y.-C. Chou, J.-Y. Chang, C.-A. Sun, Association Between Age-Related Macular Degeneration and Risk of Heart Failure: A Population-Based Nested Case-Control Study. J Am Heart Assoc 10, e020071 (2021).

76. M. M. Mauschitz, R. P. Finger, Age-Related Macular Degeneration and Cardiovascular Diseases: Revisiting the Common Soil Theory. Asia Pac J Ophthalmol (Phila) 11, 94–99.

77. W. Bin Wei, L. Xu, J. B. Jonas, L. Shao, K. F. Du, S. Wang, C. X. Chen, J. Xu, Y. X. Wang, J. Q. Zhou, Q. S. You, Subfoveal choroidal thickness: The Beijing Eye Study. Ophthalmology 120 (2013), doi:10.1016/j.ophtha.2012.07.048.

78. G. Mulè, M. Vadalà, T. La Blasca, R. Gaetani, G. Virone, M. Guarneri, M. Castellucci, G. Guarrasi, M. Terrasi, S. Cottone, Association between early-stage chronic kidney disease and reduced choroidal thickness in essential hypertensive patients. Hypertension Research 42 (2019), doi:10.1038/s41440-018-0195-1.

79. M. Kong, Y. Kwun, J. Sung, D. Il Ham, Y. M. Song, Association between systemic hypertension and macular thickness measured by optical coherence tomography. Invest Ophthalmol Vis Sci 56 (2015), doi:10.1167/iovs.14-16080.

80. L. Shao, L. X. Zhou, L. Xu, W. Bin Wei, The relationship between Subfoveal Choroidal Thickness and Hypertensive Retinopathy. Sci Rep 11 (2021), doi:10.1038/s41598-021-84947-7.

81. M. G. Abd El-Naser, H. M. Abd El-Rahman, Z. R. Adawy, M. M. Aly, Optical Coherence Tomography Study of Retinal and Choroidal Changes in Patients with Chronic Obstructive Pulmonary Disease. Egypt J Hosp Med 75 (2019), doi:10.21608/ejhm.2019.30967.

82. Ö. Kocamiş, D. Zorlu, Choroid and Retinal Nerve Fiber Layer Thickness in Patients with Chronic Obstructive Pulmonary Disease Exacerbation. J Ophthalmol 2018 (2018), doi:10.1155/2018/1201976.

83. S. Alim, H. D. Demir, A. Yilmaz, S. Demir, A. Güneş, in *Journal of Ophthalmology*, (2019), vol. 2019.

84. M. Gok, M. A. Ozer, S. Ozen, B. Botan Yildirim, The evaluation of retinal and choroidal structural changes by optical coherence tomography in patients with chronic obstructive pulmonary disease. Curr Eye Res 43 (2018), doi:10.1080/02713683.2017.1373824.

85. N. O. Ahmed, Y. M. Shaaban, H. G. Ezzelregal, Evaluation of the impact of COPD severity grading and oxygen saturation on the retinal nerve fiber layer thickness and subfoveal choroidal thickness in COPD patients. The Egyptian Journal of Bronchology 15 (2021), doi:10.1186/s43168-021-00092-9.

86. P. M. Marinho, A. A. A. Marcos, A. C. Romano, H. Nascimento, R. Belfort, Retinal findings in patients with COVID-19The Lancet 395 (2020), doi:10.1016/S0140-6736(20)31014-X.

87. B. Burgos-Blasco, N. Güemes-Villahoz, B. Vidal-Villegas, J. M. Martinez-de-la-Casa, J. Donate-Lopez, F. J. Martín-Sánchez, J. J. González-Armengol, J. Porta-Etessam, J. L. R. Martin, J. Garcia-Feijoo, Optic nerve and macular optical coherence tomography in recovered COVID-19 patients. Eur J Ophthalmol (2021), doi:10.1177/11206721211001019.

88. D. Szkodny, E. Wylęgała, P. Sujka-Franczak, E. Chlasta-Twardzik, R. Fiolka, T. Tomczyk, A. Wylęgała, Retinal oct findings in patients after covid infection. J Clin Med 10 (2021), doi:10.3390/jcm10153233.

89. D. G. Vavvas, D. Sarraf, S. V. R. Sadda, D. Eliott, J. P. Ehlers, N. K. Waheed, Y. Morizane, T. Sakamoto, M. Tsilimbaris, J. B. Miller, Concerns about the interpretation of OCT and fundus findings in COVID-19 patients in recent Lancet publication Eye (Basingstoke) 34 (2020), doi:10.1038/s41433-020-1084-9.

90. A. Wylęgała, D. Szkodny, E. Wylęgała, Comment on: Optical Coherence Tomography Angiography Features in Post-COVID-19 Pneumonia Patients: A Pilot Study. Am J Ophthalmol (2021), doi:10.1016/j.ajo.2021.08.021.

91. E. Ugurlu, G. Pekel, B. Cengiz, K. Bozkurt, G. Altinisik, in Sarcoidosis Vasculitis and Diffuse Lung Diseases, (2017), vol. 34.

92. J. Britze, J. L. Frederiksen, Optical coherence tomography in multiple sclerosis Eye (Basingstoke) 32 (2018), doi:10.1038/s41433-017-0010-2.

93. A. Petzold, J. F. de Boer, S. Schippling, P. Vermersch, R. Kardon, A. Green, P. A. Calabresi, C. Polman, Optical coherence tomography in multiple sclerosis: A systematic review and meta-analysis Lancet Neurol 9 (2010), doi:10.1016/S1474-4422(10)70168-X.

94. A. Z. A. Tak, Y. Şengül, B. Ekmekçi, A. S. Karadağ, Comparison of optic coherence tomography results in patients with diagnosed epilepsy: Findings in favor of neurodegeneration. Epilepsy and Behavior 92 (2019), doi:10.1016/j.yebeh.2018.12.021.

95. N. Bayraktar Bilen, A. P. Titiz, S. Bilen, B. Polat Gultekin, M. Sahin Hamurcu, D. Kalayci, Optical coherence tomography and neurodegeneration in epilepsy. Eur J Ophthalmol 31 (2021), doi:10.1177/1120672119881982.

96. P. Vermersch, O. Outteryck, A. Petzold, Optical coherence tomography - A new monitoring tool for multiple sclerosis? Eur Neurol Rev 5 (2010), doi:10.17925/ENR.2010.05.01.73.

97. J. J. Cerqueira, D. A. S. Compston, R. Geraldes, M. M. Rosa, K. Schmierer, A. Thompson, M. Tinelli, J. Palace, Time matters in multiple sclerosis: Can early treatment and long-term follow-up ensure everyone benefits from the latest advances in multiple sclerosis?J Neurol Neurosurg Psychiatry 89 (2018), doi:10.1136/jnnp-2017-317509.

98. M. H. Orum, A. Kalenderoglu, Decreases in retinal nerve fiber layer thickness correlates with cumulative alcohol intake. J Addict Dis 38 (2020), doi:10.1080/10550887.2020.1776083.

99. Y. Liu, L. Huang, Z. Wang, J. Chen, Q. bian, J. Sun, L. Jiang, F. Yang, The changes in retinal nerve fiber layer and macular thickness in Chinese patients with alcohol dependency. Drug Alcohol Depend 229 (2021), doi:10.1016/j.drugalcdep.2021.109130.

100. S. álvarez-Sesmero, F. J. Povedano-Montero, F. Arias-Horcajadas, M. Marín-Mayor, P. Navarrete-Chamorro, I. Raga-Martínez, G. Rubio, F. López-Muñoz, Retinal nerve fiber layer in patients with alcohol use disorder. Applied Sciences (Switzerland) 9 (2019), doi:10.3390/app9245331.

101. T. K. Yang, X. G. Huang, J. Y. Yao, Effects of Cigarette Smoking on Retinal and Choroidal Thickness: A Systematic Review and Meta-Analysis J Ophthalmol 2019 (2019), doi:10.1155/2019/8079127.

102. A. Fry, T. J. Littlejohns, C. Sudlow, N. Doherty, L. Adamska, T. Sprosen, R. Collins, N. E. Allen, Comparison of Sociodemographic and Health-Related Characteristics of UK Biobank Participants With Those of the General Population. Am J Epidemiol 186, 1026–1034 (2017).

