## Supplementary Figures for "Insights into human health from phenome- and genome-wide analyses of UK Biobank retinal optical coherence tomography phenotypes"

**Supplementary Information Table of Contents:**

A. **Supplementary Methods**

1. Supplementary Note 1: Quality control filters used in whole exome sequencing data

B. **Supplementary Figures**

1. **Supplementary Figure 1:** Comparison of right versus left eye retinal layer thicknesses.
2. **Supplementary Figure 2:** Univariate associations of layer thickness with **a**. sex, b. smoking status, **c**. age, height, weight, and spherical equivalent.
3. **Supplementary Figure 3:** Associations of retinal layer thickness with glaucoma and AMD.
4. **Supplementary Figure 4:** Associations of retinal layer thickness with mortality.
5. **Supplementary Figure 5:** Associations of retinal layer thickness with incident mortality using different covariates in a Cox survival model.
6. **Supplementary Figure 6:** Associations of prevalent conditions with retinal layer thickness.
7. **Supplementary Figure 7:** Associations of retinal layer thickness with incident disease.
8. **Supplementary Figure 8:** Significant associations effect sizes of retinal layer thickness on incident disease.
9. **Supplementary Figure 9:** Associations of clinical quantitative traits with retinal layer thickness.
10. **Supplementary Figure 10:** Phenotypic associations with photoreceptor segment (PS) layer thickness after additionally adjusting for hypertension (incident or prevalent combined), type 2 diabetes (incident or prevalent combined), HbA1c, and a 25-factor smoking covariate.
11. **Supplementary Figure 11:** Manhattan plot of genome-wide significant association results for the retinal layer thicknesses.
12. **Supplementary Figure 12:** Relative association effects of genome-wide significant association results compared by retinal layer.
13. **Supplementary Figure 13:** Genome-wide association results plotted by layer for GCC, RNFL, and GCL through Manhattan plots.
14. **Supplementary Figure 14:** Genome-wide association results plotted by layer for IPL, INL, and OPL+ONL through Manhattan plots.
15. **Supplementary Figure 15:** Genome-wide association results plotted by layer for PS, RPE+BM, and CSI through Manhattan plots.
16. **Supplementary Figure 16:** Top POPs genes within 1MB of a top variant by chromosome colored by retinal layer
17. **Supplementary Figure 17:** Top POPs genes by chromosome and retinal layer for the GCC, RNFL, and GCL layers.
18. **Supplementary Figure 18:** Top POPs genes by chromosome and retinal layer for the INL, IPL, and ONL+OPL layers.
19. **Supplementary Figure 19:** Top POPs genes by chromosome and retinal layer for the PS, RPE+BM, and CSI layers.
20. **Supplementary Figure 20:** Reactome gene set enrichment analysis analysis using POPs genes with score > 1 across the GCC, RNFL, and GCL layers.
21. **Supplementary Figure 21:** Reactome gene set enrichment analysis analysis using POPs genes with score > 1 across the INL, IPL, and ONL+OPL layers.
22. **Supplementary Figure 22:** Reactome gene set enrichment analysis analysis using POPs genes with score > 1 across the PS, RPE, and CSI layers.
23. **Supplementary Figure 23:** Replication of GWAS summary statistics in the LIFE cohort. Correlation of UKBB versus LIFE beta and 95% CI across top 259 independent, genome-wide significant variants identified in UKBB plotted by layer.
24. **Supplementary Figure 24:** 1-sample Mendelian randomization analysis, of retinal layer thickness polygenic risk scores with combined incident and prevalent phenotypes.
25. **Supplementary Figure 25:** Comparative epidemiological (a) versus genetic (b) significant and concordant associations of retinal layer thickness with common ophthalmic conditions.
26. **Supplementary Figure 26:** Comparative (a) epidemiological versus (b) genetic significant and concordant associations of quantitative phenotypes with retinal layer thickness.

### Supplementary Methods

#### Supplementary Methods:

**Supplementary Note 1:**

**Quality control filters used in whole exome sequencing data**

In addition to any quality-control that was performed centrally, we applied extensive additional genotype, variant and sample quality-control procedures to ensure a high-quality dataset for analyses for both the common variant analyses using array genotyping data and rare variant analyses using whole exome sequencing data.

For the whole exome sequencing data used in rare variant analyses, we utilized the OQFE WES pVCF files provided by the UK Biobank, which contained calls for 200,643 sequenced samples. We applied genotype refinement to the raw genotype calls in the pVCF files using Hail-0.2 (https://hail.is/docs/0.2/index.html). We first split multi-allelic sites to represent separate bi-allelic sites.

We first performed genotype quality control, filtering out calls that did not pass the following hard filters were then set to no-call in our analysis:

- For homozygous reference calls: Genotype Quality < 20; Genotype Depth < 10; Genotype Depth > 200
- For heterozygous calls: (A1 Depth + A2 Depth)/Total Depth < 0.9; A2 Depth/Total Depth < 0.2; Genotype likelihood[ref/ref] < 20; Genotype Depth < 10; Genotype Depth > 200
- For homozygous alternative calls: (A1 Depth + A2 Depth)/Total Depth < 0.9; A2 Depth/Total Depth < 0.9; Genotype likelihood[ref/ref] < 20; Genotype Depth < 10; Genotype Depth > 200

These filters removed 9% of the 3,573,574,459,423 raw genotype calls leaving 3,214,727,581,104 genotype calls across 17,981,897 variant sites and 200,643 samples.

We then performed variant-level quality control. We removed variants that failed the following filters:

- Call rate of < 90% (restricting to males for Y chromosomal markers) (N= 4,023,284)
- Failed a liberal Hardy-Weinberg Equilibrium test (HWE) at $P < 10^{-14}$ among unrelated samples (not applied to Y chromosomal markers) (N=136,869)
- Present in Ensembl low-complexity regions (N=748,116)
- Monomorphic in the final dataset (N=55,614)

After performing these variant filters, 13,003,057 variants remained of which 12,756,075 were autosomal.

To perform sample level quality control, we computed a number of quality metrics to identify bad-quality or duplicated samples. We first used KING6 (version 2.2.5) to calculate pairwise heterozygote concordance rates for each pair of samples, using the high-quality independent autosomal markers. Then we used the high-quality autosomal variants present in both WES and array datasets to compute per-sample heterozygote concordance rates between WES calls and genotyping array calls. We inferred the genetic sex of each participants with the --check-sex option in PLINK, using the high-quality independent X-chromosomal markers. We set any sample with F > 0.8 to male, while samples with F < 0.5 were set to female. Finally, using all ~12.7M autosomal WES variants, we computed a number of additional metrics including sample call rate, transition/transversion ratio (Ti/Tv), heterozygote/homozygote ratio (Het/Hom), SNV/indel ratio (SNV/indel) and the number of singletons. After computing these metrics, we excluded participants based on the following criteria:

- Decided to revoke their consent
- Sample duplicates based on heterozygote concordance rates > 0.8 (N=0)
- Samples with blatant discordance between self-reported and genetically inferred sex
- Discordance between WES and array calls with heterozygote concordance rates < 0.8
- Call rate < 90%
- Samples further than 8 standard deviations from the mean for Ti/Tv (n=0), Het/Hom (N=100), SNV/indel (N=1) and number of singletons (N=111)

After applying these filters 200,337 samples remained for analysis.

### Supplementary Figures

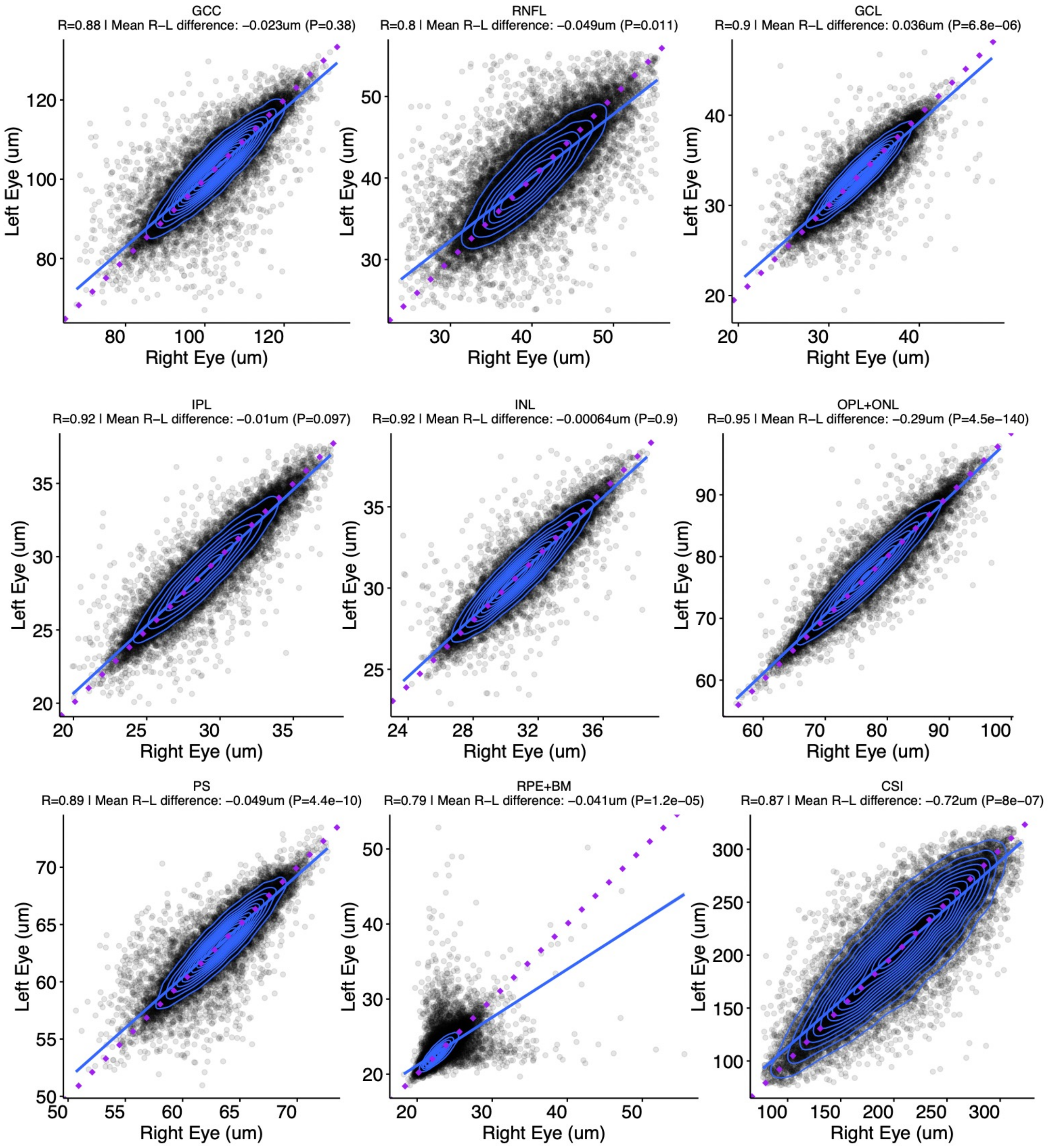


**Supplementary Figure 1: Comparison of right versus left eye retinal layer thicknesses.** Right versus left eye retinal layer thicknesses in um are plotted for each analyzed retinal layer. Blue line and contours reflects the best fit to the data, and purple line reflects the unity line (y=x). Spearman correlation (R) and mean right – left difference P-value is provided using a two sample paired t-test.

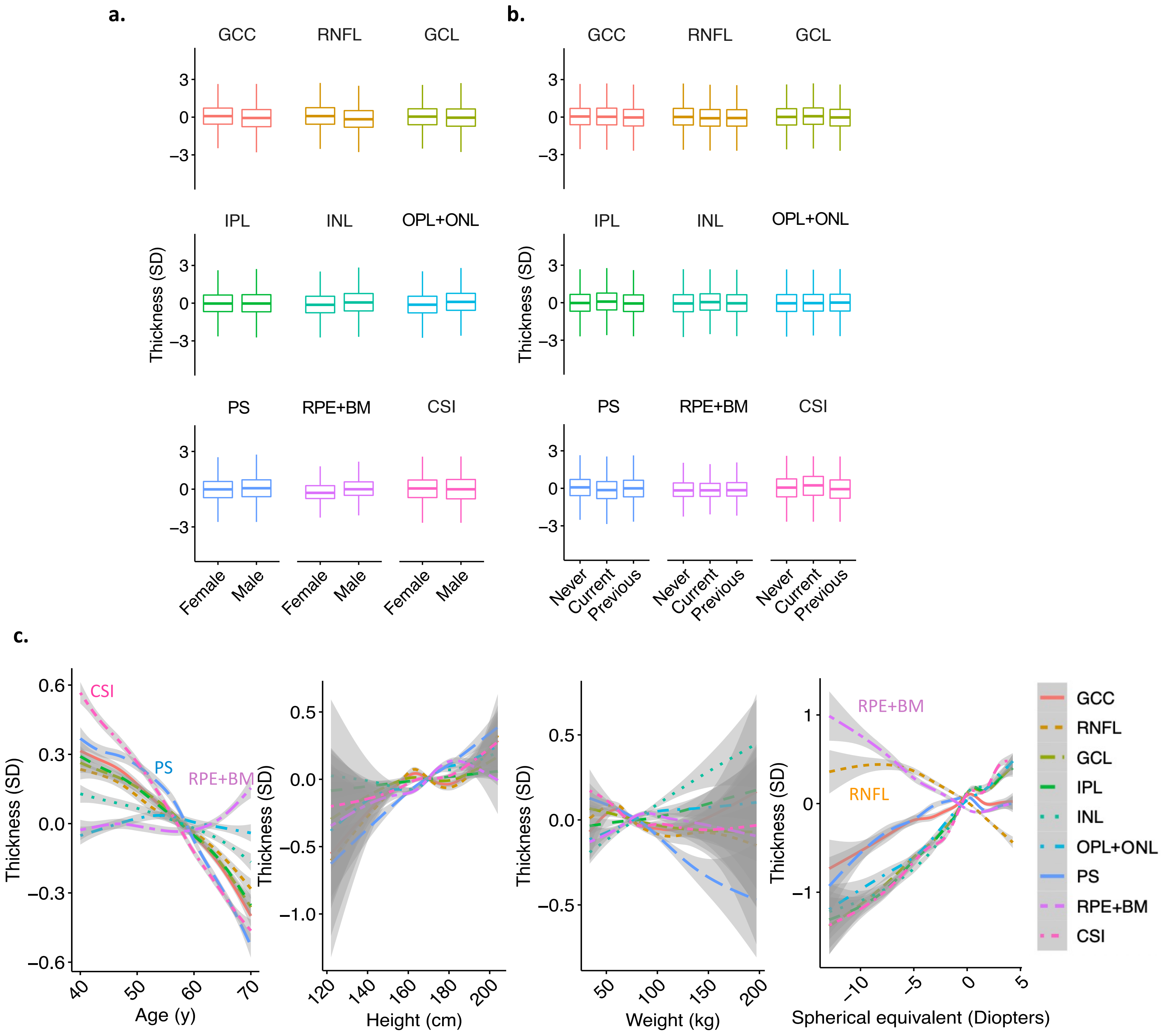


**Supplementary Figure 2:** Univariate associations of layer thickness with **a**. sex, b. smoking status, **c**. age, height, weight, and spherical equivalent.

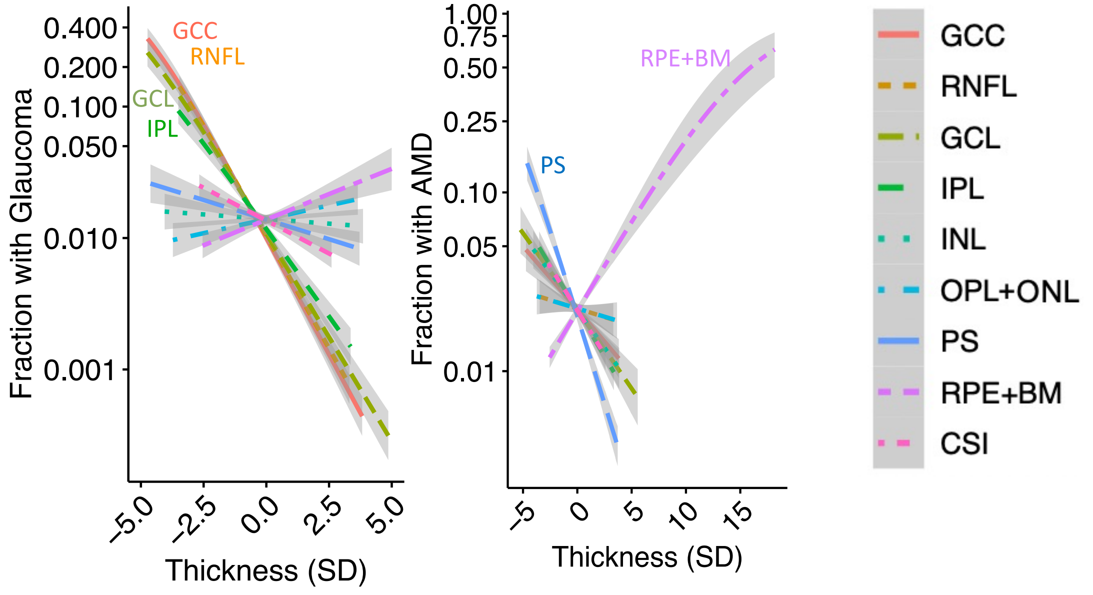


**Supplementary Figure 3:** Associations of retinal layer thickness with glaucoma and AMD.

**a.**

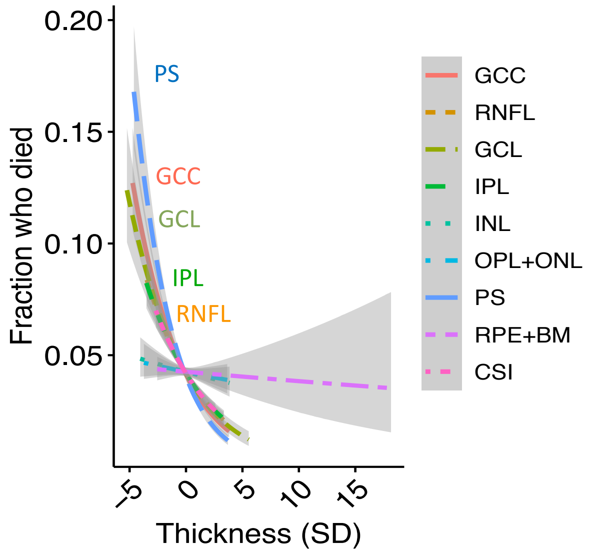


**b.**

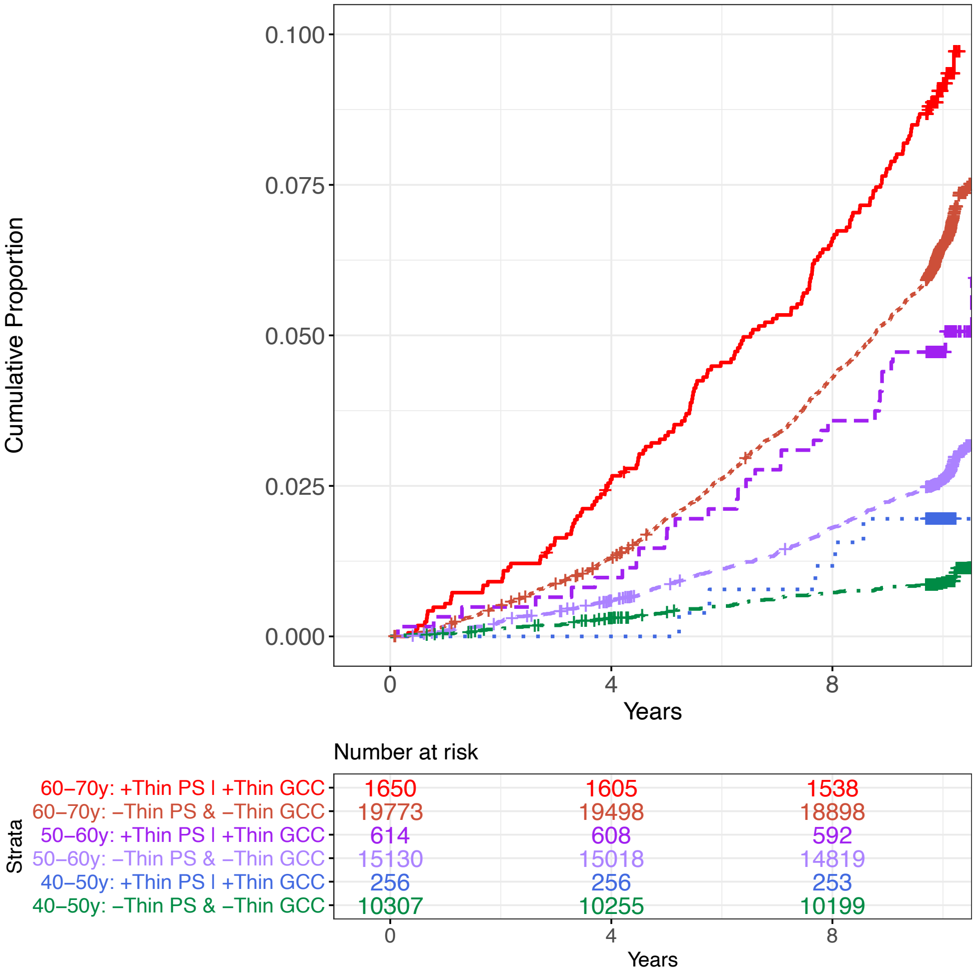


**Supplementary Figure 4:** Associations of retinal layer thickness with mortality. a) association spline plots of retinal layer thickness with incident mortality, with layers labeled showing negative correlation with mortality. b) Kaplan-Meyer curve showing the cumulative proportion of individuals who died by year after enrollment stratified by age deciles, as well as whether they had thin (defined as >2 SD lower than the mean) PS or thin GCC compared to individuals who didn't have thin PS and did not have thin GCC layers.

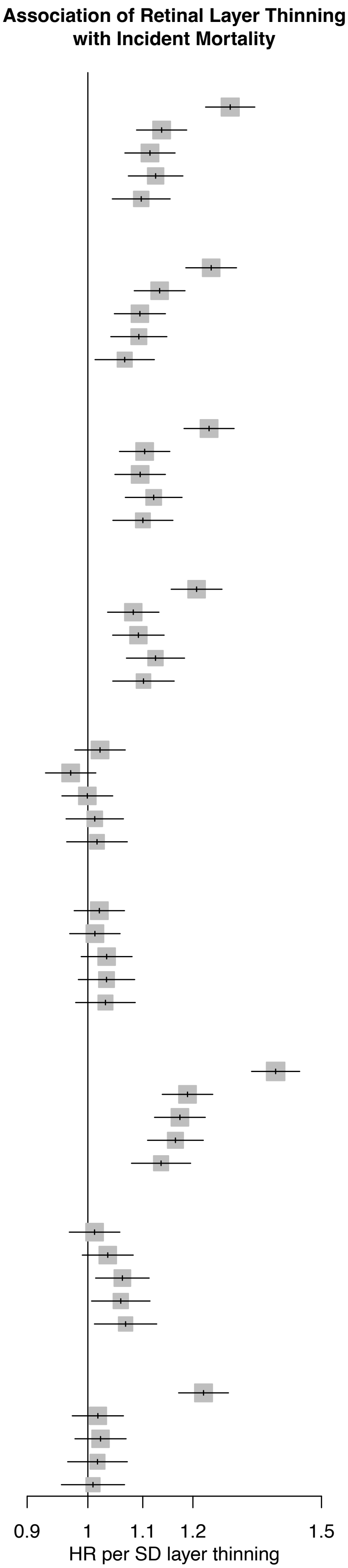


| | HR | 95% CI | P | Cases (N) | Controls (N) |
|---|---|---|---|---|---|
| **GCC** | | | | | |
| Unadjusted | 1.28 | [1.23; 1.34] | 9.5e−30 | 2,025 | 45,705 |
| +age,age^2 | 1.14 | [1.09; 1.19] | 8.1e−09 | 2,025 | 45,705 |
| +sex,smoking,ethnicity | 1.11 | [1.07; 1.16] | 1.2e−06 | 2,017 | 45,549 |
| +Height,Weight,SphericalEquivalent | 1.12 | [1.07; 1.18] | 1.1e−06 | 1,746 | 39,188 |
| +CAD,HbA1c,SBP,DBP | 1.10 | [1.04; 1.15] | 0.0003 | 1,561 | 35,887 |
| **RNFL** | | | | | |
| Unadjusted | 1.24 | [1.19; 1.29] | 1.6e−21 | 2,025 | 45,705 |
| +age,age^2 | 1.13 | [1.08; 1.18] | 3e−08 | 2,025 | 45,705 |
| +sex,smoking,ethnicity | 1.09 | [1.05; 1.14] | 6.5e−05 | 2,017 | 45,549 |
| +Height,Weight,SphericalEquivalent | 1.09 | [1.04; 1.15] | 0.00037 | 1,746 | 39,188 |
| +CAD,HbA1c,SBP,DBP | 1.07 | [1.01; 1.12] | 0.015 | 1,561 | 35,887 |
| **GCL** | | | | | |
| Unadjusted | 1.23 | [1.18; 1.29] | 2.5e−21 | 2,025 | 45,705 |
| +age,age^2 | 1.10 | [1.06; 1.15] | 9.9e−06 | 2,025 | 45,705 |
| +sex,smoking,ethnicity | 1.10 | [1.05; 1.14] | 4.9e−05 | 2,017 | 45,549 |
| +Height,Weight,SphericalEquivalent | 1.12 | [1.07; 1.18] | 5.4e−06 | 1,746 | 39,188 |
| +CAD,HbA1c,SBP,DBP | 1.10 | [1.04; 1.16] | 0.00033 | 1,561 | 35,887 |
| **IPL** | | | | | |
| Unadjusted | 1.21 | [1.16; 1.26] | 5.1e−17 | 2,025 | 45,705 |
| +age,age^2 | 1.08 | [1.03; 1.13] | 0.00053 | 2,025 | 45,705 |
| +sex,smoking,ethnicity | 1.09 | [1.04; 1.14] | 0.00012 | 2,017 | 45,549 |
| +Height,Weight,SphericalEquivalent | 1.12 | [1.07; 1.18] | 5.1e−06 | 1,746 | 39,188 |
| +CAD,HbA1c,SBP,DBP | 1.10 | [1.04; 1.16] | 0.00039 | 1,561 | 35,887 |
| **INL** | | | | | |
| Unadjusted | 1.02 | [0.98; 1.07] | 0.34 | 2,025 | 45,705 |
| +age,age^2 | 0.97 | [0.93; 1.01] | 0.18 | 2,025 | 45,705 |
| +sex,smoking,ethnicity | 1.00 | [0.96; 1.04] | 0.97 | 2,017 | 45,549 |
| +Height,Weight,SphericalEquivalent | 1.01 | [0.96; 1.06] | 0.64 | 1,746 | 39,188 |
| +CAD,HbA1c,SBP,DBP | 1.02 | [0.96; 1.07] | 0.55 | 1,561 | 35,887 |
| **OPL+ONL** | | | | | |
| Unadjusted | 1.02 | [0.98; 1.07] | 0.37 | 2,025 | 45,705 |
| +age,age^2 | 1.01 | [0.97; 1.06] | 0.58 | 2,025 | 45,705 |
| +sex,smoking,ethnicity | 1.03 | [0.99; 1.08] | 0.15 | 2,017 | 45,549 |
| +Height,Weight,SphericalEquivalent | 1.03 | [0.98; 1.08] | 0.19 | 1,746 | 39,188 |
| +CAD,HbA1c,SBP,DBP | 1.03 | [0.98; 1.09] | 0.25 | 1,561 | 35,887 |
| **PS** | | | | | |
| Unadjusted | 1.39 | [1.33; 1.44] | 9.9e−53 | 2,025 | 45,705 |
| +age,age^2 | 1.19 | [1.14; 1.24] | 1e−14 | 2,025 | 45,705 |
| +sex,smoking,ethnicity | 1.17 | [1.12; 1.23] | 1.1e−12 | 2,017 | 45,549 |
| +Height,Weight,SphericalEquivalent | 1.16 | [1.11; 1.22] | 7.5e−10 | 1,746 | 39,188 |
| +CAD,HbA1c,SBP,DBP | 1.14 | [1.08; 1.20] | 1.2e−06 | 1,561 | 35,887 |
| **RPE+BM** | | | | | |
| Unadjusted | 1.01 | [0.97; 1.06] | 0.6 | 2,025 | 45,705 |
| +age,age^2 | 1.04 | [0.99; 1.08] | 0.13 | 2,025 | 45,705 |
| +sex,smoking,ethnicity | 1.06 | [1.01; 1.11] | 0.011 | 2,017 | 45,549 |
| +Height,Weight,SphericalEquivalent | 1.06 | [1.01; 1.11] | 0.027 | 1,746 | 39,188 |
| +CAD,HbA1c,SBP,DBP | 1.07 | [1.01; 1.13] | 0.018 | 1,561 | 35,887 |
| **CSI** | | | | | |
| Unadjusted | 1.22 | [1.17; 1.28] | 9.2e−20 | 2,025 | 45,705 |
| +age,age^2 | 1.02 | [0.97; 1.06] | 0.44 | 2,025 | 45,705 |
| +sex,smoking,ethnicity | 1.02 | [0.98; 1.07] | 0.33 | 2,017 | 45,549 |
| +Height,Weight,SphericalEquivalent | 1.02 | [0.97; 1.07] | 0.53 | 1,746 | 39,188 |
| +CAD,HbA1c,SBP,DBP | 1.01 | [0.96; 1.07] | 0.75 | 1,561 | 35,887 |

**Supplementary Figure 5:** Associations of retinal layer thickness with incident mortality using different covariates in a Cox survival model.

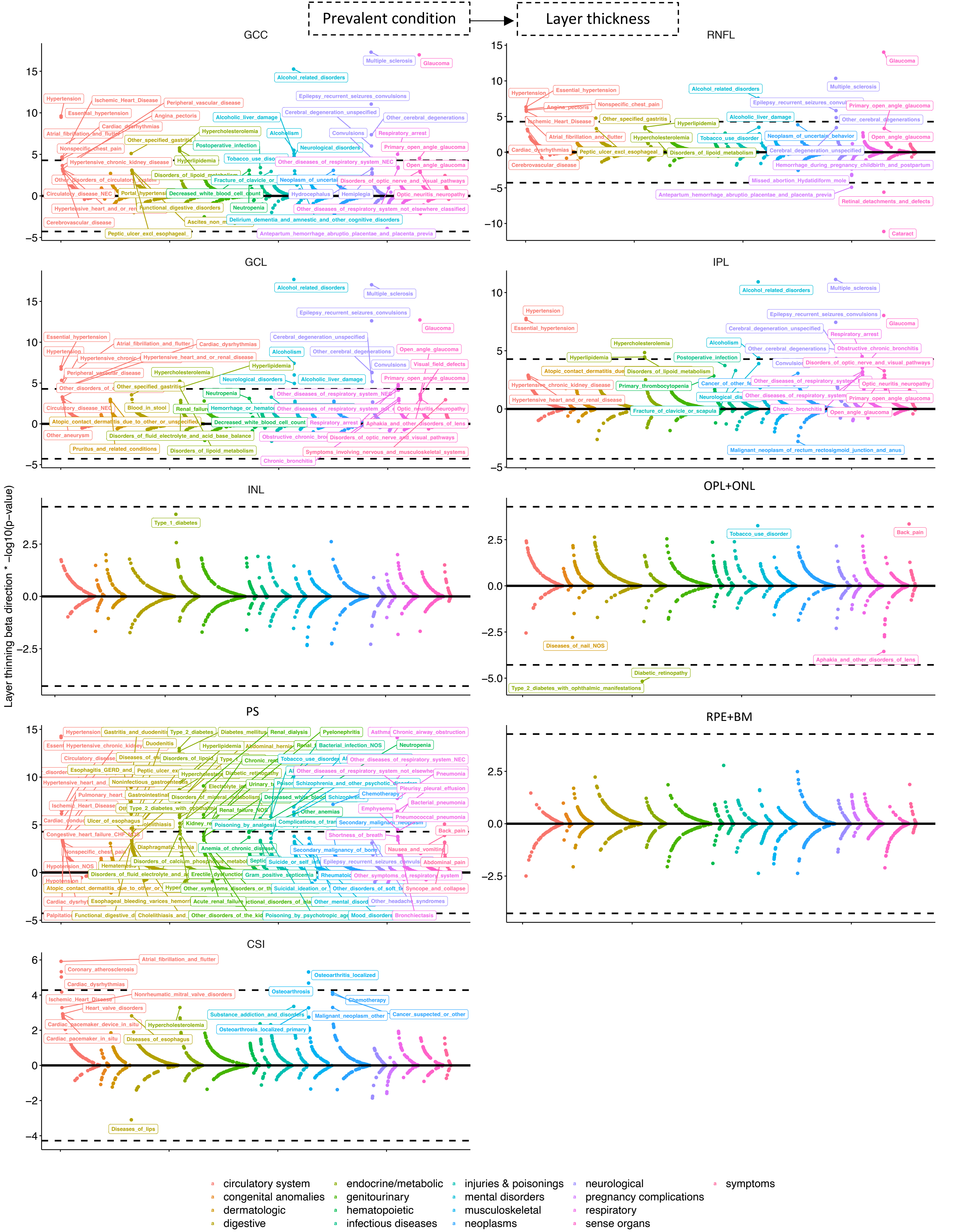


**Supplementary Figure 6:** Associations of prevalent conditions with retinal layer thickness. Plotted are prevalent conditions ordered by system and p-value of significance (x-axis) by the –log10(p-value) x layer thinning beta direction (whereby >0 on the y-axis reflects association with retinal layer thinning). Labeled are prevalent conditions colored by relevant organ system which are significant using an FDR<0.05 threshold. Dotted line reflects the Bonferroni threshold for significance for each layer for reference.

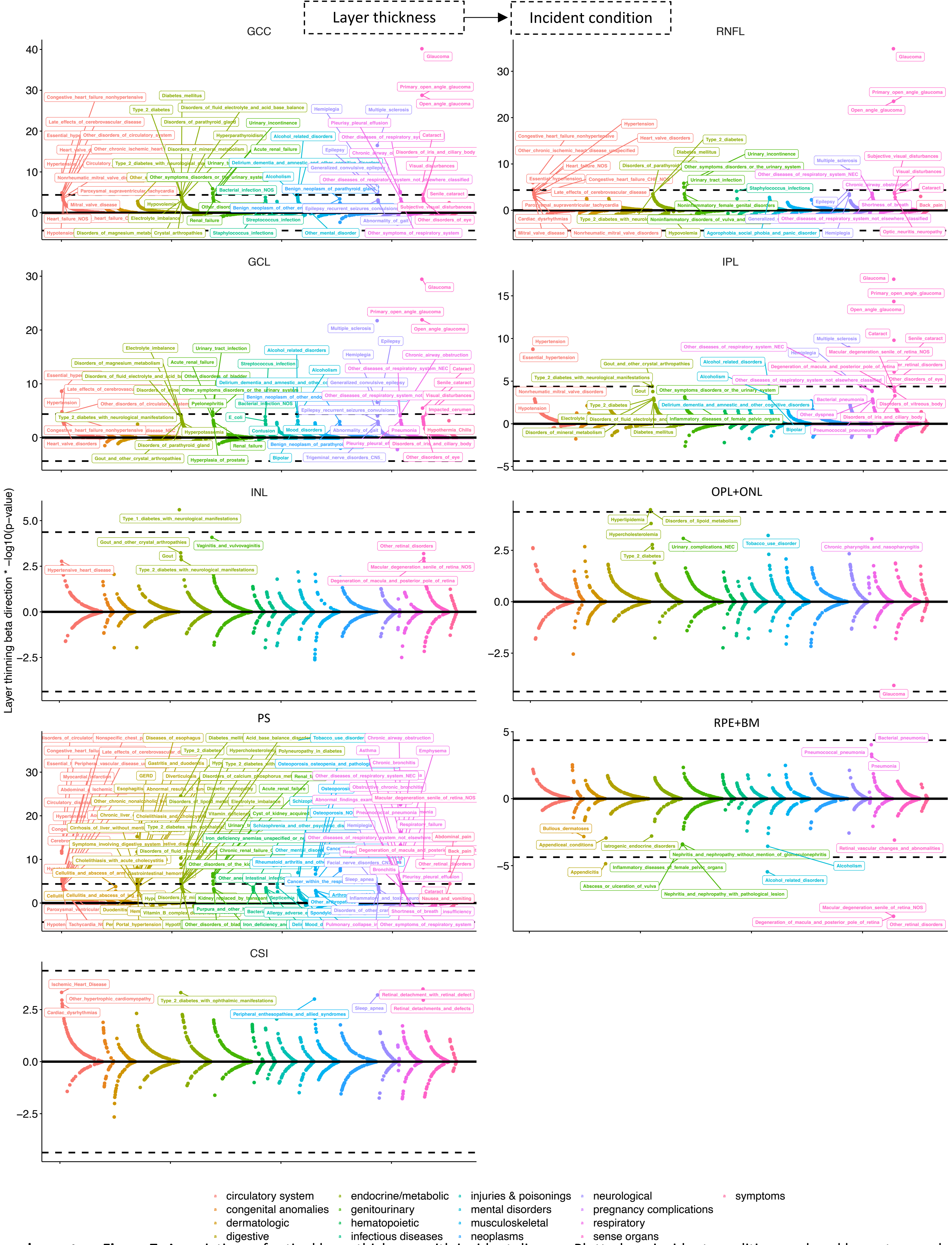


**Supplementary Figure 7:** Associations of retinal layer thickness with incident disease. Plotted are incident conditions ordered by system and p-value of significance (x-axis) by the –log10(p-value) x layer thinning beta direction (whereby >0 on the y-axis reflects association with retinal layer thinning). Labeled are incident conditions which are significant using an FDR<0.05 threshold. Dotted line reflects the Bonferroni threshold for significance for each layer for reference.

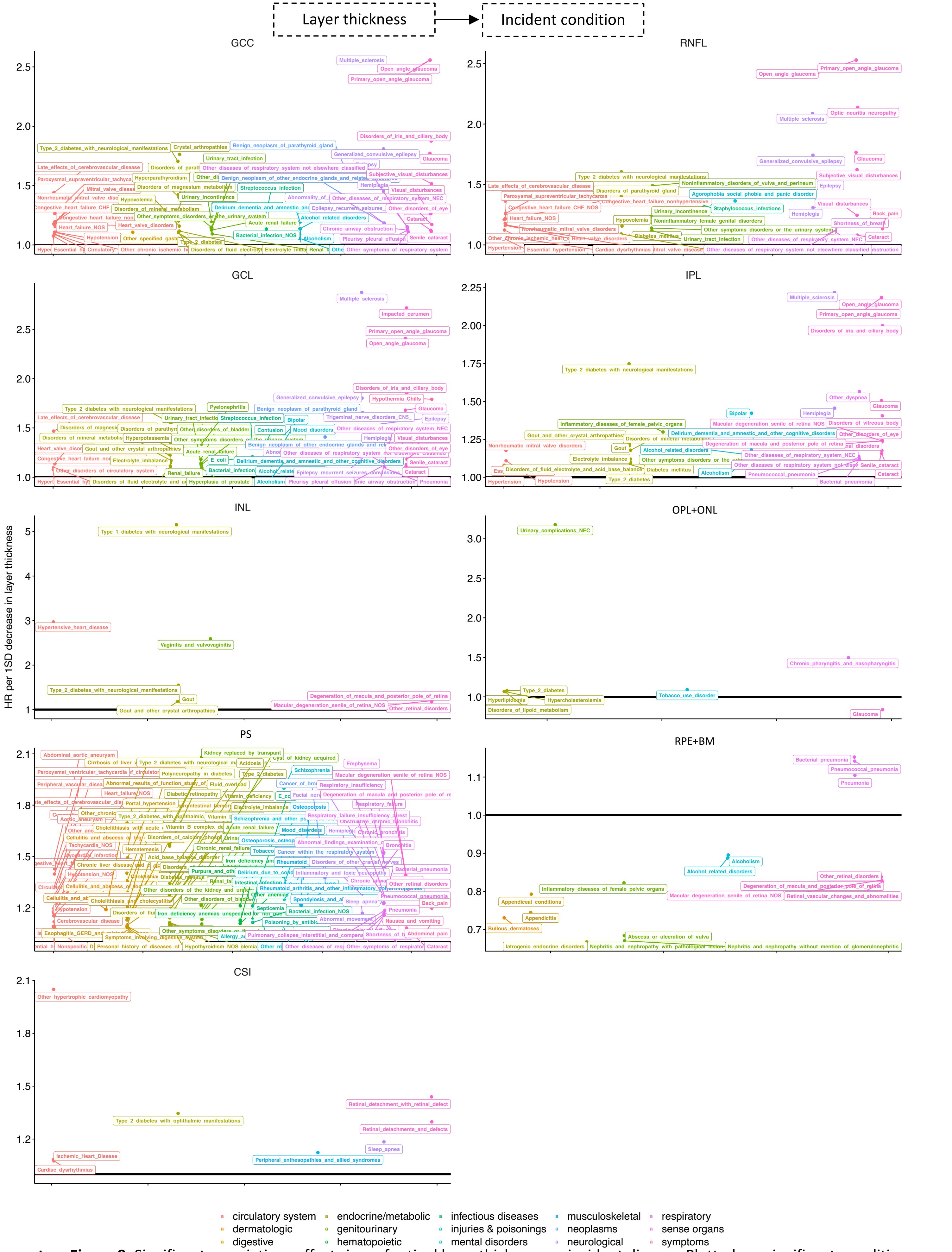


**Supplementary Figure 8:** Significant associations effect sizes of retinal layer thickness on incident disease. Plotted are significant conditions ordered by system and p-value of significance (x-axis) by the HR per 1 SD decrease in layer thickness. Labeled are incident conditions which are significant using an FDR<0.05 threshold.

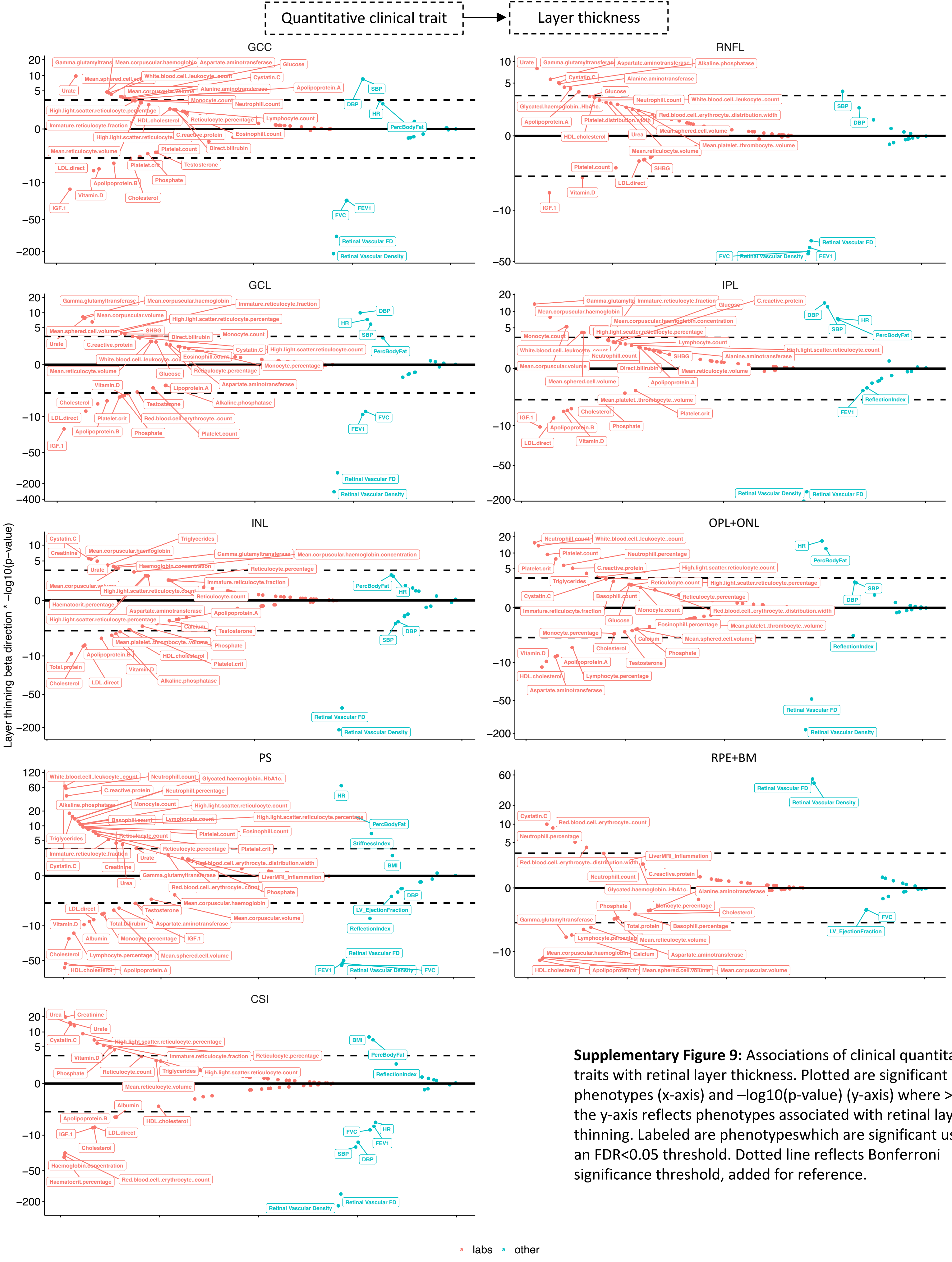


**Supplementary Figure 9:** Associations of clinical quantitative traits with retinal layer thickness. Plotted are significant phenotypes (x-axis) and –log10(p-value) (y-axis) where >0 on the y-axis reflects phenotypes associated with retinal layer thinning. Labeled are phenotypeswhich are significant using an FDR<0.05 threshold. Dotted line reflects Bonferroni significance threshold, added for reference.

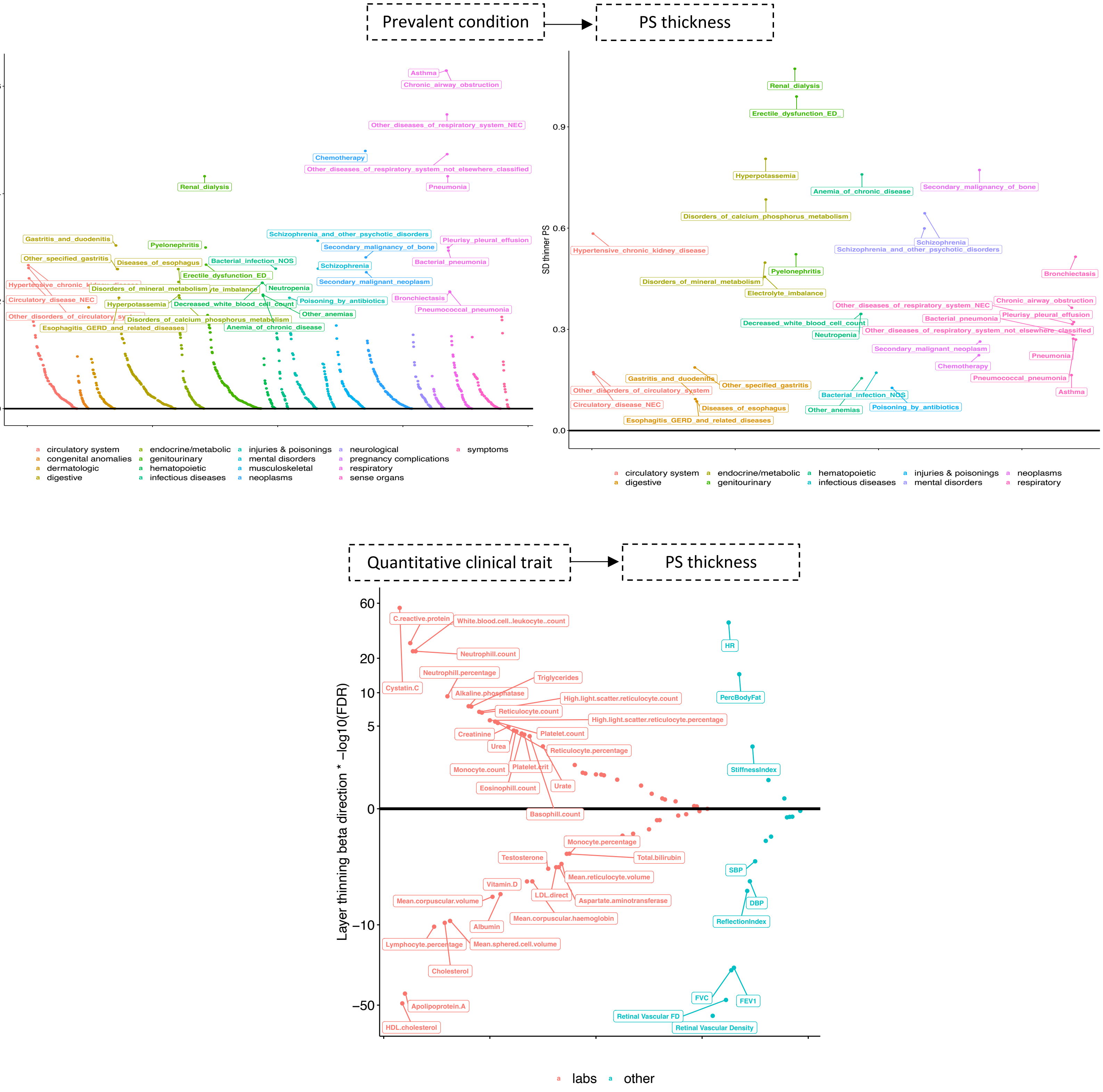


**Supplementary Figure 10:** Phenotypic associations with photoreceptor segment (PS) layer thickness after additionally adjusting for hypertension (incident or prevalent combined), type 2 diabetes (incident or prevalent combined), HbA1c, and a 25-factor smoking covariate. Plotted are significant phenotypes (x-axis) and –log10(False Discovery Rate, FDR) (y-axis). Labeled are phenotypes which are significant using an FDR<0.01 threshold. **a.** Association of prevalent disease with PS thickness, **b.** Association of Quantitiative clinical traits with PS thickness.

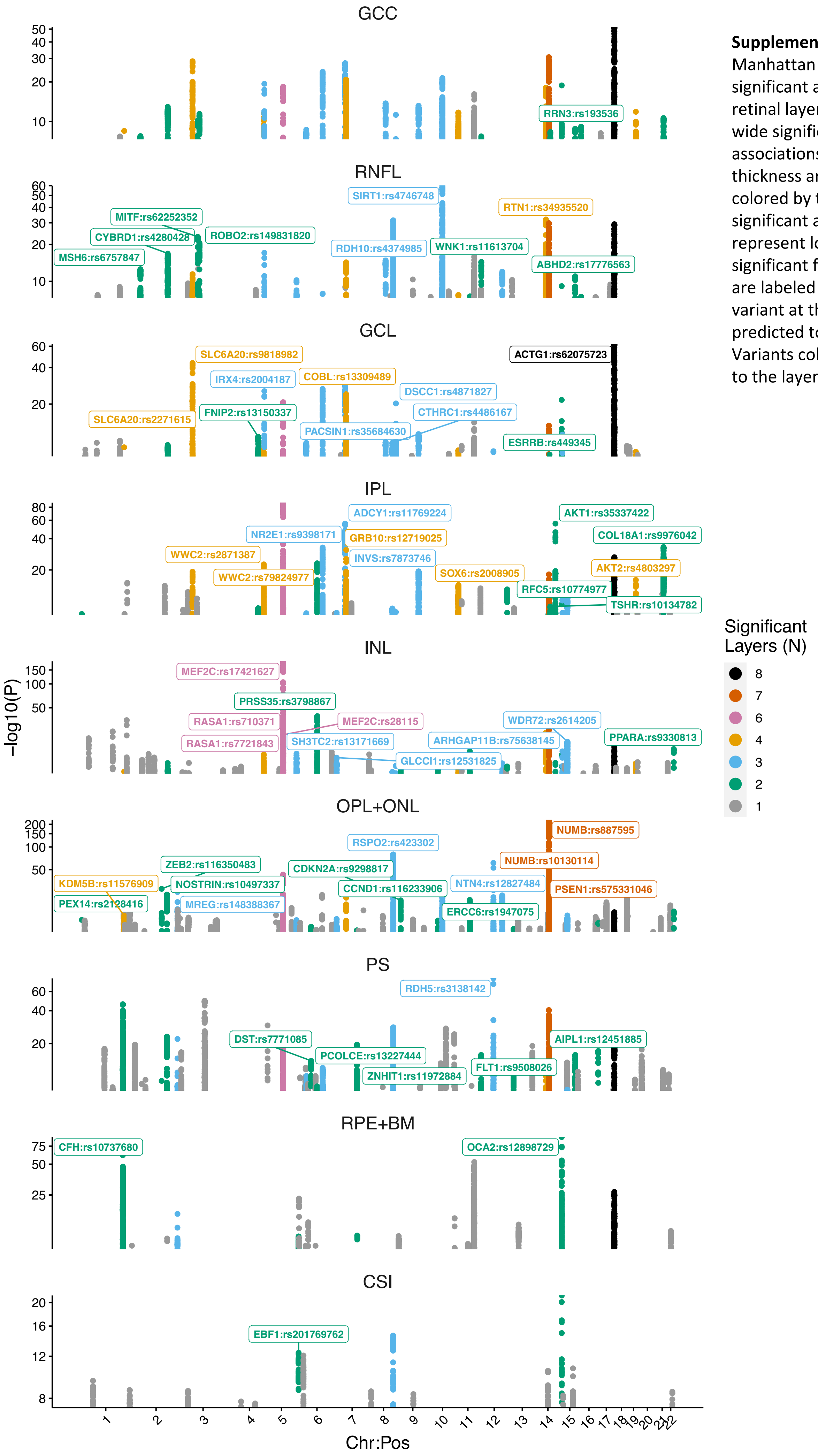


**Supplementary Figure 11:** Manhattan plot of genome-wide significant association results for the retinal layer thicknesses. Genome-wide significant ($P<5x10^{-8}$) associations with retinal layer thickness are provided by layer, colored by the number of layers significant at that locus. Labeled loci represent loci genome-wide significant for at least two layers and are labeled with the most significant variant at that locus and the predicted top PoPS gene within 1MB. Variants colored as grey are unique to the layer of interest.

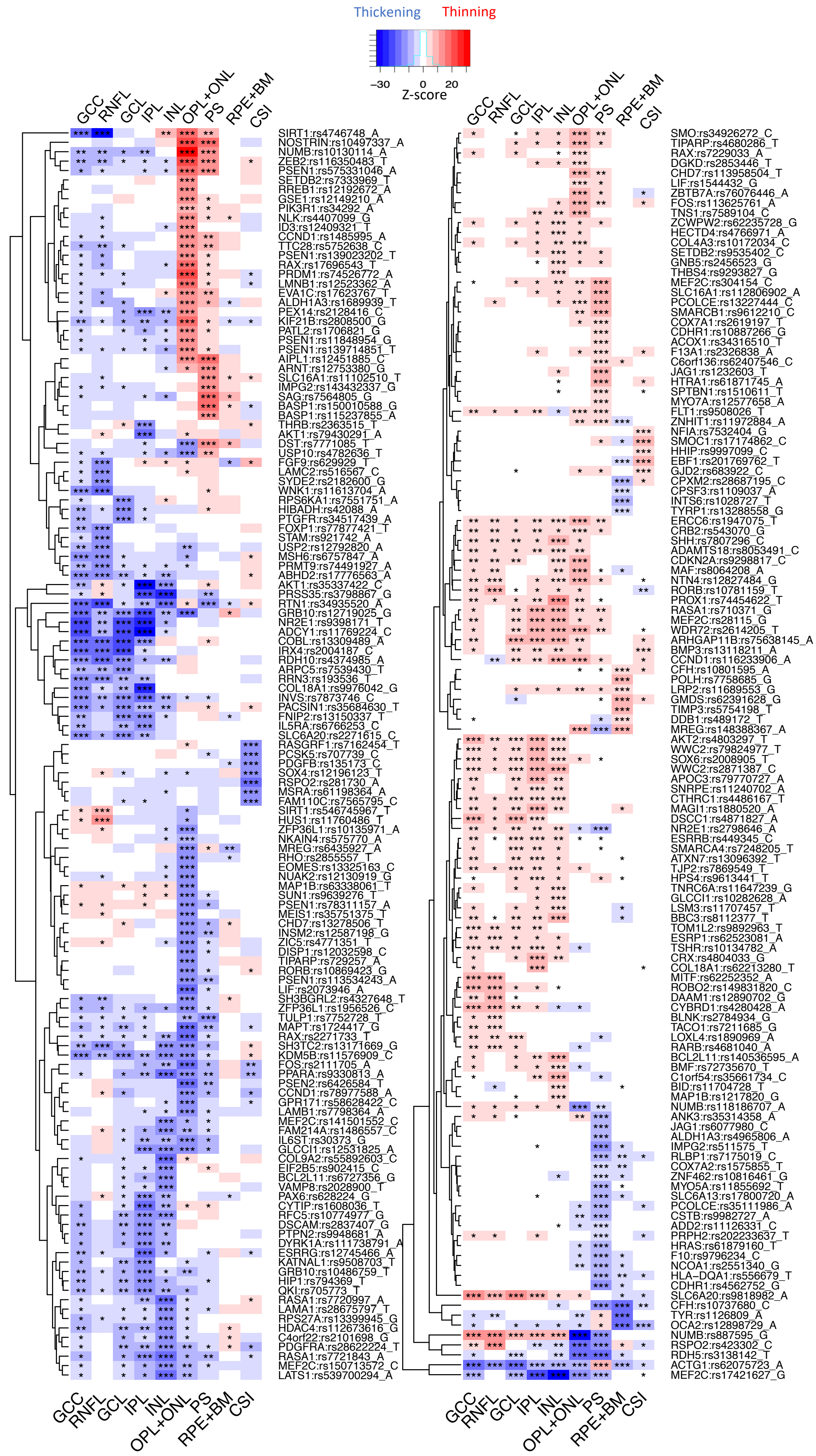


**Supplementary Figure 12:** Relative association effects of genome-wide significant association results compared by retinal layer. Heat maps comparing the 259 unique genome-wide significant ($P<5x10^{-8}$), independent (LD clumping $r^2<0.1$), associations across retinal layers.Color reflects the z-score (beta/se) of association, with red reflecting layer thinning associated with the indicated allele and blue representing layer thickening. Labeled loci are labeled with the predicted top PoPS gene within 1MB of the given variant and the effect allele used for plotting the z-score. Within cells, association strength is denoted as such: ***$P<5x10^{-8}$, **$5x10^{-8}\leq P<0.0001$, *$0.0001\leq P<0.05$. Full results are provided in **Supplementary Table 7**.

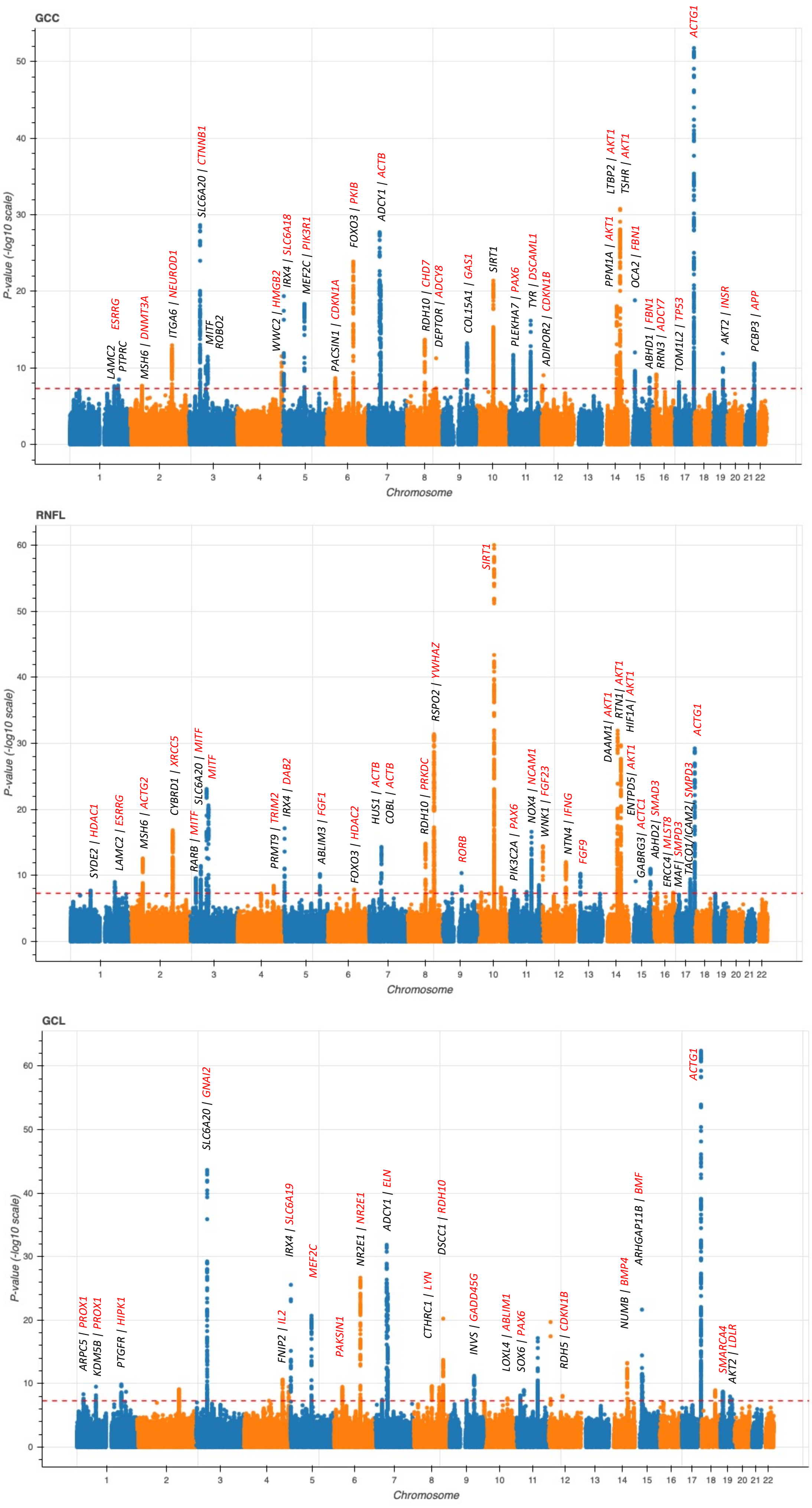


**Supplementary Figure 13:** Genome-wide association results plotted by layer for GCC, RNFL, and GCL through Manhattan plots. Predicted genes at each loci are labeled using top PoPS locus within 1MB (black)and 50MB (red) of the top variant as annotated in Supplementary Table 7.

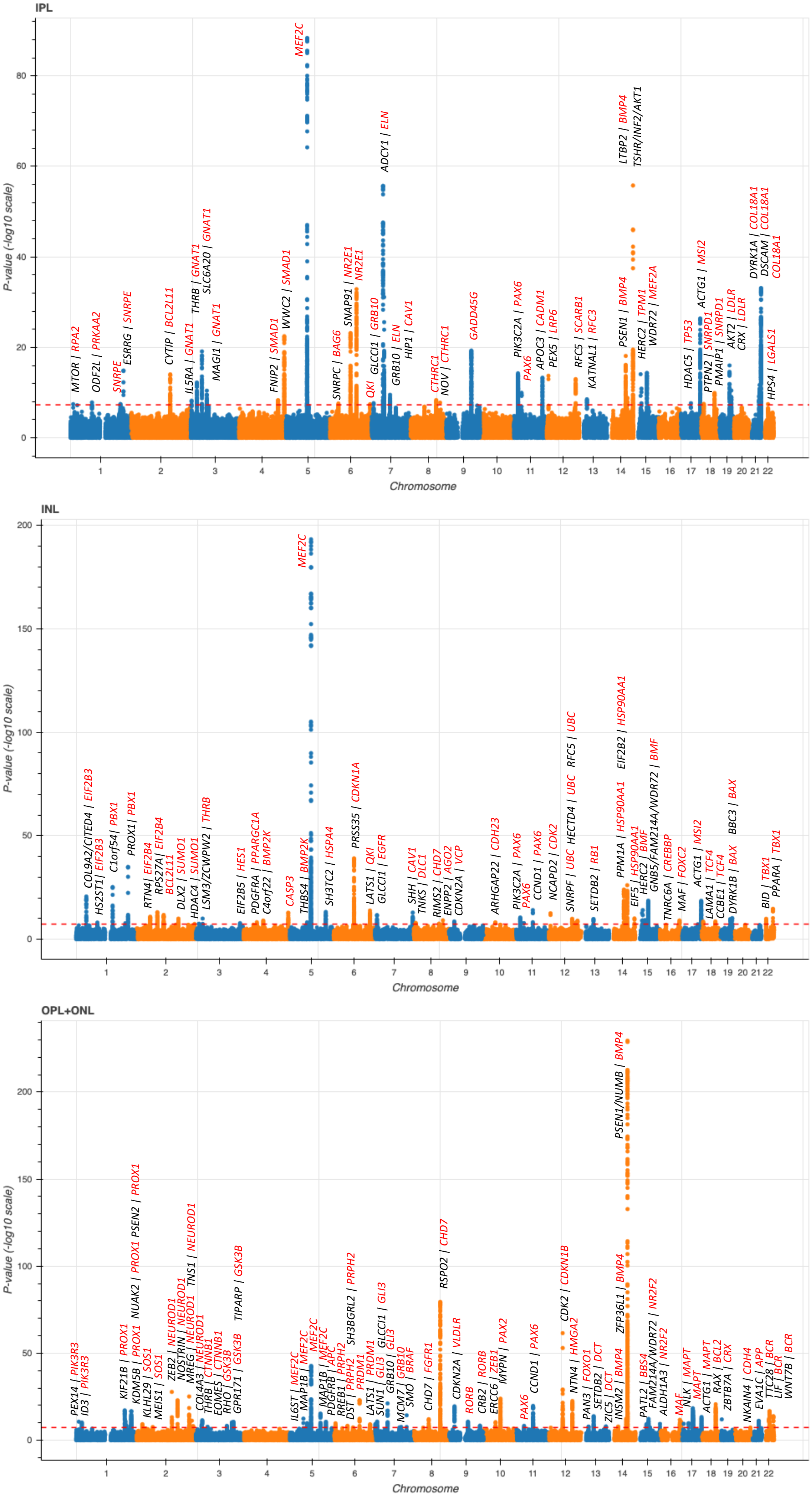


**Supplementary Figure 14:** Genome-wide association results plotted by layer for IPL, INL, and OPL+ONL through Manhattan plots. Predicted genes at each loci are labeled using top PoPS locus within 1MB (black)and 50MB (red) of the top variant as annotated in Supplementary Table 7.

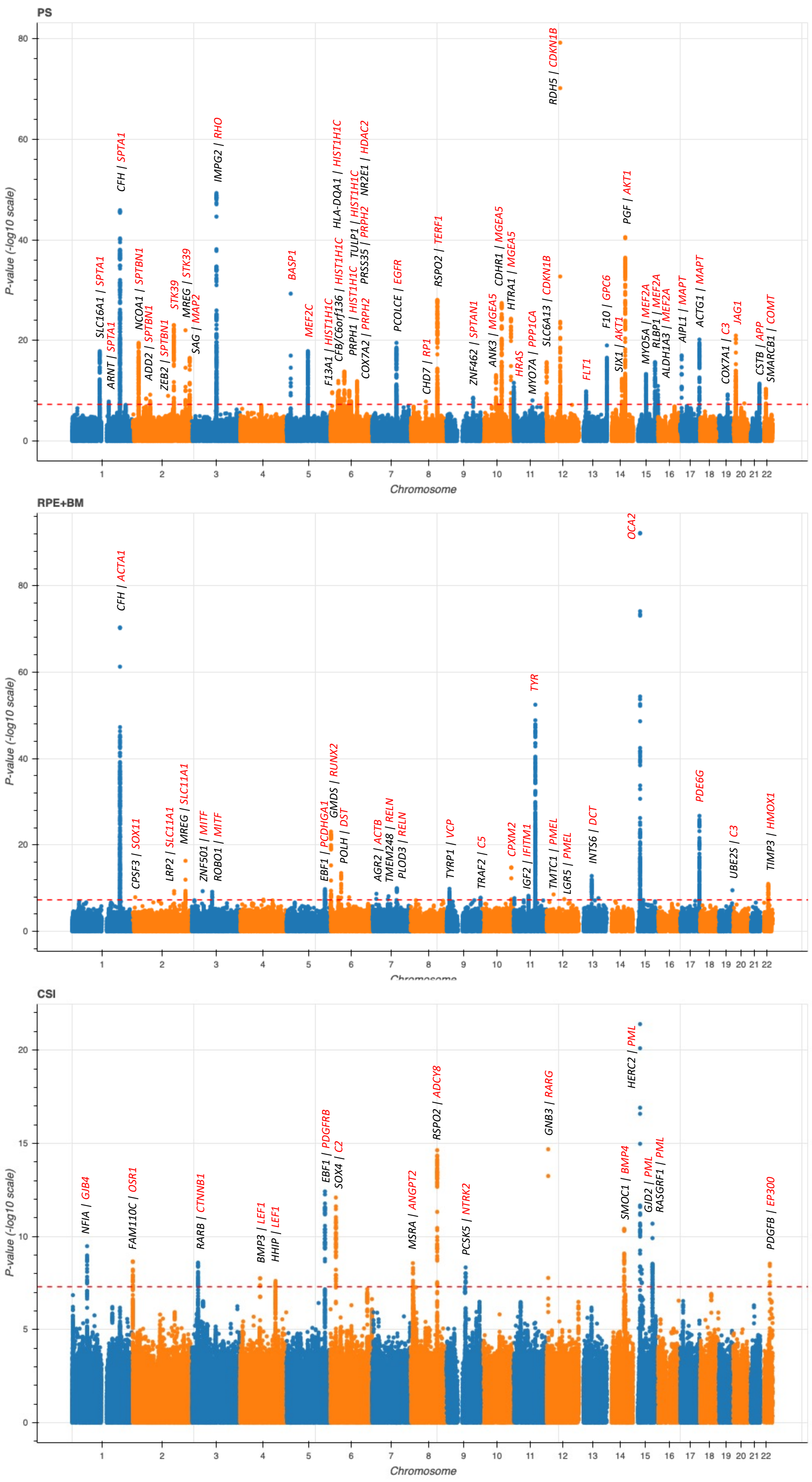


**Supplementary Figure 15:** Genome-wide association results plotted by layer for PS, RPE+BM, and CSI through Manhattan plots. Predicted genes at each loci are labeled using top PoPS locus within 1MB (black)and 50MB (red) of the top variant as annotated in Supplementary Table 7.

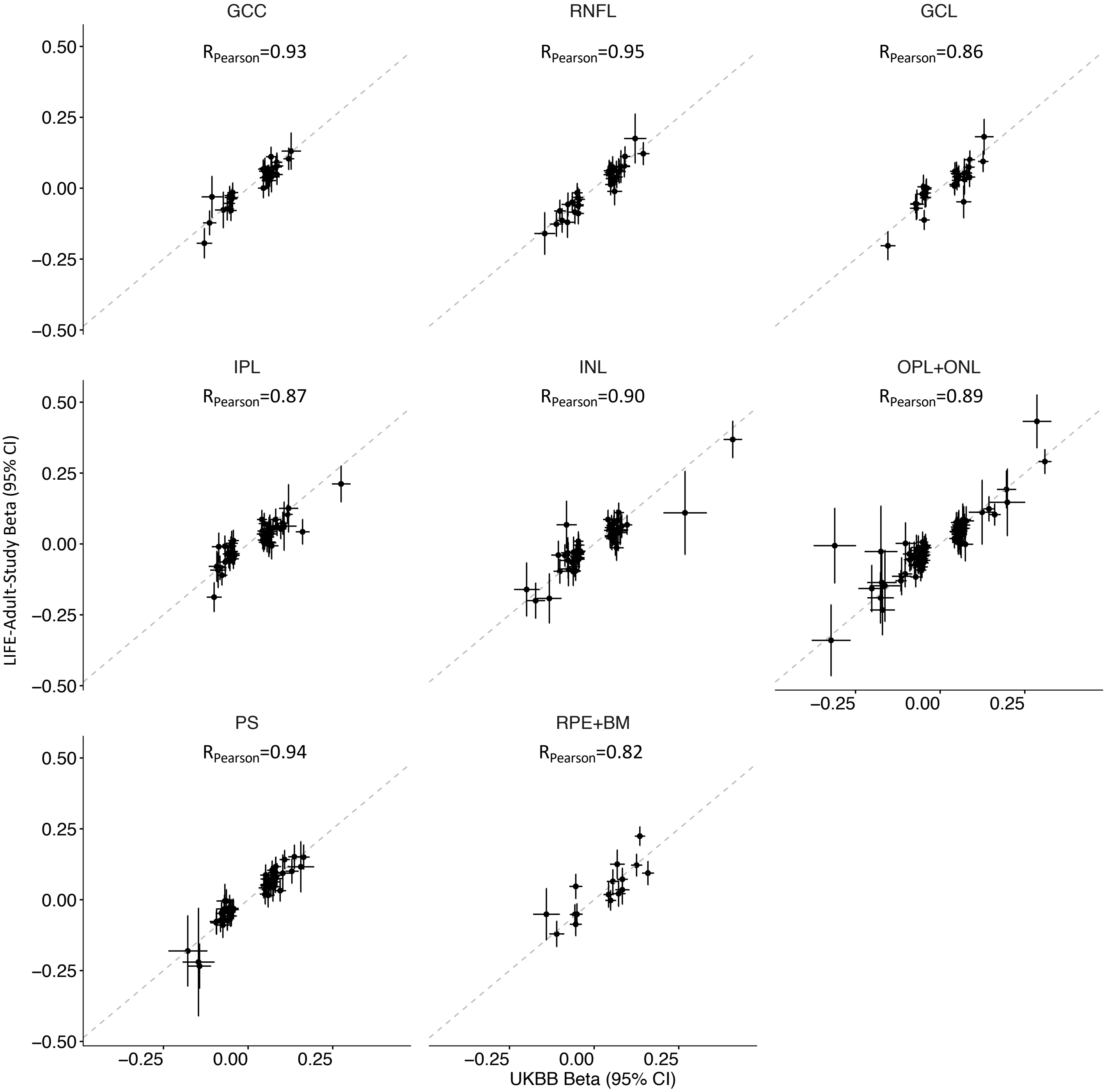


**Supplementary Figure 16:** Replication of GWAS summary statistics in the LIFE-Adult-Study cohort. Correlation of UKBB versus LIFE-Adult-Study beta and 95% CI across top 259 independent, genome-wide significant variants identified in UKBB plotted by layer. The dotted abline reflects x=y. Pearson correlations by layer are labeled. Of note, CSI was not available in the LIFE-Adult-Study cohort, limiting replication.

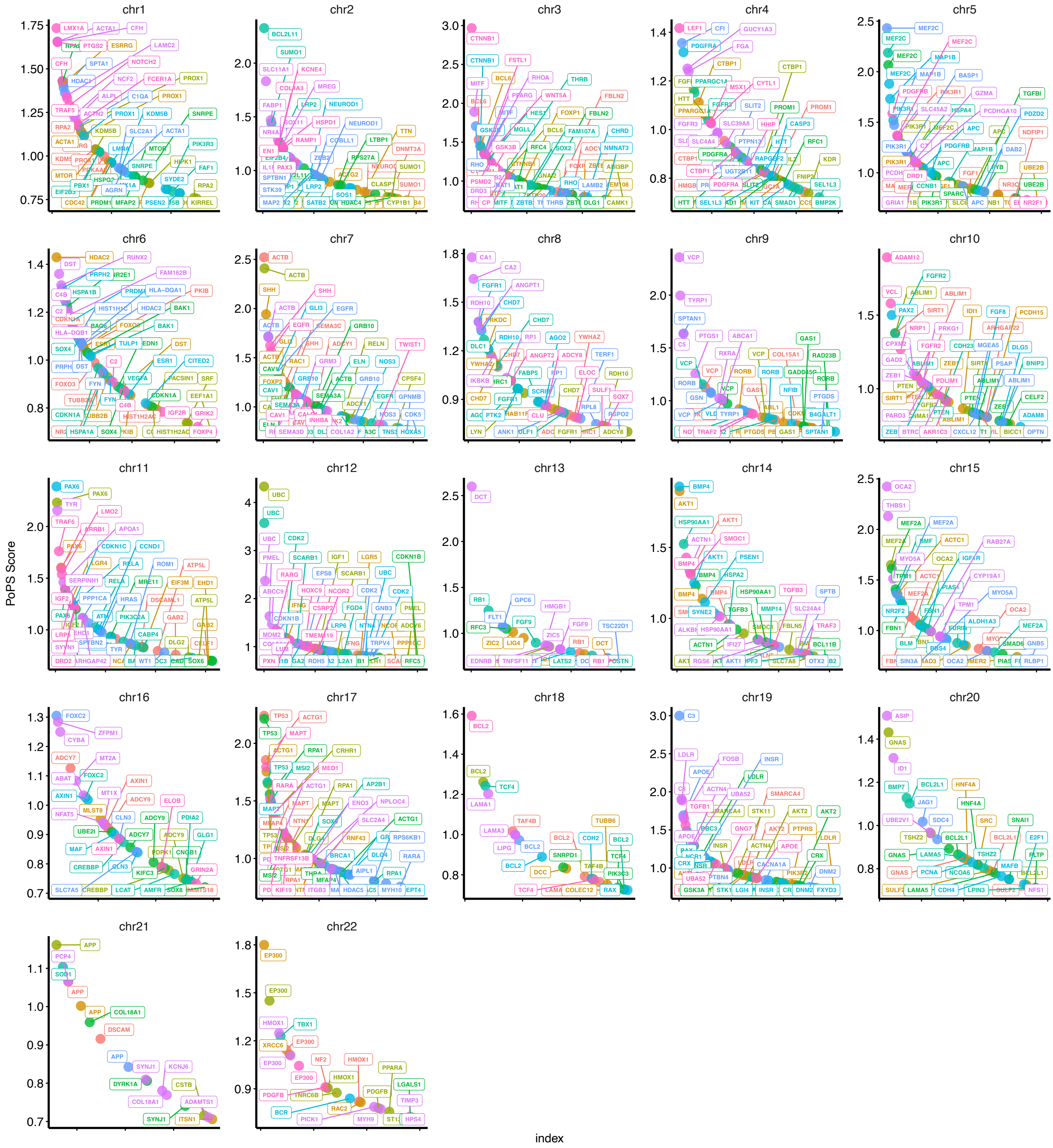


**Supplementary Figure 17:** Top POPs genes within 1MB of a top variant by chromosome colored by retinal layer

**Supplementary Figure 18:** Top POPs genes by chromosome and retinal layer for the GCC, RNFL, and GCL layers.

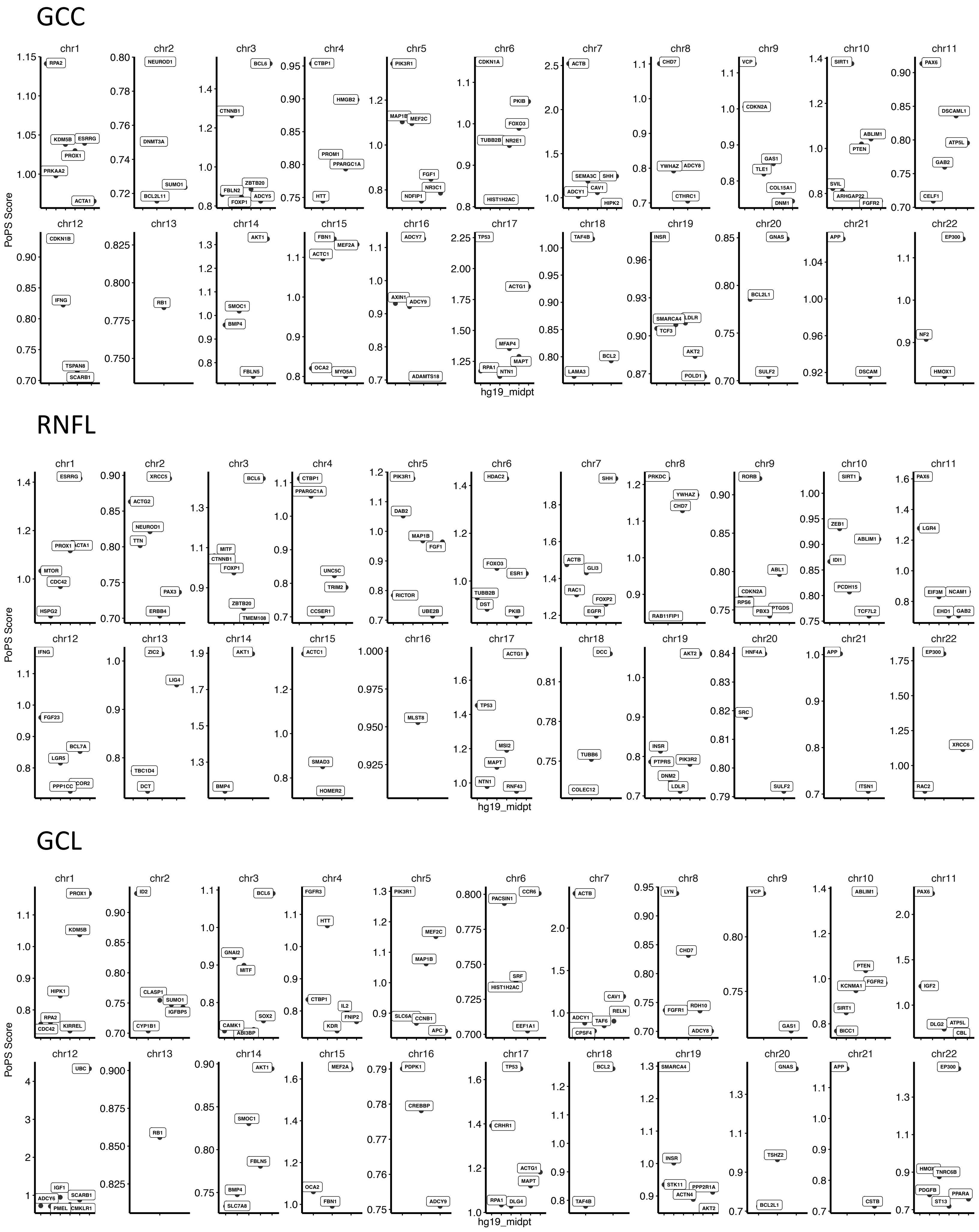

**Supplementary Figure 19:** Top POPs genes by chromosome and retinal layer for the INL, IPL, and ONL+OPL layers.

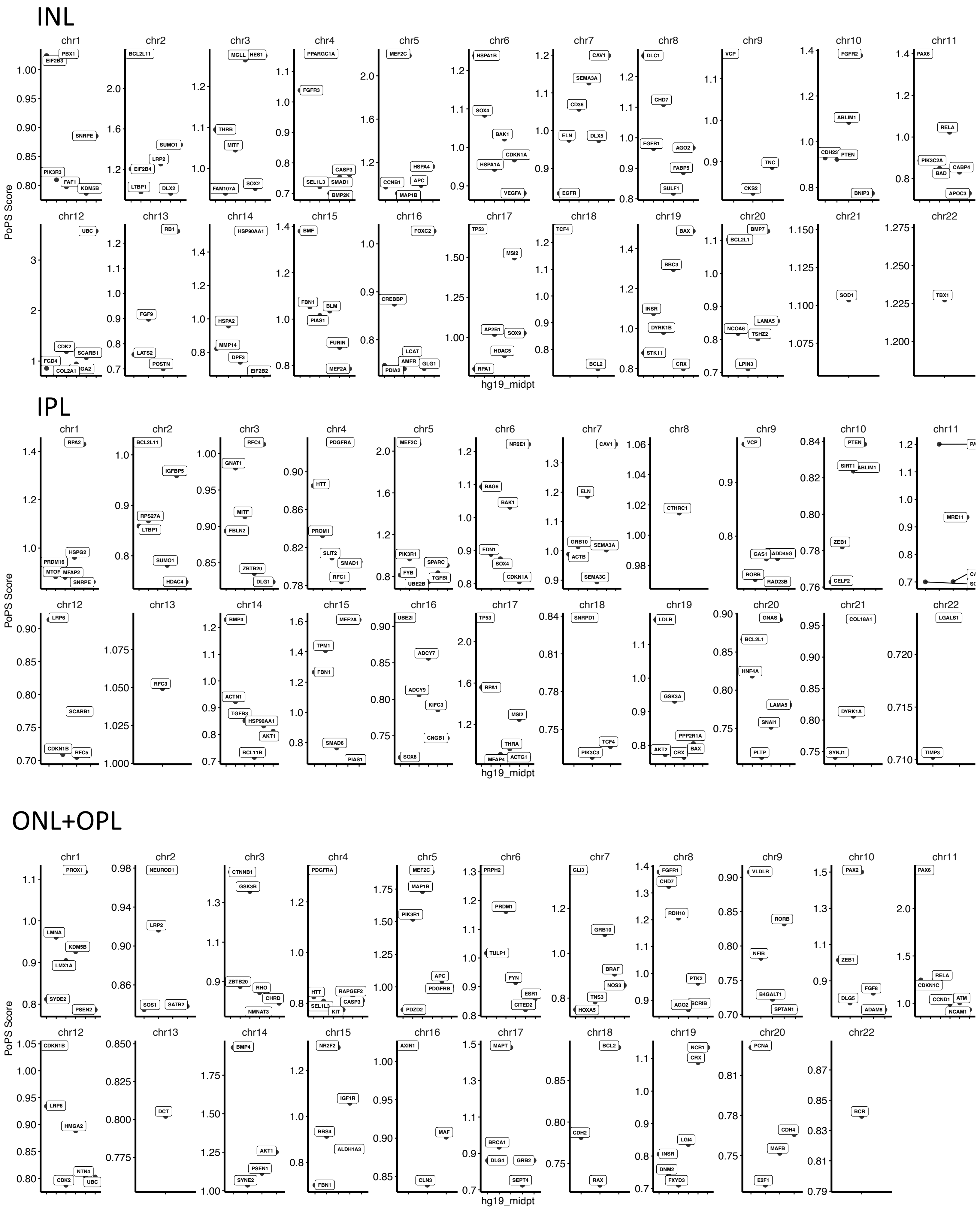

**Supplementary Figure 20:** Top POPs genes by chromosome and retinal layer for the PS, RPE+BM, and CSI layers.

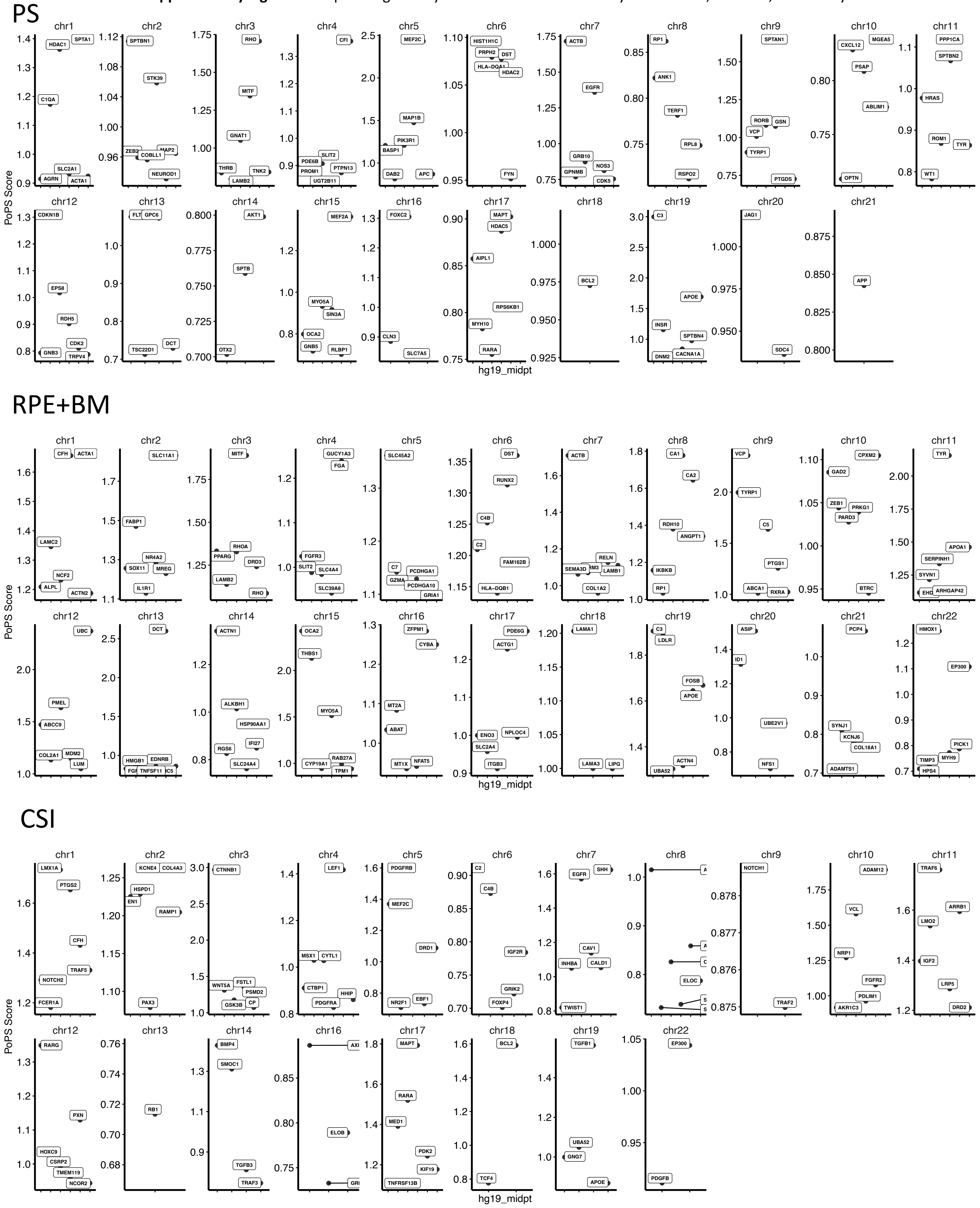

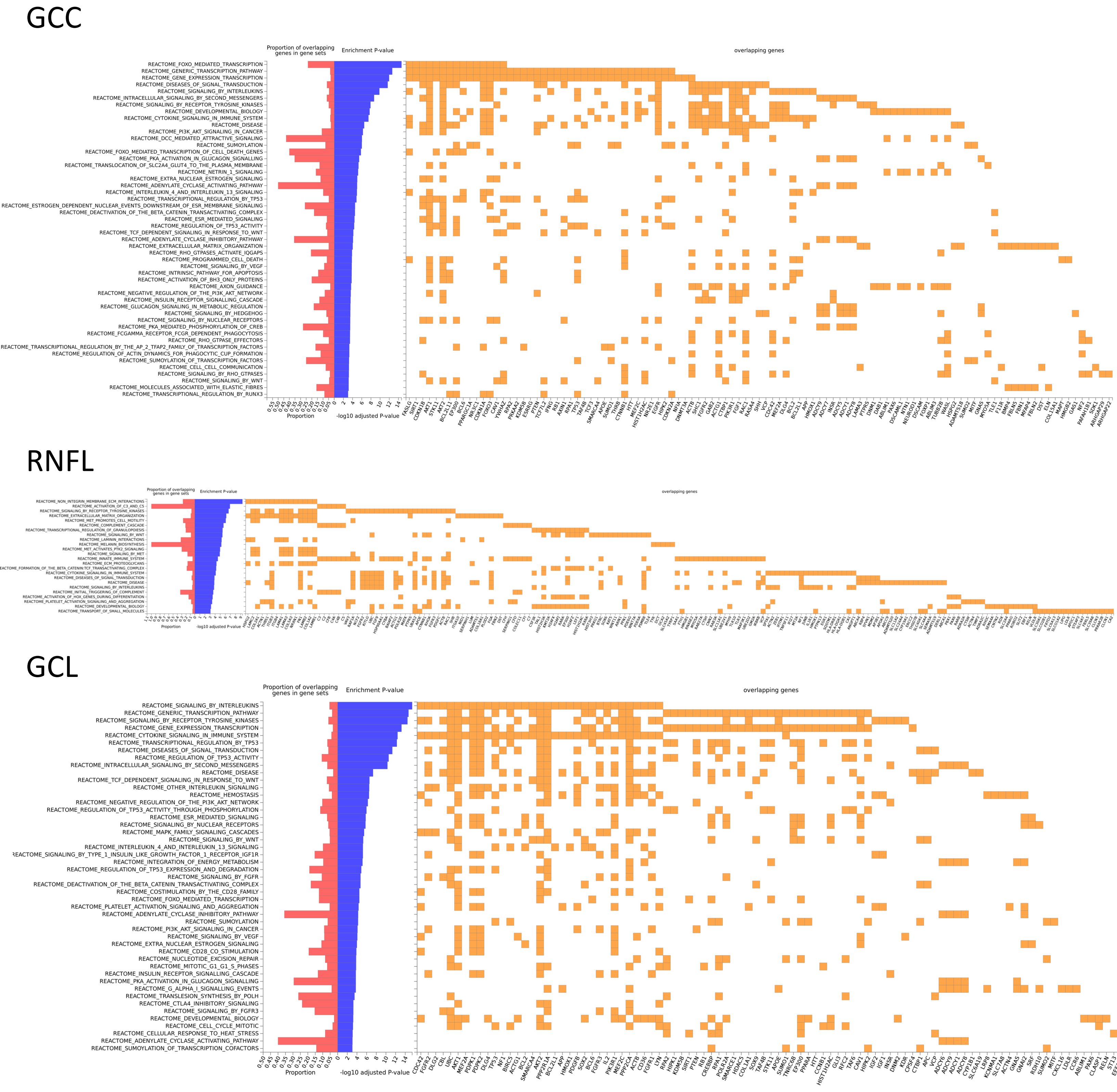


**Supplementary Figure 21:** Reactome gene set enrichment analysis analysis using POPs genes with score > 1 across the GCC, RNFL, and GCL layers.

**Supplementary Figure 22:** Reactome gene set enrichment analysis analysis using POPs genes with score > 1 across the INL, IPL, and ONL+OPL layers.

INL

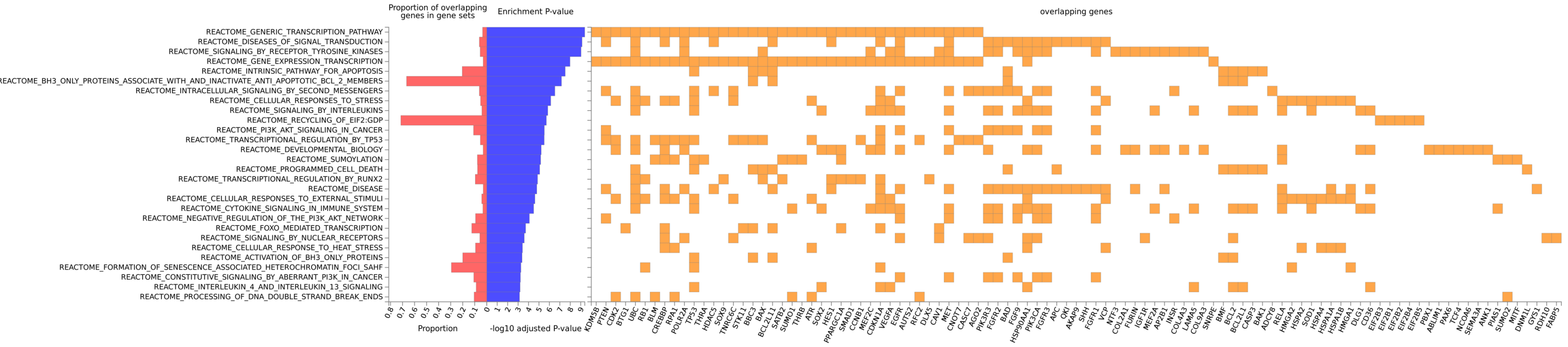


IPL

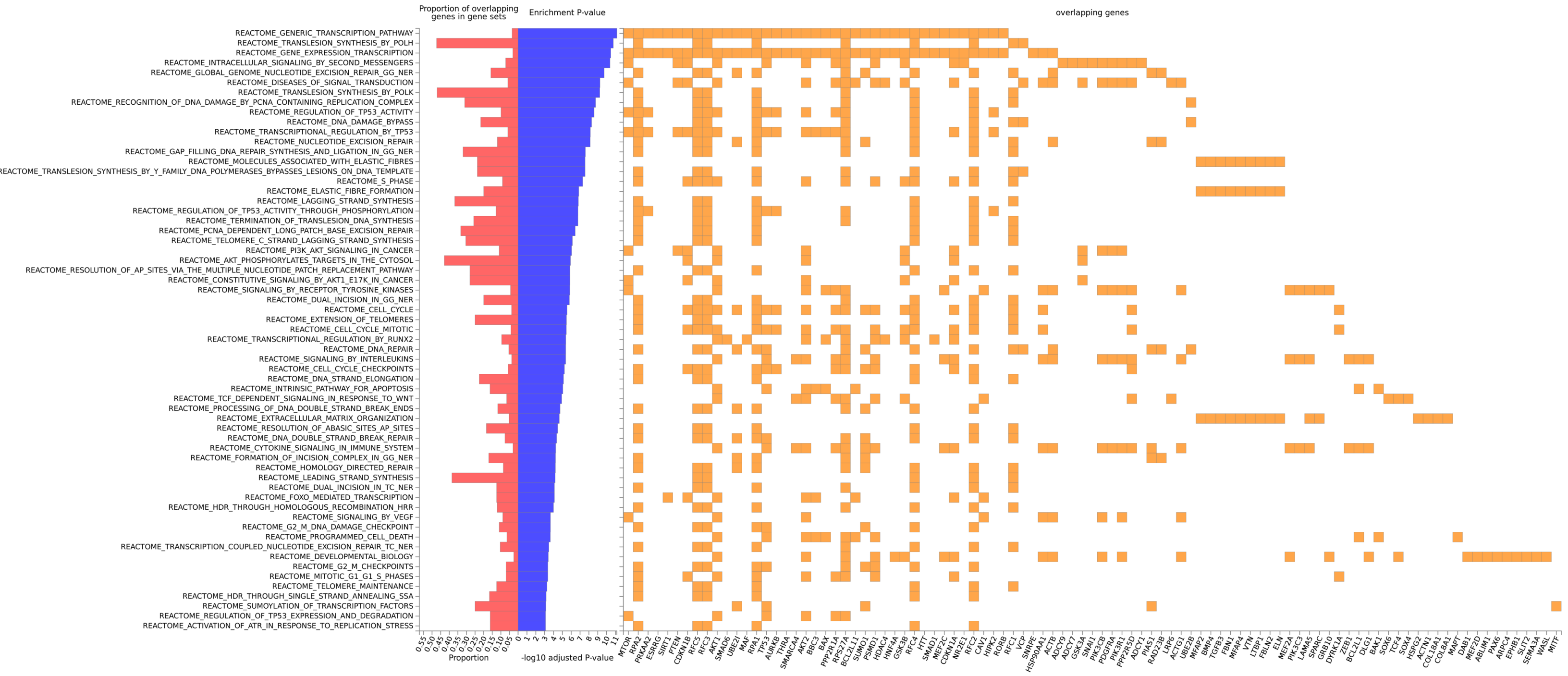


ONL+OPL

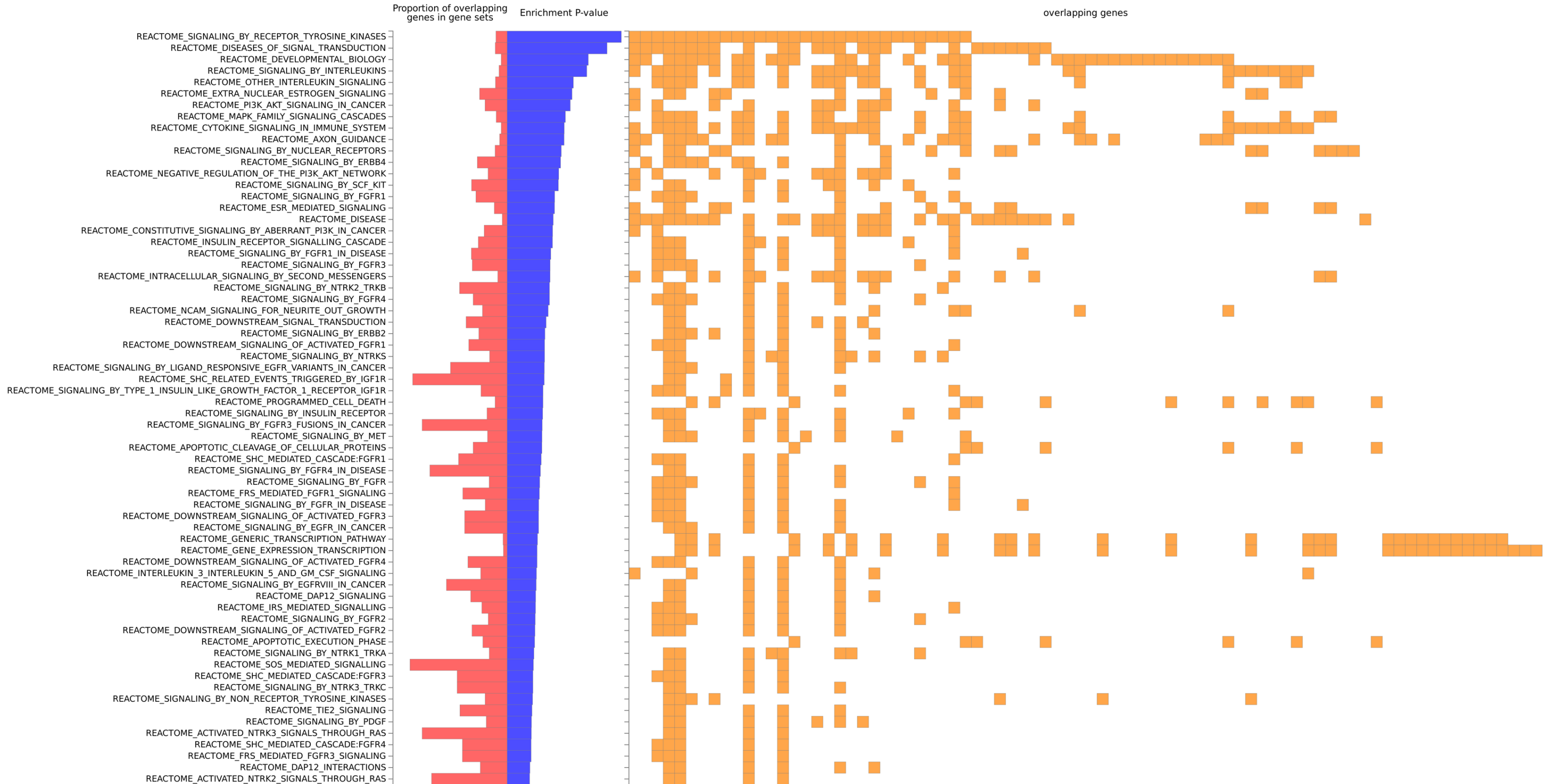

# PS

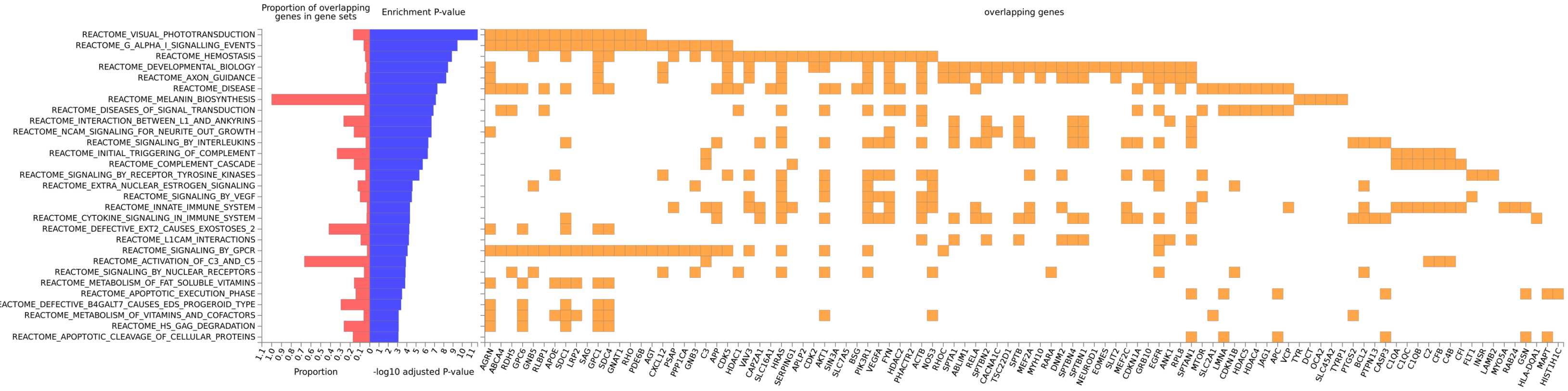


### RPE

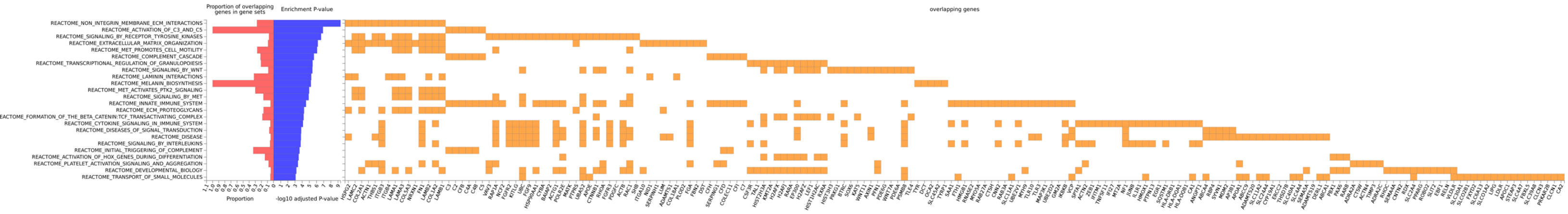


### CSI

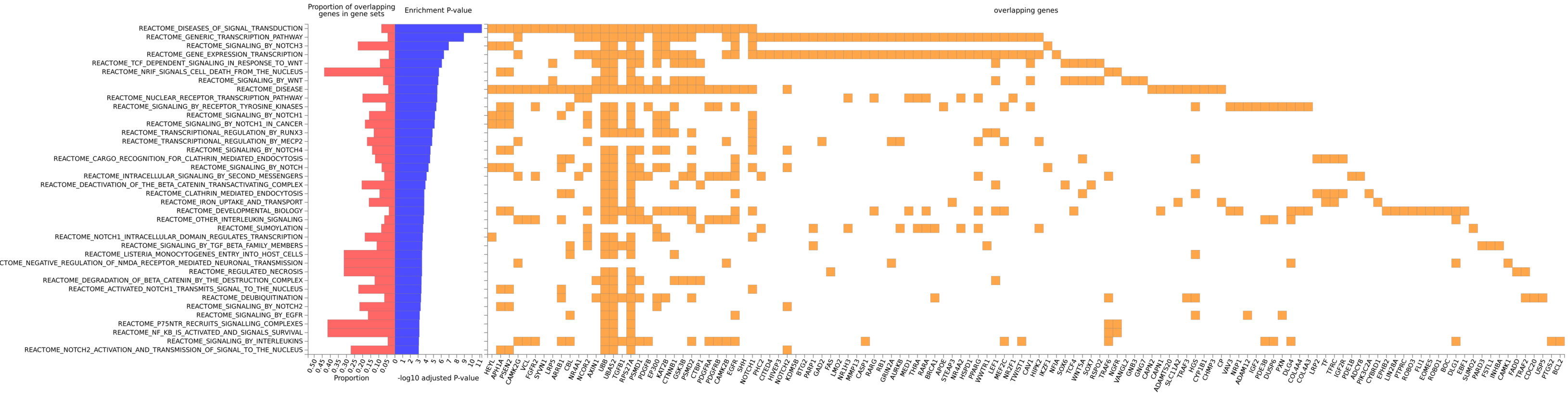


**Supplementary Figure 23:** Reactome gene set enrichment analysis analysis using POPs genes with score > 1 across the PS, RPE, and CSI layers.

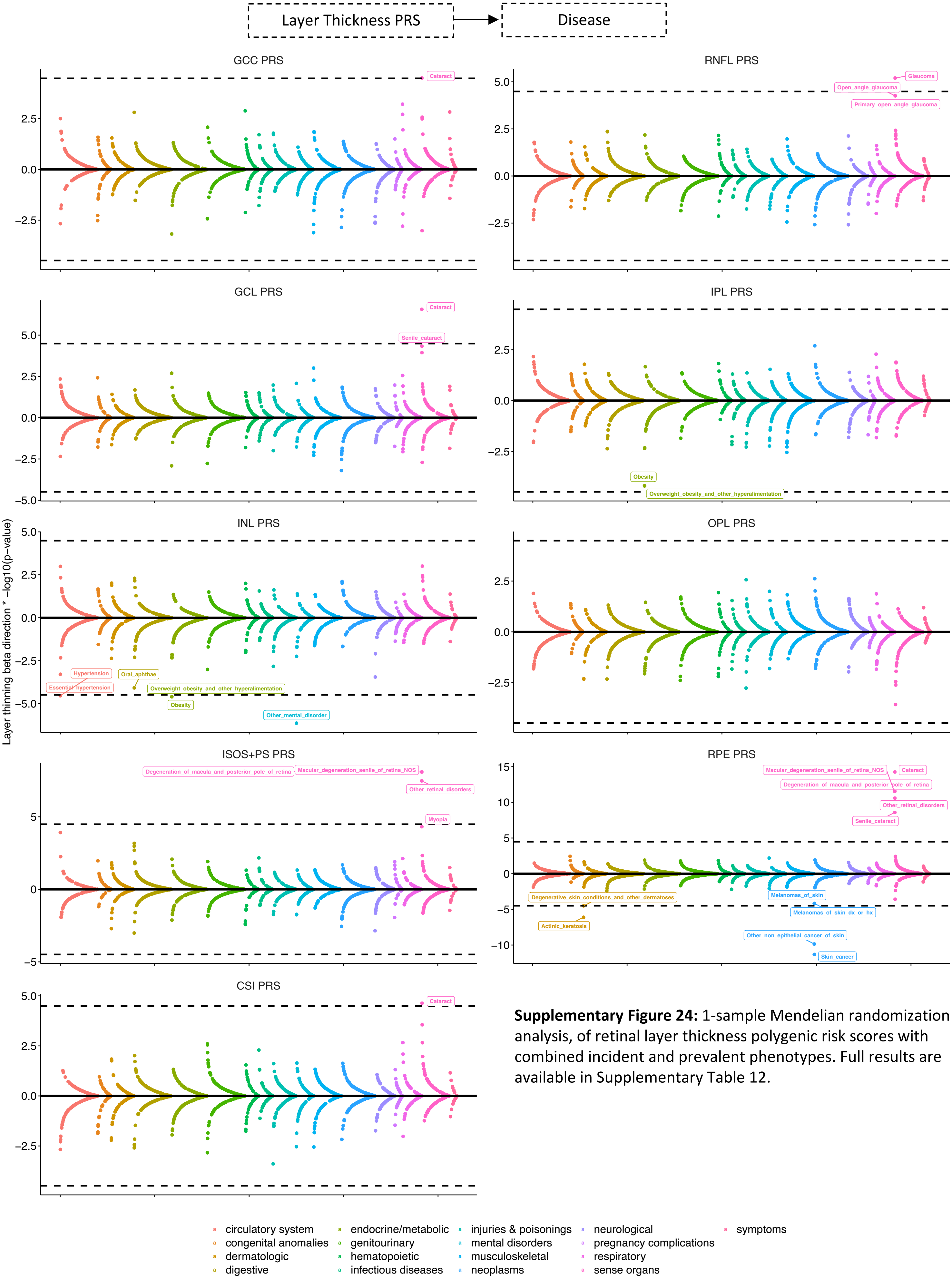


**Supplementary Figure 24:** 1-sample Mendelian randomization analysis, of retinal layer thickness polygenic risk scores with combined incident and prevalent phenotypes. Full results are available in Supplementary Table 12.

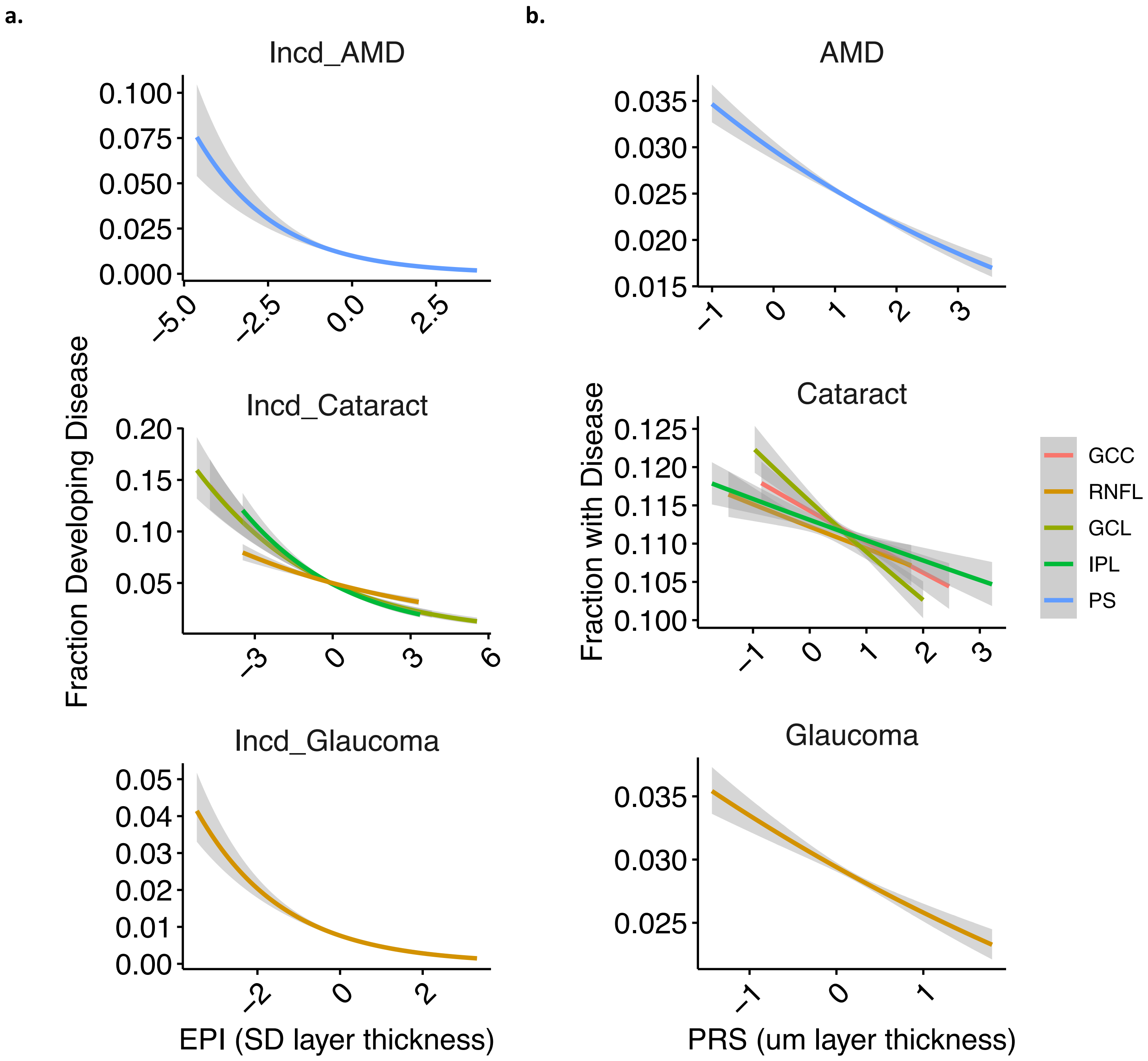


**Supplementary Figure 25:** Comparative epidemiological (a) versus genetic (b) significant and concordant associations of retinal layer thickness with common ophthalmic conditions. Full results after adjustment for the genetic analyses are available in Supplementary Table 12.

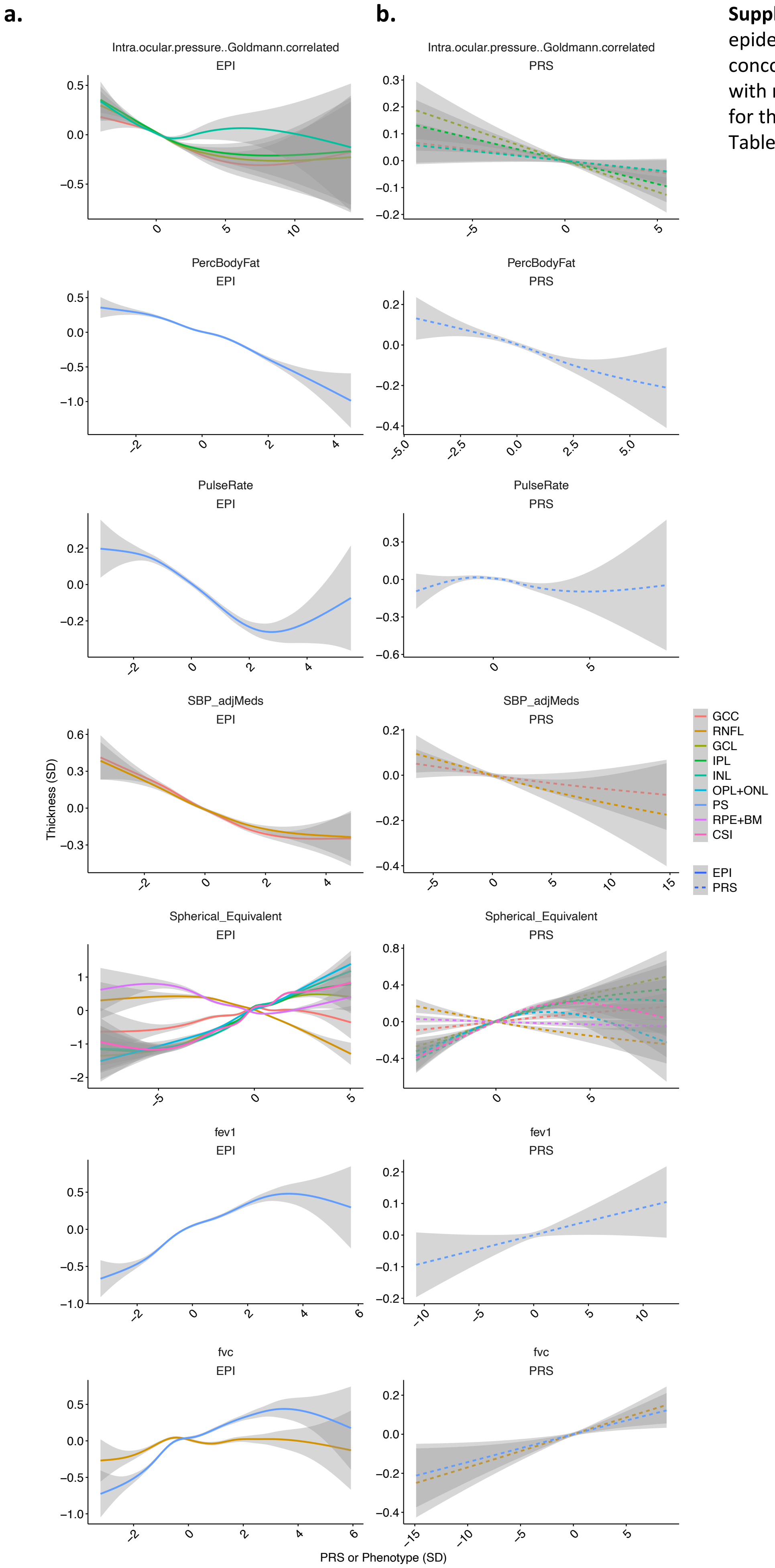


**Supplementary Figure 26:** Comparative (a) epidemiological versus (b) genetic significant and concordant associations of quantitative phenotypes with retinal layer thickness. Full results after adjustment for the genetic analyses are available in Supplementary Table 12.
